# BRAIN-MAGNET: A novel functional genomics atlas coupled with convolutional neural networks facilitates clinical interpretation of disease relevant variants in non-coding regulatory elements

**DOI:** 10.1101/2024.04.13.24305761

**Authors:** Ruizhi Deng, Elena Perenthaler, Anita Nikoncuk, Soheil Yousefi, Kristina Lanko, Rachel Schot, Michela Maresca, Eva Medico-Salsench, Leslie E. Sanderson, Michael J. Parker, Wilfred F.J. van Ijcken, Joohyun Park, Marc Sturm, Tobias B. Haack, Genomics England Research Consortium, Gennady V Roshchupkin, Eskeatnaf Mulugeta, Tahsin Stefan Barakat

## Abstract

Genome-wide assessment of genetic variation is becoming routine in human genetics, but functional interpretation of non-coding variants both in common and rare diseases remains extremely challenging. Here, we employed the massively parallel reporter assay ChIP-STARR-seq to functionally annotate the activity of >145 thousand non-coding regulatory elements (NCREs) in human neural stem cells, modelling early brain development. Highly active NCREs show increased sequence constraint and harbour *de novo* variants in individuals affected by neurodevelopmental disorders. They are enriched for transcription factor (TF) motifs including YY1 and p53 family members and for primate-specific transposable elements, providing insights on gene regulatory mechanisms in NSCs. Examining episomal NCRE activity of the same sequences in human embryonic stem cells identified cell type differential activity and primed NCREs, accompanied by a rewiring of the epigenome landscape. Leveraging the experimentally measured NCRE activity and nucleotide composition of the assessed sequences, we built BRAIN-MAGNET, a functionally validated convolutional neural network that predicts NCRE activity based on DNA sequence composition and identifies functionally relevant nucleotides required for NCRE function. The application of BRAIN-MAGNET allows fine-mapping of GWAS loci identified for common neurological traits and prioritizing of possible disease-causing rare non-coding variants in currently genetically unexplained individuals with neurogenetic disorders, including those from the Genomics England 100,000 Genomes project, identifying novel enhanceropathies. We foresee that this NCRE atlas and BRAIN-MAGNET will help reduce missing heritability in human genetics by limiting the search space for functionally relevant non-coding genetic variation.

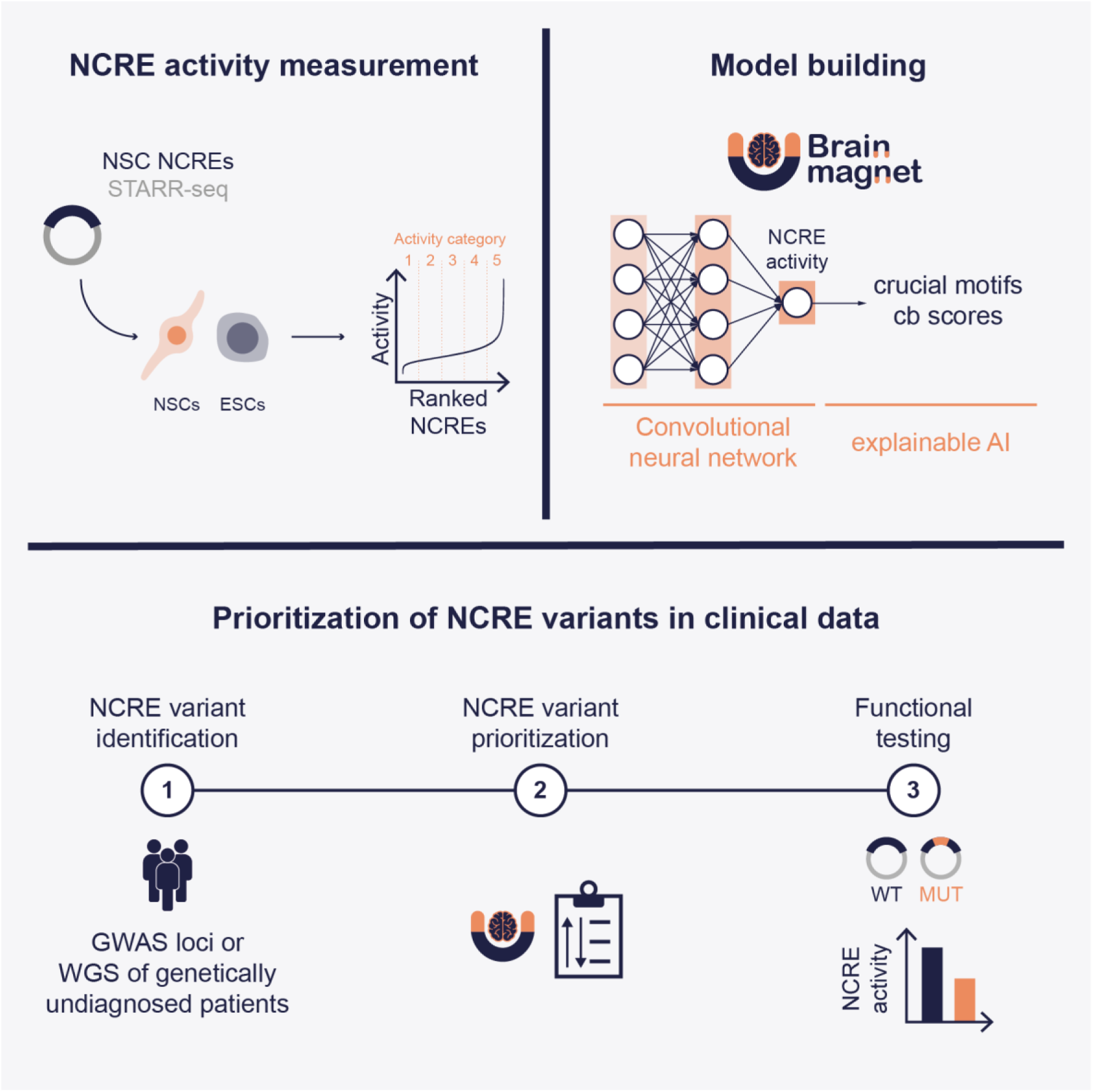

## Introduction

Around 98% of the human genome does not directly encode protein coding genes and contains by far the most genetic variation^1^. Although rapid advances in genomic sequencing technologies have made the identification of this genetic variation rather trivial, interpreting the functionality of non-coding genomic sequences and effects of variants herein remains tremendously challenging^1^. This is exemplified by the plethora of common single nucleotide polymorphisms (SNPs) associated with disease traits by genome-wide association studies (GWAS) for which the underlying biological function and direct effects on gene regulation have not been resolved^2^. Another example concerns the rare disease field, where current genomic diagnostic testing modalities even when using whole genome sequencing (WGS) mostly focus on interpreting potential disease-causing variants directly affecting protein coding genes^3^. Such approaches at best identify a genetic cause for rare disorders in 30-50% of affected individuals^4,5^. Given that in a clinical setting most of the non-coding genetic variation is not routinely assessed, and an increasing amount of evidence demonstrates that genetic alterations of non-coding regulatory elements (NCREs) such as enhancers can cause human disorders^6–8^, it seems likely that a considerable amount of missing heritability in human genetics might be caused by non-coding genetic variation. To resolve this missing heritability, it is crucial to functionally annotate NCREs, characterize their activity in various cell types and to develop novel approaches that help to predict the impact of genomic variants on NCRE function.

Here, we employed the massively parallel reporter assay ChIP-STARR-seq^9^ to functionally annotate NCREs at high throughput and measure their activity in neural stem cells (NSCs) as a model system for developing brain cells. Our in-depth characterisation revealed that increased NCRE activity is associated with increased expression and likelihood of loss-of-function intolerance of the target genes. Highly active NSC NCREs are characterized by increased sequence constraint and are enriched for TF binding motifs including YY1 and p53 family members and primate-specific MER61 and LTR10 transposable elements, providing insights into the mechanisms underlying gene regulation in NSCs. Functional examination of the same genomic regions in human embryonic stem cells (ESCs) identified differential NCRE activity between the two cell types that was accompanied by a rewiring of the epigenome landscape, pinpointing NCREs that are primed for activation upon neural differentiation. Finally, we developed, benchmarked and functionally validated BRAIN-MAGNET, a convolutional neural network model trained on the experimentally measured NCRE activity data to predict nucleotides and motifs required for NCRE activity solely based on DNA composition. We applied BRAIN-MAGNET to prioritize the effects of genomic variants in the context of brain-related neurological traits from GWAS data and to identify likely disease-causing non-coding variants in individuals with rare diseases, including those from the Genomics England 100,000 Genomes Project^10^. Together, these data comprise a novel NCRE atlas and analysis framework that will be invaluable tools for the reduction of missing heritability in human genetics.

## Results

### ChIP-STARR-seq in neural stem cells functionally annotates NCREs with different activity levels

To functionally identify NCREs genome-wide in human ESCs, we previously used the massively parallel reporter assay (MPRA) ChIP-STARR-seq^9^, herein referred to as ESC ChIP-STARR-seq. In this assay, putative NCREs obtained upon chromatin immunoprecipitation (ChIP) are cloned in the STARR-seq reporter plasmid^11^ downstream of a minimal-promoter driven GFP and upstream of the polyadenylation signal. The minimal promoter alone is insufficient to drive GFP expression. However, if the cloned sequence is a functional NCRE, it will enhance GFP transcription and, being located upstream of the polyadenylation signal, it will also be self-transcribed. Thus, performing RNA-seq on GFP-containing mRNAs synthesised upon plasmid cell transfection allows the identification of sequences with functional NCRE activity in the tested cell type.

Here, we applied an adapted ChIP-STARR-seq approach (see **Methods, Fig. 1a, Extended Data Fig. 1** and **Supplementary Note 1** for details) in human NSCs, referred to as NSC ChIP-STARR-seq. NSCs are multipotent stem cells reminiscent of the stem cell population in the ventricular zone during human brain development^12^ and can proliferate and differentiate into multiple neural cell types. Characterizing the functionally active NCRE landscape of NSCs can thus likely identify NCREs involved in early stages of human neurodevelopment, considered relevant for human neurodevelopmental disorders. For ChIP experiments and plasmid library generation we focussed on the TFs YY1 and SOX2, which are important to NSCs^13,14^, and on the histone modifications H3K4me1 and H3K27ac, which are associated with putative NCREs^15^. This allowed us to enrich for sequences that likely reflect active chromatin at the endogenous loci in NSCs even when tested in an episomal MPRA.

**Fig. 1:**
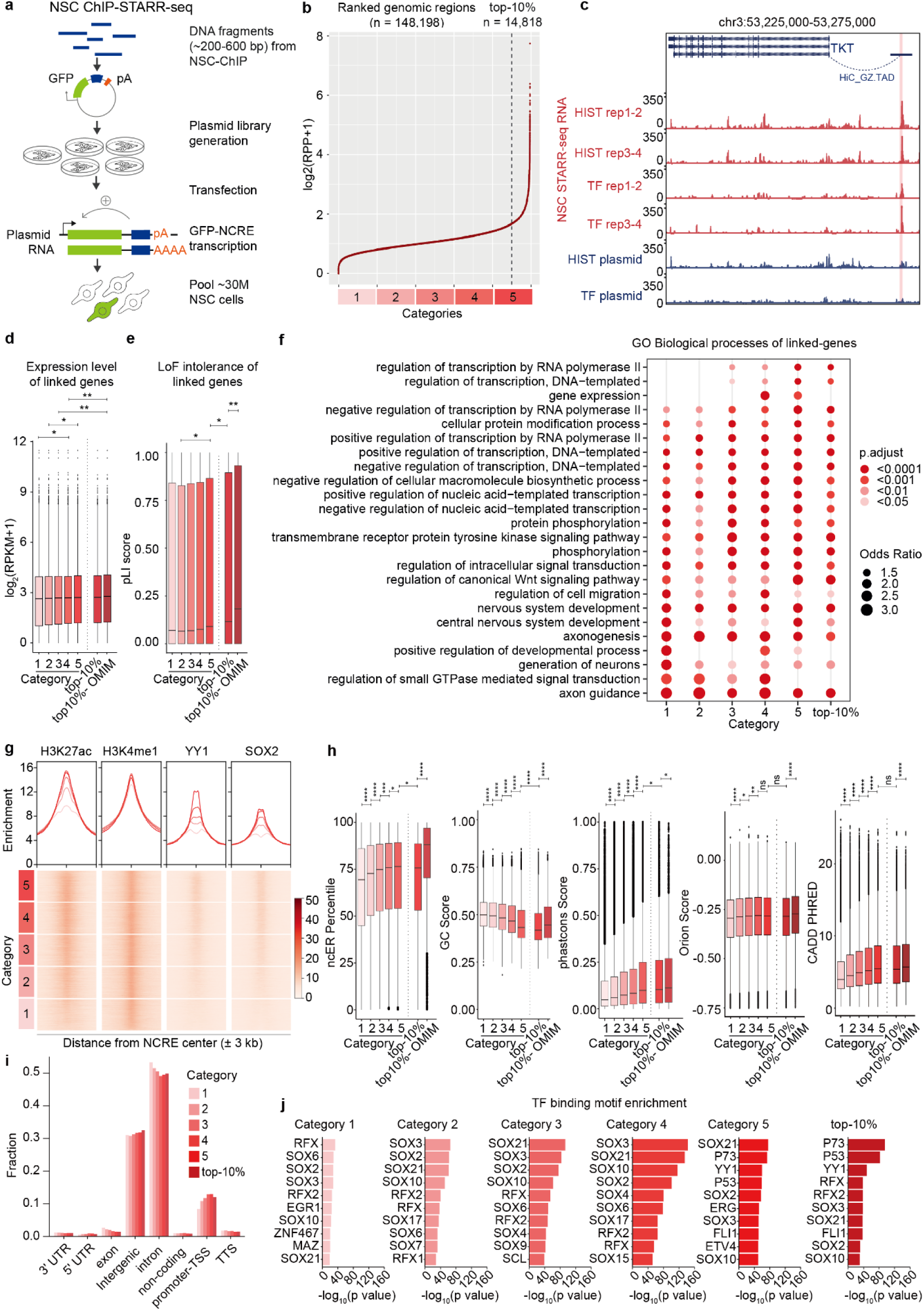
C**h**IP**-STARR-seq in neural stem cells identifies NCREs with different activity levels and sequence characteristics** a) Schematic representation of the experimental strategy used in this study. b) Line plot showing NCRE activity (enrichment of ChIP-STARR-seq RNA over plasmids per region of the scaffold; log2) of the 148,198 assessed scaffold regions, ranked from lowest to highest activity. In gradient of red are depicted the 5 categories (groups of 20 percentiles) of increasing NCRE activity detected by NSC ChIP-STARR-seq used throughout this study. The dotted line demarcates the top-10% most active NCREs. c) Genome browser tracks surrounding the *TKT* locus showing a distal NCRE showing activity by NSC ChIP-STARR-seq and interaction of this NCRE with the *TKT* gene confirmed by HiC from foetal brain. Blue tracks: sequencing of plasmid DNA. Red tracks: sequencing of ChIP- STARR-seq RNA. d) Expression level (log2(RPKM+1)) of the genes linked to each of the 5 NCRE activity categories, to the top-10% most active NCREs and of the OMIM disease-related genes linked to the top-10% most active NCREs. Boxes represent the interquartile range (IQR); lines represent the median; whiskers extend to 1.5 the IQR; dots represent outliers. (* p < 0.05; ** p < 0.01, Wilcoxon test). e) As in d) showing the pLI score of the genes linked to the different NCRE categories. (* p < 0.05; *** p < 0.001 Wilcoxon test). f) Gene ontology analysis using Enrichr of biological processes for the protein-coding genes linked to the NCREs of the 5 categories and the top-10% most active NCREs. g) Top: line plot showing the average enrichment of ChIP-seq signal from NSCs across the genomic regions encompassed by NCREs from the 5 categories of NCRE activity (bins of 3 kb up and down centred at the middle of the NCRE). Bottom: corresponding heatmaps of H3K27ac, H3K4me1, YY1 and SOX2 ChIP-seq signals in NSCs across the 148,198 assessed scaffold regions ranked by activity from the highest to the lowest. h) Box plots, showing from left to right ncER percentile, GC content score, phastcons score, Orion score, and CADD score, for all the 5 NCRE activity categories, the top-10% most active NCREs (top-10%) and the top-10% most active NCREs linked to genes with a known OMIM phenotype. Boxes represent the interquartile range (IQR); lines represent the median; whiskers extend to 1.5 the IQR; dots represent the outliers. (* p < 0.05; ** p < 0.01; *** p < 0.001; **** p< 0.0001; ns not significant; Wilcoxon tests). i) Bar graphs showing the fraction of NCREs per genomic region using Homer, for all NCRE categories. UTR = untranslated region; non-coding = non-coding RNAs such as snRNA, lncRNA, etc; TSS = Transcriptional start site; TTS = Transcriptional termination site. j) Bar graph showing the 10 most enriched TF motifs in the NCREs belonging to the different activity categories in NSCs. Plotted is the -log10 p-value.

To identify active NCREs, we first generated an analysis scaffold consisting of all merged ChIP-seq and plasmid DNA-seq peaks (148,198 genomic regions in total, see **Methods**). Large merged sequences were split in scaffold regions of a maximum of 1,000 bp, which were approximately equally divided over all human chromosomes except for the Y chromosome, which was not present in the female cell line used for the experiments (**Extended Data Fig. 2**a). Subsequently, we counted ChIP-STARR-seq RNA and plasmid reads in each of these scaffold regions. The ratio of normalized RNA-seq to DNA-seq (plasmid) reads per scaffold region was then used as a measure of NCRE activity of that region. This allowed ranking of NCREs based on their activity level in different categories (**Fig. 1b**)^16^. Visual inspection of RNA and plasmid tracks at multiple loci confirmed the presence of NCREs, as testified by the loci surrounding the *TKT* and *CHD8* genes, which showed NCREs ranking within the top-10% activity category (**Fig. 1c and Extended Data Fig. 2**b). Independent testing of 15 NCREs confirmed their NCRE activity (**Extended Data Fig. 2**c). To assess whether NCRE activity correlates with the expression of putative target genes in NSCs, we first linked NCREs to their presumed target genes by combining the closest gene method and available Hi-C data obtained from the germinal zone of human foetal brain at gestation weeks 17–18^17^, representing the anatomical location of NSCs in the developing human brain. We observed that genes linked to NCREs with higher activity show a significantly higher expression level compared to genes linked to regions with lower NCRE activity, reaching the highest expression level when focussing on the top-10% most active NCREs (n=14,818 NCREs, n=7,752 unique genes) (**Fig. 1d**). Gene expression further increased when focussing on the subset of genes involved in human disease according to the *Online Mendelian Inheritance in Man* (OMIM) catalogue (**Fig. 1d**). As several NCREs with different activity levels can be linked to the same gene, we performed the same analysis using only expression data for those genes uniquely linked to each NCRE activity group and observed similar findings (**Extended Data Fig. 2**d).

Loss of function (LoF) of human genes has been studied as a driver of human disease. We investigated the probability of LoF intolerance (pLI) of the genes linked to the different NCRE activity categories and observed that genes regulated by highly active NCREs, besides being expressed at a higher level, are also more intolerant to LoF, suggesting they might play important biological roles (**Fig. 1e**). Gene ontology analysis using Enrichr^18,19^ of the protein-coding genes linked to the various NCRE activity groups revealed that genes linked to the most active NCREs are mainly involved in the regulation of transcription, whereas genes involved in various processes such as neurodevelopment and axon guidance were linked to NCREs belonging to all activity classes (**Fig. 1f**).

Next, to investigate whether the presence of a certain histone modification (H3K27ac or H3K4me1) or the binding of one of the tested TFs (YY1, SOX2) would largely predict NCRE activity, we analysed the ChIP-seq signal enrichment across the different NCRE activity categories. We observed that increased NCRE activity was associated with an increased H3K27ac, YY1 and SOX2 ChIP-seq signal, while the H3K4me1 signal was rather constant across all the activity categories (**Fig. 1g**). Looking at the ratio between the overlap of each NCRE category with each ChIP-seq and the total overlap of the 148,198 scaffold regions with each ChIP-seq, both normalized for the group size, we observed the same trend (**Extended Data Fig. 2**e).

We conclude that by employing NSC ChIP-STARR-seq, we can rank NCREs according to their activity, which correlates with the expression level of presumed NCRE target genes of cell-type relevance.

### Active NCREs in neural stem cells show distinguishing sequence characteristics

To further characterize NCREs active in NSCs, we focused on their sequence characteristics and investigated a variety of *in silico* metrics, including GC content. The non-coding essential regulation (ncER) score measures the essentiality of each nucleotide within a given genome sequence, ranging from 0 (non-essential) to 1 (putative essential)^20^; the phastcons score indicates the probability of each nucleotide of a given sequence to be conserved among multiple species^21^; the Orion score is based on the depletion of variation in the human population^22^; and finally the CADD score takes into account the likelihood of a given nucleotide variant to be deleterious^23^. We observed that NCRE activity in NSC ChIP-STARR-seq is positively associated with an increase in all these scores except for GC content (**Fig. 1h**), which is in line with previous findings showing that cell-type specific NCREs have lower GC content compared to more broadly active NCREs^24,25^. When focussing on the top-10% most active NCREs linked to genes associated with known OMIM phenotypes, we noticed a further increase in these scores (**Fig. 1h**). We previously performed a large-scale integrative computational analysis of epigenome datasets during foetal brain development^26^ and observed that genes linked to differentially active enhancers (as measured by variability of epigenome marks over time) are less tolerant to LoF, whereas non-differentially active regions are more tolerant to LoF. We therefore wondered whether NCRE activity measured by NSC ChIP-STARR-seq would also correlate with intolerance to LoF. To investigate this, we used previously published LoF tolerance scores determined from structural variants of whole genome sequences^27^. We observed that the investigated scaffold regions largely show intolerance to LoF, without a clear difference emerging between the different NCRE activity groups (**Extended Data Fig. 2**f). This possibly reflects the fact that the investigated scaffold regions were selected for being endogenously marked by H3K27ac, H3K4me1, or bound by YY1, or SOX2 and thus might already be enriched for regions with intolerance to LoF, independent of their NCRE activity in NSCs. Next, we investigated the localization of NCREs relative to the transcription start site (TSS) of genes and found that NCREs belonging to all the categories of activity show a similar distance distribution (**Fig. 1i**) and location as annotated by Homer (**Extended Data Fig. 2**g). It is important to note that the design of the STARR-seq reporter construct, with the NCRE being positioned downstream of the reporter open reading frame, does not allow the measurement of promoter activity. Thus, even when an active region is located in proximity to a TSS, the observed NCRE activity reflects enhancer activity and not promoter activity.

To determine whether differential TF binding might influence NCRE activity, we assessed the top-10 TF binding motifs in each NCRE activity category. Lower activity categories were mainly enriched for motifs of various SOX family TFs important for neural development, and motifs of the RFX family, involved in the regulation of differentiation of various cell types^28^, while the top-10% most active NCRE category was characterized by p53 family motifs (p53 and p73), that play a crucial role in regulating the development of the central nervous system^29,30^ and the YY1 motif (**Fig. 1j**). Also enriched in the top-10% most active NCREs was the Fli1 motif, belonging to the ETS family that regulates a wide variety of processes^31^.

Finally, we assessed the presence of transposable elements (TEs) at the identified NCREs. TEs are present throughout the human genome and are enriched in TF binding sites^32^. Moreover, in our previous work^9^, we showed that some classes of TEs are enriched in active NCREs in ESCs, including satellite repeats and some LTR family elements^33^. The most enriched TEs in the top-10% most active NCREs in NSCs belong to the primate-specific subfamilies of MER61 and LTR10, both members of the ERV1 type of long terminal repeats (LTRs) (**Extended Data Fig. 2**h). These TEs have previously been associated with p53 binding sites^34,35^, in line with our observation that p53 binding sites are enriched in highly active NCREs (**Fig. 1j**). On the other hand, TEs enriched in the classes with low NCRE activity are mostly simple repeats (**Extended Data Fig. 2**h). Importantly, we did not observe any overlap between the 10 most enriched TEs in NSCs and the 10 most enriched TEs we identified previously in ESC ChIP-STARR-seq^9^, suggesting that the NSC NCRE landscape is shaped mainly by different TEs. However, our top hits in NSCs, MER61C and MER61E, were included in the top-20 most enriched TEs in active NCREs in ESCs.

Together this indicates that NCREs from different NCRE activity categories, despite being located at similar genomic locations relative to TSSs, show differences in sequence characteristics including sequence constraint, and enrichment for TF motifs and TEs.

### Comparative ChIP-STARR-seq using NSC-derived plasmid libraries transfected in ESCs identifies NCREs which are primed in ESCs for future NCRE activity

Reasoning that NCREs with a role during neural development might show higher NCRE activity specifically in NSCs whereas NCREs regulating genes with broader biological roles might show activity in multiple cell types, we next investigated cell-type specificity of NCRE activity. Therefore, we transfected ESCs with the same NSC-derived ChIP-STARR-seq plasmid libraries (**Fig. 2a and Extended Data Fig. 1**i,j), which we refer to as comparative ChIP-STARR-seq. In NSCs, these sequences are marked by H3K27ac, H3K4me1, YY1 or SOX2 resulting in their capture into the NSC plasmid library. Testing their activity in an earlier developmental cell type such as ESCs, where these sequences are not necessarily endogenously enriched for active chromatin marks, might additionally allow direct assessment of the role of the nucleotide composition itself driving NCRE activity. As done for NSCs, we ranked the 148,198 NSC-derived scaffold regions based on the NCRE activity in ESCs (**Fig. 2b**). As expected we found active NCREs, given that some NCREs show activity in multiple cell types and because 56.35% of the NSC-derived scaffold overlapped with the ESC-derived scaffold tested in ESC ChIP-STARR-seq^9^, representing genomic regions marked by OCT4, NANOG, H3K27ac and H3K4me1 in ESCs (**Extended Data Fig. 3**a). First, comparing for each of the 148,198 scaffold regions the changes in NCRE activity groups between NSC ChIP-STARR-seq and comparative ChIP-STARR-seq, we noticed that most regions remained in the same or in an adjacent higher or lower activity category, and only a minority showed extreme changes (e.g., going from category 5 to category 1 and vice versa) (**Fig. 2c**). To further interpret comparative ChIP-STARR-seq findings, we linked NCREs to presumed target genes using proximity and available HiC data in H9 ESCs^36^. Contrary to NSCs, we observed that increased NCRE activity in comparative ChIP-STARR-seq is not accompanied by a major increase in gene expression of the presumed target genes in ESCs (**Fig. 2d**). Like in NSCs, we observed an increased intolerance to LoF of the presumed ESC target genes (**Fig. 2e**). Focussing on sequence characteristics, we did not notice a consistent stepwise increase of the various scores following increased comparative ChIP-STARR-seq NCRE activity as found in NSCs, with the exception of the phastcons and CADD score. Rather, we observed a decrease in essentiality (ncER), Orion and GC score, the latter mirroring our findings in NSCs (**Fig. 2f**). Next, we investigated the localization of NCREs relative to the TSS of genes and noticed that NCREs with increased activity in comparative ChIP-STARR-seq in ESCs tended to be more distally located (**Extended Data Fig. 3**b). When performing TF binding motif analysis, whereas the top-10% most active NCRE category in NSCs was dominated by motifs for p73, p53 and YY1 (**Fig. 1j**), these motifs were no longer found amongst the top-10 most enriched motifs in any of the NCRE activity categories from comparative ChIP-STARR-seq in ESCs (**Extended Data Fig. 3**c). Other TF motifs, including SOX2, SOX3, SOX10, RFX and RFX2 were enriched in the top-10% category in ESCs, like in NSCs, while ERG and ETV2, other members of the above mentioned ETS family, were only found enriched in the top-10% most active NCRE category in ESCs but not in NSCs (**Extended Data Fig. 3**c). Interestingly, except for the OCT4-SOX2-TCF-NANOG motif only found in NCRE category 5 of comparative ChIP-STARR-seq in ESCs, we did not observe major differences in TF motif enrichment between NCREs ranked in the 5 activity categories in ESCs, as they all showed SOX and RFX family TFs (**Extended Data Fig. 3**c). When investigating the 10 most enriched TEs in the different categories of NCRE activity in comparative ChIP-STARR-seq, we observed similar results as for the NSCs (**Extended Data Fig. 3**d), with a large overlap of TEs identified in highly active NCREs in both cell types.

**Fig. 2:**
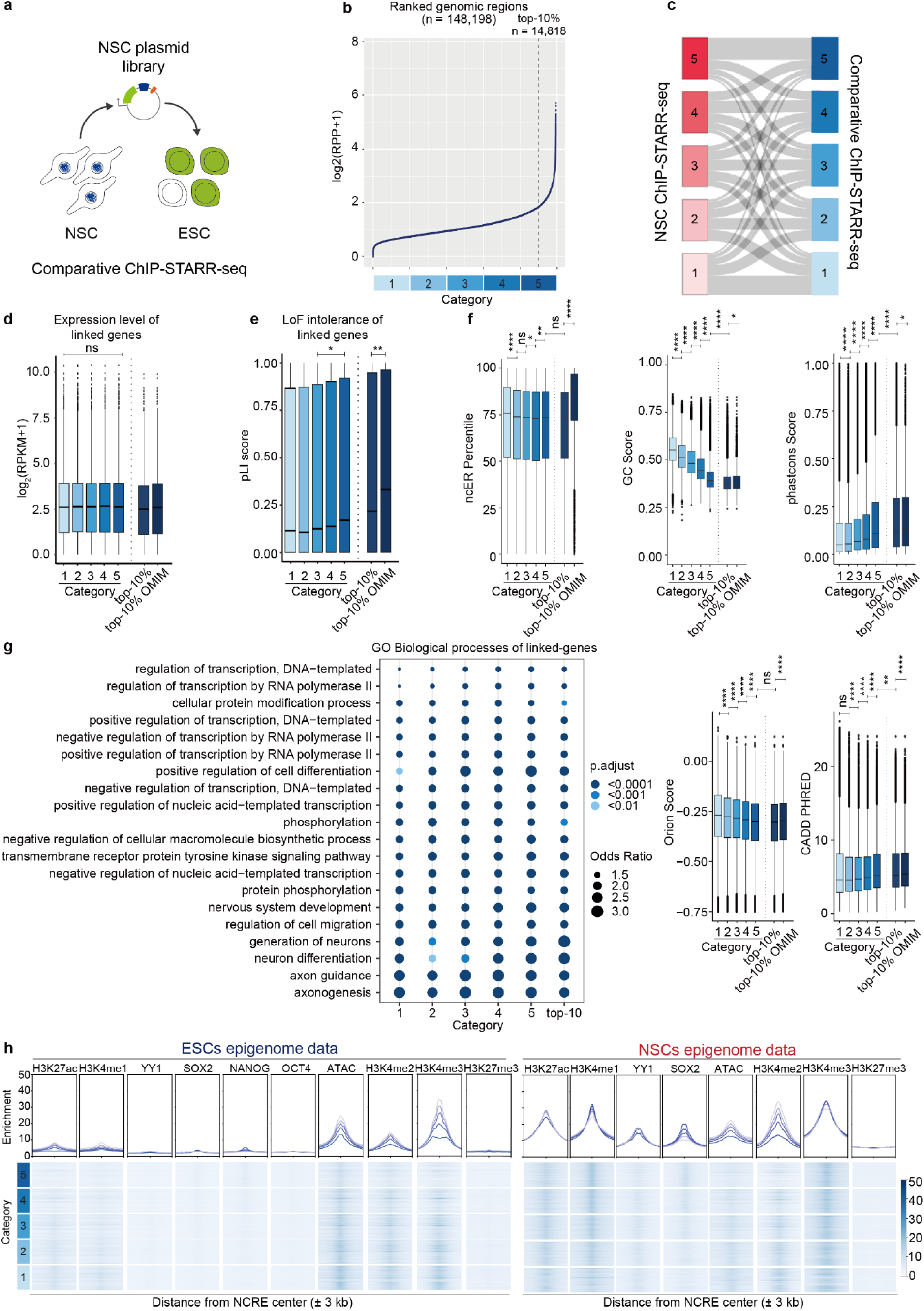
Comparative ChIP-STARR-seq using NSC derived plasmid libraries in ESCs identifies NCREs which are primed for future NCRE activity a) Schematic representation of the experimental strategy used for transfecting NSC derived ChIP-STARR-seq libraries in ESCs, referred to as comparative ChIP-STARR-seq. b) Line plot showing NCRE activity (enrichment of ChIP-STARR-seq RNA over plasmids per region of the scaffold; log2) of the 148,198 assessed scaffold regions, ranked from lowest to highest activity from comparative ChIP-STARR-seq in ESCs. In gradient of blue are depicted the 5 categories (groups of 20 percentiles) of increasing NCRE activity used throughout this study. The dotted line demarcates the top-10% most active NCREs. c) Sankey plot showing the changes between activity categories for the NCREs measured in NSC ChIP-STARR-seq (red) and in comparative ChIP-STARR-seq (blue). For the latter, we transfected the NSC-derived plasmid libraries in ESCs. d) Expression level (log2(RPKM+1)) of the genes linked to each of the 5 NCRE activity categories, to the highly active NCREs (top-10%) and to the highly active NCREs linked to genes with a known OMIM phenotype. Boxes represent the interquartile range (IQR); lines represent the median; whiskers extend to 1.5 the IQR; dots represent outliers. (* p < 0.05; Wilcoxon test). e) As in d) showing the pLI score of the genes linked to the different NCRE categories. (** p < 0.01; Wilcoxon test). f) Box plots as in d), showing from left to right on the upper row ncER percentile, GC content score, phastcons score and on the lower row Orion score, and CADD score. (* p < 0.05; ** p < 0.01; **** p< 0.0001; ns not significant; Wilcoxon tests;). g) Gene ontology analysis using Enrichr of biological processes for the protein-coding genes linked to the NCREs of the 5 categories and the top-10% most active NCREs. h) Epigenome profiling of NCREs with comparative ChIP-STARR-seq activity in ESCs (plasmid libraries derived from ChIP experiments in NSCs). Top: line plot showing the average enrichment of ChIP-seq (or ATAC-seq) signal from ESCs (left panel) or NSCs (right panel) across the genomic regions encompassed by NCREs from the 5 categories of NCRE activity from comparative ChIP-STARR-seq in ESCs (bins of 3 kb up and down centred at the middle of the NCRE). Bottom: corresponding heatmaps of H3K27ac, H3K4me1, YY1, SOX2, NANOG, OCT4, H3K4me2, H3K4me3 and H3K27me3 ChIP-seq signals and ATAC-seq in ESCs (left), or H3K27ac, H3K4me1, YY1, SOX2, H3K4me2, H3K4me3 and H3K27me3 ChIP- seq signals and ATAC-seq in NSCs (right) across the 148,198 assessed scaffold regions ranked by activity as measured by comparative ChIP-STARR-seq in ESCs from the highest to the lowest.

Gene ontology analysis of the protein-coding genes linked to the various classes of ESC NCRE activity showed that genes involved in the regulation of transcription are enriched amongst all ESC NCRE activity categories (**Fig. 2g**). Categories with higher NCRE activity in comparative ChIP-STARR-seq were also enriched for processes involved in nervous system development and neuronal differentiation (**Fig. 2g**). This was surprising given that these experiments were performed in ESCs where we would have expected to find a more ESC-related gene ontology for genes linked to the most active NCREs. Together with the lack of a major increase in target gene expression in ESCs upon increased NCRE activity from comparative ChIP-STARR-seq in ESCs (**Fig. 2d**) and the absence of the typical ESC TF and TE enrichments that we previously observed in ESC ChIP-STARR-seq with ESC-derived libraries^9^, this potentially indicates that the NCREs showing episomal activity in comparative ChIP-STARR-seq in ESCs might not all be active at the endogenous loci in ESCs. This could possibly reflect the NSC origin of the plasmid libraries and the fact that these sequences in the ESC experiments are uncoupled from their cell type specific environment and chromatin landscape. Thus, they might not be active at the endogenous loci in ESCs, in contrast to NSCs where these NCREs are endogenously marked by H3K27ac, H3K4me1, YY1 or SOX2. Indeed, when determining the ChIP-seq signal enrichment obtained from ESC data for the NCREs from the various activity categories from comparative ChIP-STARR-seq in ESCs, we only noticed a limited enrichment of H3K27ac and H3K4me1 in ESCs at these sites, not correlating with their measured NCRE activity (**Fig. 2h**). Likewise, enrichment of YY1, SOX2, OCT4 and NANOG at these sites in ESCs was limited. Both of these trends were slightly increased when assessing only those scaffold regions showing an overlap between the NSC-derived scaffold used herein and the previously used ESC-derived scaffold from ESC ChIP-STARR-seq^9^ (**Extended Data Fig. 3**e), but still dramatically reduced when compared to the enrichment of H3K27ac, H3K4me1, YY1 and SOX2 in NSCs over these regions (**Fig. 2h and Extended Data Fig. 3**e). Surprisingly, despite the lack of H3K27ac and H3K4me1 in ESCs, these regions were still located in accessible chromatin in ESCs, as determined using ATAC-seq, and depleted for the repressive mark H3K27me3 in ESCs (**Fig. 2h**). Additionally, these same regions were enriched for H3K4me2 and H3K4me3, with the latter being more enriched in the less active NCRE categories from comparative ChIP-STARR-seq in ESCs (**Fig. 2h**). Previously, so called “primed enhancers” have been shown to lack H3K27ac and to be characterized by H3K4 methylation^37^. Together, the epigenome profile indicates that some of the NCREs identified by comparative ChIP-STARR-seq belong to such a class of primed NCREs, which are primed in ESCs for activity at later developmental stages. An extensive differential NCRE activity analysis and epigenome profiling (**Supplementary Note 2 and Extended Data Fig. 4**), comparing NCREs with high activity in the one cell type and low activity in the other, and vice versa, likewise pointed to priming of NCREs for later activity along the trajectory of neural differentiation, although future studies will be required to completely resolve the underlying mechanisms.

### BRAIN-MAGNET, a convolutional neural network model, predicts NCRE activity based on DNA sequence and facilitates interpretation of biological consequences of non-coding variants

The NSC ChIP-STARR-seq performed herein provides a comprehensive atlas of annotated NCREs ranked by activity in a cell type relevant to early brain development. We hypothesized that such an NCRE atlas will aid the interpretation of non-coding genetic variants encountered in neurodevelopmental disorders. The diagnostic yield for most of these disorders using current genomic technologies, which mainly focus on coding sequences, is well under 50%^4,5^, suggesting that some causes of missing heritability might be located in NCREs. If this would be the case, disease-relevant NCREs should be relatively depleted of rare genomic variation in the healthy population compared to other genome sequences. Indeed, using the recently released gnomAD v4 data, containing 76,215 genomes from individuals that did not present with early onset neurodevelopmental phenotypes^1^, we found that NCREs with higher activity in NSCs harboured fewer rare variants (minor allele frequency (MAF) <0.1%) compared to NCREs with lower activity (**Extended Data Fig. 5**). An even stronger depletion of rare variants was observed upon increasing NCRE activity in comparative ChIP-STARR-seq (**Extended Data Fig. 5**). As expected, the coding sequences (CDS) of protein coding genes showed the strongest depletion of rare variants. Nevertheless the relative depletion of rare variants in the highly active NCREs in both NSCs and ESCs was in a similar range as for other sets of NCREs, including VISTA enhancers^38^, candidate *cis* regulatory elements from ENCODE^39^, differentially active enhancers (DAEs) and non-differentially active enhancers (nDAEs) from an epigenome study of human foetal brain^26^, and for the 5’ and 3’ UTRs of genes (**Extended Data Fig. 5**). Although this relative depletion of rare variants is in line with the hypothesis that the identified NCREs from our atlas could be disease relevant, the 148,198 NCREs assessed, encompassing a total size of 112,314,966 nucleotides, still harboured 28,486,102 rare variants (MAF <0.1%) in healthy individuals. Most of these variants are likely not disease relevant as they will not interfere with NCRE function, but it is currently difficult to assess their potential pathogenicity, especially given the lack of a genetic code for NCREs contrary to that of the protein coding exome. If a rare variant would affect a nucleotide directly relevant for the NCRE function and consequent downstream gene expression, it would be more likely to be disease relevant. This underscores the necessity to develop tools that can predict the function of individual nucleotides within NCREs, enabling the prioritization of variants that are more likely to be pathogenic.

To be able to interpret the potential effects of NCRE variants, we set out to build an artificial intelligence based prediction model. As identifying non-coding variants that affect NCRE function is reminiscent of searching for needles in a haystack, and needles are more likely to be identified using a magnet, we refer to the model as BRAIN-MAGNET (*BRain-focussed Artificial INtelligence Method to Analyse Genomes for Non-coding regulatory Element mutation Targets*). BRAIN-MAGNET is a sequence-based single-task convolutional neural network (CNN) model^40^ that allows predicting NCRE activity for NSCs and ESCs individually (**Fig. 3a, Extended Data Fig. 6**b,c**, Methods**). This model uses the genomic sequences assessed by ChIP-STARR-seq as input and links to their measured activity as output, determining what is causing a given sequence to have a given activity. BRAIN-MAGNET trained on both NSC ChIP-STARR-seq and comparative ChIP-STARR-seq in ESCs is able to precisely predict the experimentally measured activity of NCREs, contrary to an existing model (**Extended Data Fig. 6**a-c), with the ESC-trained model slightly outperforming the NSC- trained model (**Extended Data Fig. 6**b,c). Possibly this reflects the fact that some NCREs identified by comparative ChIP-STARR-seq in ESCs potentially are primed, and might be more dependent on their sequence composition for episomal activity, which might be more amenable for training of the sequence-based model.

**Fig. 3:**
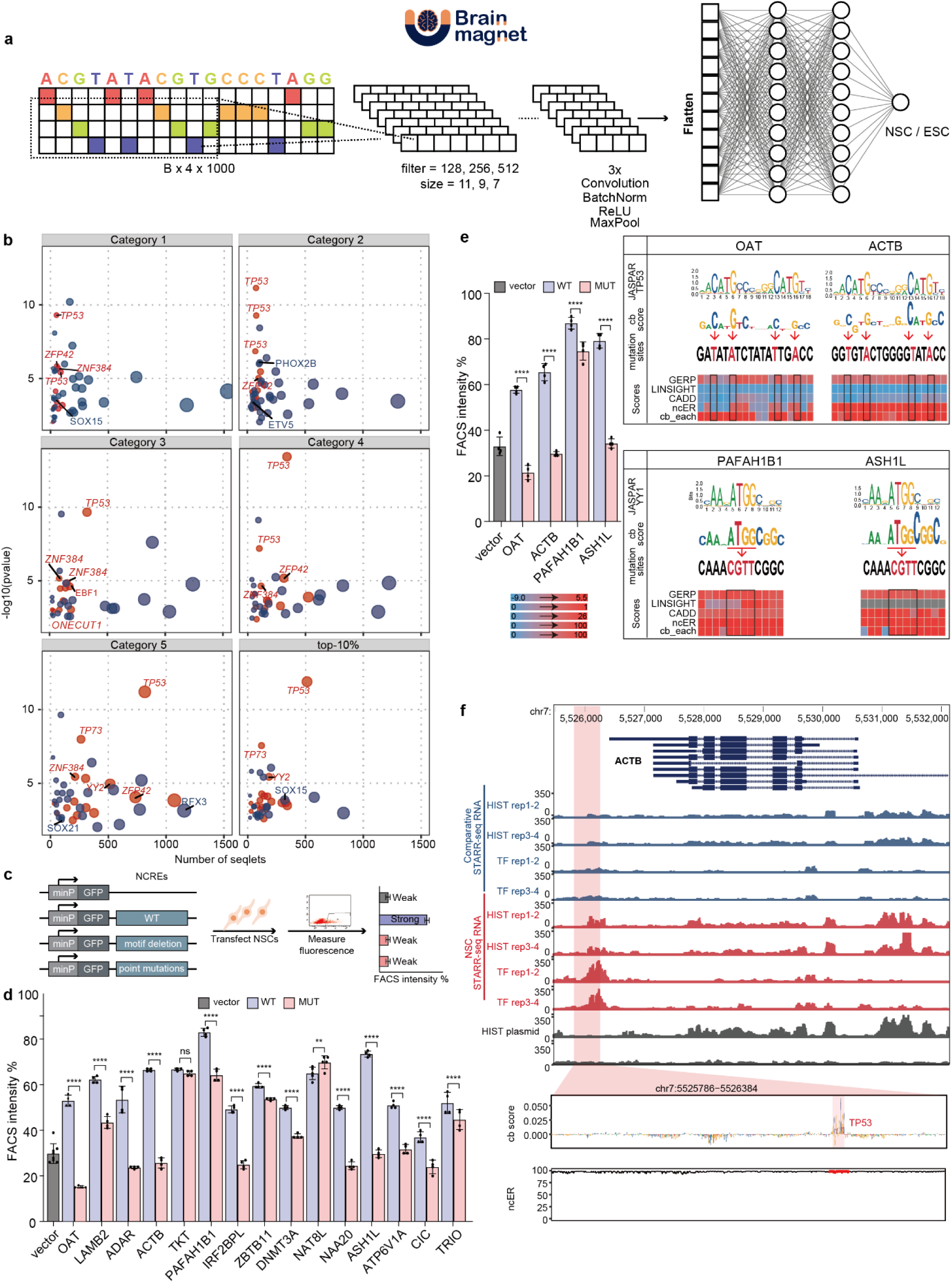
BRAIN-MAGNET, a sequence based convolutional neural network model, predicts NCRE activity and facilitates interpretation of biological consequences of genomic variation a) BRAIN-MAGNET is a deep convolutional neural network architecture trained to predict NCRE activity from an input sequence of the tested element. See **Extended Data Fig. 6** for performance and benchmarking analysis of BRAIN-MAGNET. b) Enrichment of TF motifs in sequences with high BRAIN-MAGNET cb scores of the various NSC NCRE activity categories discovered by TF-MoDISco-lite^41^. The number of seqlets compared with significance (-log10 p-value) for the most matched TF motif per motif cluster. Red dots denote enriched motifs in sequences with positive cb scores, and blue dots represent motifs enriched at sequences with negative cb scores. The motifs with FDR < 0.05 are highlighted as *italic*. c) Schematic drawing of the experimental validation strategy for BRAIN-MAGNET predictions. d) Bar plot showing the percentage of GFP+ cells in NSCs upon cell transfection experiments (as outlined in c)) with reporter plasmids containing either wild type (blue) or motif deleted NCREs (pink) (n=15) or empty reporter plasmid control (grey). NCREs are indicated with the name of the presumed target gene. Plotted is the percentage of GFP+ in cells co-transfected with an mCherry expressing plasmid, to correct for transfection efficiency. Bars show the average from two independent experiments, with each NCRE tested in duplicate. Error bars represent standard deviation. * p < 0.05; ** p < 0.01; *** p < 0.001; **** p < 0.0001 (one-way ANOVA test followed by multiple comparison test (Fisher’s LSD test). e) As d), but now showing 4 NCREs with point mutations of the predicted functional relevant motifs. Right insert shows the predicted motifs from JASPAR, a visual representation of the cb scores for each nucleotide, the created point mutations of the motifs that were tested and other variant prioritization outputs from LINSIGHT, CADD, GERP and ncER scores for the same region displayed in a heatmap. f) Genome browser view showing the *ACTB* locus and the upstream NCRE tested in panel d and e for which BRAIN-MAGNET predicts a functional motif. Top panel: genome browser tracks showing ChIP-STARR-seq RNA in ESCs (blue) and NSCs (red) and sequencing of plasmid (grey); highlighted region in magenta indicates the NCRE. The zoom-in of the NCRE shows the BRAIN-MAGNET contribution (cb) scores from the NSC model and the ncER scores of the same region. A region with high cb scores and an overlapping TP53 motif is highlighted.

To further interpret the predictions of BRAIN-MAGNET, we next utilized the explainable AI DeepExplainer framework^41,42^ to calculate BRAIN-MAGNET contribution (cb) scores. These cb scores indicate for each of the 112,314,966 nucleotides in our NCRE atlas the relative contribution to NCRE activity, with higher cb score ranking pointing to functionally more important nucleotides. We then identified functionally important motifs in sequences with high cb scores using TF-MoDISco-lite (see **Methods**) (**Fig. 3b**). For example, in the NSC NCREs we found prominent enrichment for the TP53 and TP73 motifs when using BRAIN-MAGNET trained on NSC ChIP-STARR-seq (**Fig. 3b**). Another motif frequently identified was ZFP42 (REX1). REX1 originated from a duplication event of YY1 in the eutherian lineage^43^, thus it recognizes the same TF binding motif. Since REX1 is not expressed in NSCs, the observed enrichment for the ZFP42 motif likely represents YY1 bindings sites. Indeed, the YY1 motif is the third redundant motif called by the algorithm at the ZFP42 motif sites. Hence, the results from BRAIN-MAGNET are in line with the previous motif discovery analysis (**Fig. 1j**), but additionally indicate that the binding of these TFs might also be functionally important for NCRE activity given the high cb scores of these motifs. To functionally validate BRAIN-MAGNET findings and to confirm that the prioritized motifs were indeed required for NCRE activity, we selected 15 NCREs highly active in NSCs that displayed a significant TF motif with high cb score (**Fig. 3c,d, Extended Data Fig. 7**-12). We cloned these NCREs into the STARR-seq reporter plasmid and generated mutations lacking 30 bp centred on the TF motif. Upon transfection in NSCs, we observed that these deletions significantly impacted the NCRE activity of 14 out of 15 tested NCREs (**Fig. 3c,d and Extended Data Fig. 7**-12). Likewise, generating point mutations disrupting the prioritized motifs of 4 NCREs similarly affected NCRE activity (**Fig. 3c,e and Extended Data Fig. 7**,8). For example, changing 4 nucleotides in the prioritized TP53 motif of an NCRE upstream of *ACTB* (**Fig. 3e,f**), a gene of which mutations cause a syndromic neurodevelopmental disorder known as Baraitser-Winter syndrome (OMIM #243310), significantly reduced NCRE activity (**Fig. 3e**). The same holds true when introducing point mutations in the TP53 motif of an NCRE interacting with *OAT*, a gene of which mutations are linked to gyrate atrophy of the choroid and retina (OMIM #258870), and in NCREs of *PAFAH1B1* and *ASH1L* (**Fig. 3e and Extended Data Fig. 7**,8), genes playing a role in lissencephaly (OMIM #607432) and intellectual disability (OMIM #617796), respectively. Previously reported variant prioritization scoring methods, including LINSIGHT^44^, GERP^45^, CADD^23^ and ncER^20^ score were not correlated with BRAIN-MAGNET cb scores across our NCRE atlas and the experimentally validated NCRE mutations (**Extended Data Fig. 6**d,e**, and Extended Data Fig. 7**-12) and were therefore less suited to identify these functional NCRE sequences and predict their mutation effects compared to BRAIN-MAGNET.

Together this indicates that BRAIN-MAGNET can predict NCRE activity based on DNA sequence composition and can be exploited to identify which nucleotides and motifs within NCREs can impact NCRE activity when mutated.

### Application of the NCRE atlas and BRAIN-MAGNET to prioritize genomic variants found in common disease helps to dissect GWAS loci

To test the utility of our NCRE atlas and BRAIN-MAGNET to prioritize genomic variants, we first applied them to genome-wide association studies (GWAS). Numerous GWAS have identified risk loci for neuropsychiatric disorders, the vast majority of which are located in the non-coding genome^2^. It has been tremendously challenging to decipher the mechanisms through which SNPs identified in GWAS might confer disease risks, as the majority of them are in linkage disequilibrium (LD) with adjacent SNPs. Thus, it remains unclear which of these genomic variants underscore the allele-specific biological effects causing the increased disease risks. Recently, MPRAs have been employed to fine-map GWAS loci and assess how the lead SNPs and adjacent SNPs in LD would affect the function of NCREs at single nucleotide resolution. In one such application, Guo et al. assessed 2,221 non-coding variants associated with 10 neuropsychiatric disorders, with an average of 5 SNPs per disease risk loci being tested^46^. Comparing the activity of matched risk versus reference allele pairs, they identified 892 SNPs that showed differential activity (daSNVs) in their MPRA depending on the genotype of the given SNP. As the BRAIN-MAGNET cb scores enable prediction of which nucleotides within a given NCRE have the highest likelihood of impacting the NCRE activity and daSNVs were already shown to affect NCRE activity in an allele-specific manner^46^, we hypothesised that daSNVs that overlapped with the NCREs measured in our study should have, on average, a higher BRAIN-MAGNET cb score than non-daSNVs. In agreement, we observed a significant difference in normalized cb score percentile between daSNVs and non- daSNVs (p=0.0374, t-test) (**Extended Data Fig. 13**a). In contrast, GERP, LINSIGHT, CADD and ncER scores did not significantly differ between the same daSNVs and non-daSNVs (**Extended Data Fig. 13**a). Since GWAS typically nominates a lead SNP, with other SNPs in LD, we next asked whether BRAIN-MAGNET cb scores could prioritize which SNP in a given GWAS locus would be more likely to be a daSNV (**Fig. 4a**). We therefore investigated GWAS-associated regions where multiple SNPs were previously functionally tested^46^ and for which multiple NCREs were present in our data set. One such locus on chromosome 6 contains in total 7 SNPs in LD which are located on 4 NCREs from our atlas, including rs401754 which was previously shown to be associated with an increased risk for schizophrenia (**Fig. 4b**). Amongst these 7 SNPs, rs200483 has relatively the highest normalized cb score compared to the other 6 SNPs (**Fig. 4c, Extended Data Fig. 13**b,c), and indeed rs200483 was previously identified as a daSNV^46^. Similarly, at a locus on chromosome 1 associated with major depression, comparing the relative normalized cb score amongst the 4 candidate SNPs in LD rightly pointed to rs301806 as the daSNV (**Fig. 4d,e and Extended Data Fig. 13**d,e). Of note is that other nucleotides within these NCREs, not overlapping with common SNPs, have higher cb scores, and alterations of those nucleotides might thus impact the activity of these sequences more severely than the daSNVs. The likely more severe effect of such genomic variants might not be compatible with the expected moderate risk effects of GWAS loci. Therefore, those nucleotides might not harbor common SNPs associated with common disease, but perhaps might harbor rare variants causing other related monogenic disorders. Together this indicates that comparing cb scores of candidate SNPs at GWAS loci can help to prioritize those SNPs that have the relatively highest likelihood to affect NCRE activity compared to the reference allele.

**Fig. 4:**
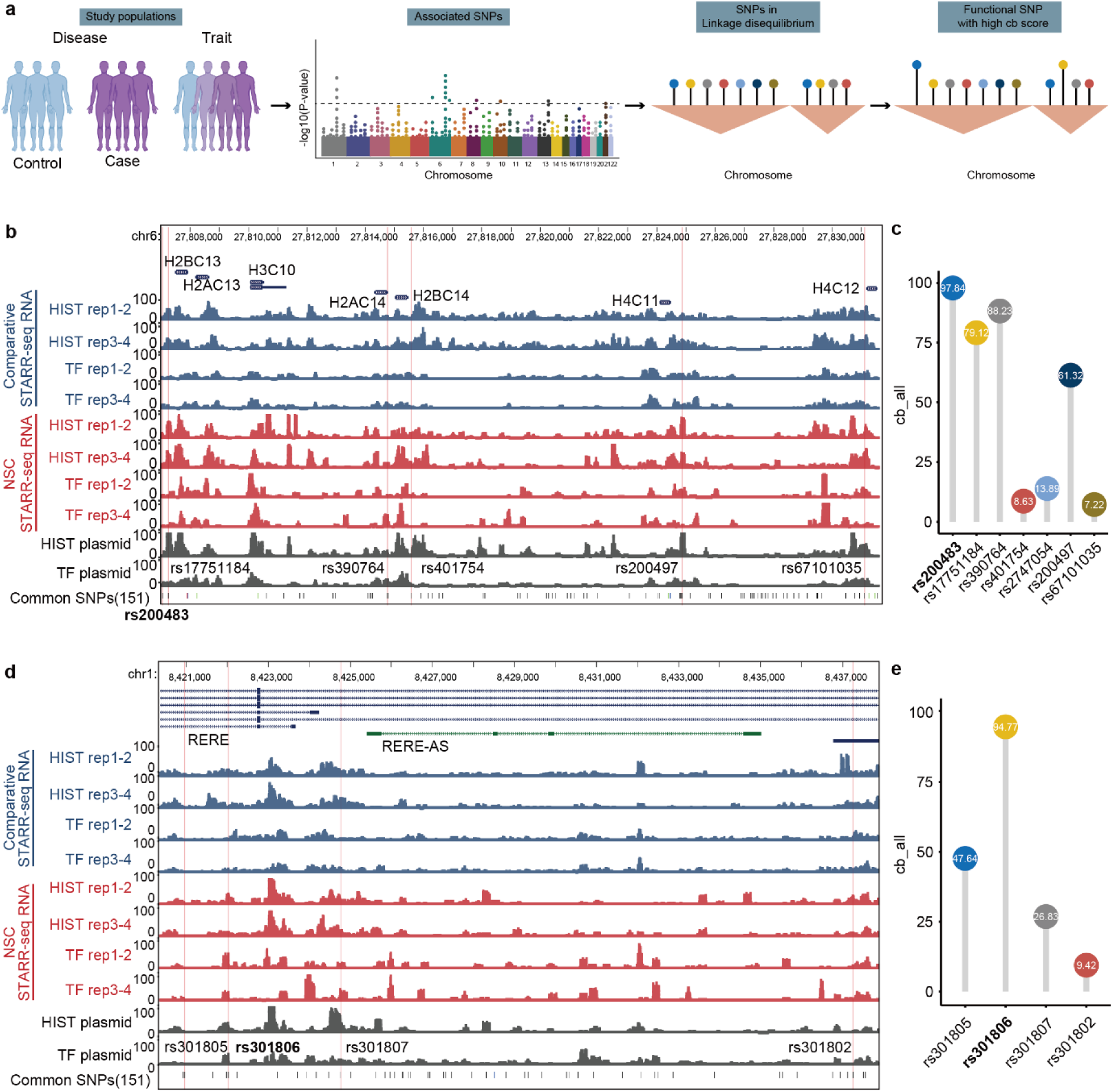
Utility of the NCRE atlas and BRAIN-MAGNET for prioritizing genomic variants found at GWAS associated regions a) Illustration showing how BRAIN-MAGNET can be used to prioritize functional SNPs amongst multiple SNPs in LD at GWAS loci. b) Genome browser view showing ChIP-STARR-seq RNA for ESCs (blue) and NSCs (red) and plasmid library sequencing tracks of a locus on chromosome 6 which GWAS studies have shown to be associated with increased risk for schizophrenia. 7 associated SNPs in LD which were also present on NCREs tested in our study are highlighted. c) Line plot showing the BRAIN-MAGNET cb scores (cb_all) for the 7 SNPs found at the schizophrenia risk locus from b). The rs200483 SNP has relatively the highest normalized cb score compared to the other 6 SNPs, and indeed rs200483 is one of the identified daSNVs from the MPRA studies by Guo et al^46^. BRAIN-MAGNET thus independently confirms previously experimentally tested data. d) As b), but now showing a GWAS locus associated with major depression on chromosome 1, with 4 GWAS associated SNPs in LD that overlap with NCREs in our study highlighted. e) As c), but now showing the BRAIN-MAGNET cb scores (cb_all) for the 4 SNPs found at the major depression risk locus from d). Again, the daSNV (rs301806) found by Guo et al^46^ has the relatively highest cb score amongst the 4 candidate SNPs.

### Application of the NCRE atlas and BRAIN-MAGNET to prioritize genomic variants found in rare disease identifies novel enhanceropathies

As the last proof of concept for the utility of the NCRE atlas and BRAIN-MAGNET, we investigated non-coding variants in WGS data from individuals with rare disease. To this end, we applied BRAIN-MAGNET to the data from the Genomics England 100,000 Genomes Project (GEL)^10^. If genetic variants at nucleotides with high cb scores would contribute to unexplained neurodevelopmental disorder phenotypes, we should observe an increased burden of *de novo* variants overlapping such nucleotides in genetically unexplained patients with neurodevelopmental disorders. To investigate this, we assessed 4,415 individuals from the unexplained neurology and neurodevelopmental cohort (NDD) of the v18 GEL data release for which 13,276 *de novo* variants overlapped with our NSC NCREs. Indeed, at population level, we observe a tendency towards an increased number of *de novo* variants at nucleotides with higher cb scores in NCREs from higher NSC ChIP-STARR-seq activity groups (**Extended Data Fig. 14**a), with similar findings for the NCREs identified by comparative ChIP-STARR-seq (**Extended Data Fig. 14**a). Likewise, we found a similar trend when investigating 4,558 genomes from individuals with a variety of other rare disease categories (**Extended Data Fig. 14**b).

We next screened the GEL data searching for variants that were overlapping with 20 bp fragments centred on the high confidence motifs found in the most active (category 4 and 5) NSC NCREs for which BRAIN-MAGNET predicts a major impact when disturbed. Following additional filtering steps, we identified 705 rare variants (each present ≤10 times in GEL) in 824 individuals with unexplained NDDs that were located within 20 bp centred on 440 motifs from NCREs that are linked to known OMIM genes (**Fig. 5a**). We randomly chose 4 heterozygous variants for functional validation which were absent in gnomAD v4, and found that three out of four variants significantly affected NCRE activity (**Fig. 5b**). The first heterozygous variant was in an NCRE linked to *GRIA4* (gene linked to autosomal dominant neurodevelopmental disorder with or without seizures and gait abnormalities, OMIM #617864), found in an individual with a complex neurological phenotype including ataxia and spasticity, and did not show a significant difference (**Fig. 5b and Extended Data Fig. 15**a). The second heterozygous variant that was identified in two unrelated individuals affected by epilepsy and ataxia and which is located in an NCRE linked to *KPTN* (gene linked to autosomal recessive intellectual developmental disorder 41, OMIM #615637) increased NCRE activity (**Fig. 5b and Extended Data Fig. 15**b). This indicates that genomic variants altering nucleotides with high cb scores do not necessarily only diminish NCRE activity but could also cause gain-of-function. The third heterozygous variant in an NCRE linked to *MN1* (gene linked to autosomal dominant CEBALID syndrome, OMIM #618774, that presents with dysmorphic features, intellectual disability, and brain malformations), significantly reduced the NCRE activity (**Fig. 5b and Extended Data Fig. 15**c). This variant was found in an unexplained individual reported to have a complicated epilepsy phenotype with amongst other features abnormalities of the cerebral cortex possibly presenting a (partial) phenotypic match. Lastly, the fourth heterozygous variant disrupts a ZFP42/YY1 binding motif in an NCRE linked by HiC to *RAB7A* and significantly reduced NCRE activity (**Fig. 5b,c**). This chr3:128681129 T>C (GRCh38/hg38) variant was identified in a genetically unexplained female clinically diagnosed with Charcot-Marie-Tooth disease and Sanger sequencing confirmed that her similarly affected mother carried the same variant. Interestingly, *RAB7A* is linked to an autosomal dominant type of Charcot-Marie-Tooth disease (type 2B, OMIM #600882)^47^ and is a dosage sensitive gene, indicating a phenotypic match. To further investigate how NCRE dysfunction might be implicated in this disorder, we first performed CRISPRi experiments targeting the *RAB7A* NCRE which resulted in significantly diminished *RAB7A* expression, confirming that *RAB7A* is under control of this NCRE (**Fig. 5d**). We next performed transgenic reporter assays in developing zebrafish larvae with wild type and patient-specific mutant *RAB7A* NCREs, both episomally and upon transposase-mediated NCRE integration. Under both conditions, and compared to the wild type, the patient-specific mutant *RAB7A* NCRE showed reduced expression in the central nervous system (CNS) 24 hours post fertilization (hpf). Also, a larger proportion of larvae with the patient-specific mutant NCRE showed bright, non-specific expression at aberrant locations not seen with the wild type *RAB7A* NCRE (**Fig. 5e, Extended Data Fig. 14**c,d**,e**). Together, this indicates that the patient-specific *RAB7A* NCRE variant leads to reduced expression in the CNS and an altered spatiotemporal NCRE activity *in vivo*, indicating NCRE dysfunction. Hence, it is currently considered that this non-coding variant is causative for the disease phenotype and represents a novel enhanceropathy. Clinical follow-up has been initiated for these individuals.

**Fig. 5:**
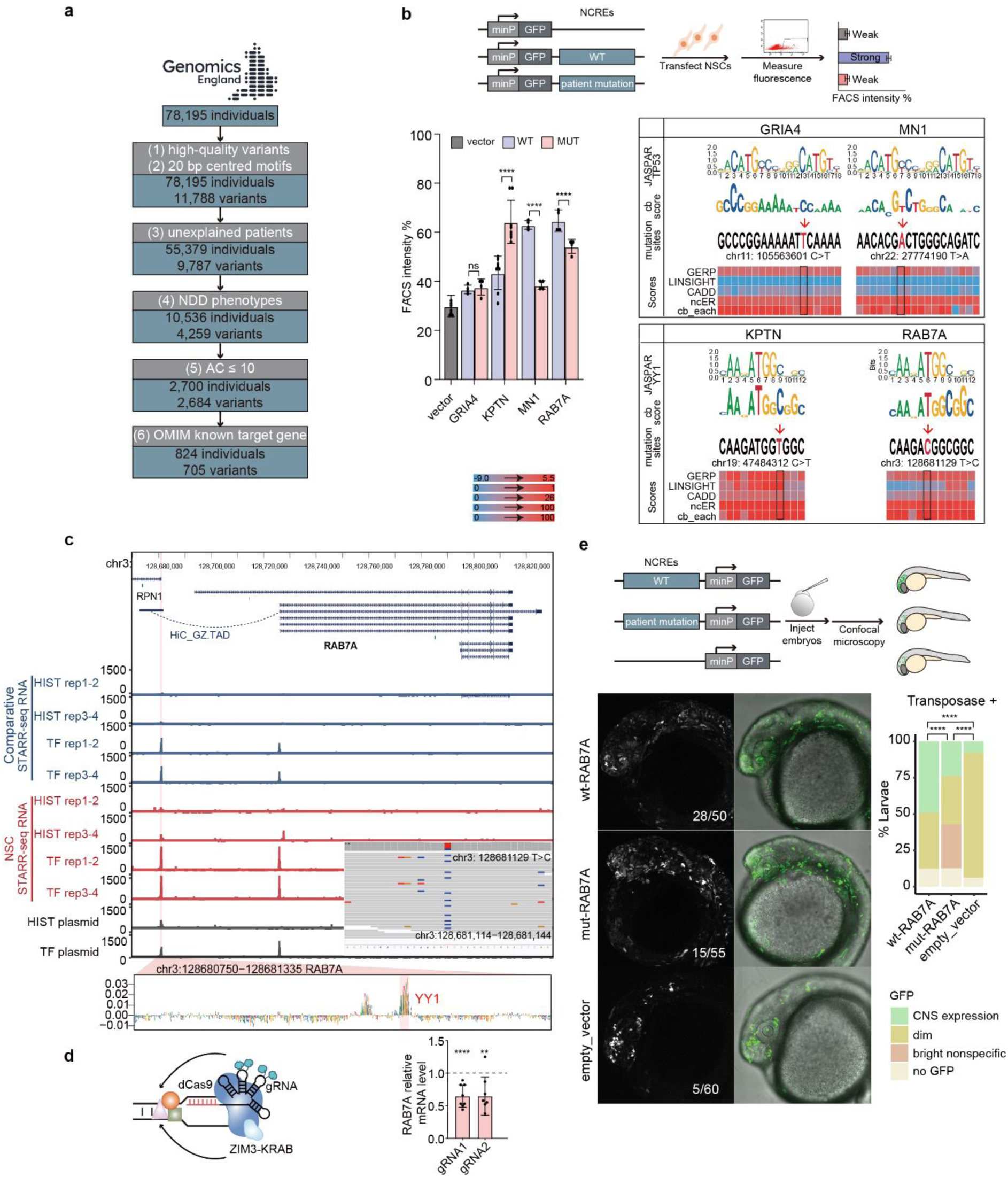
Utility of the NCRE atlas and BRAIN-MAGNET for prioritizing genomic variants found in rare disease a) Flow diagram showing the filtering of the Genomics England 100,000 Genomes Project (GEL) data for rare variants overlapping with 20 bp fragments centred on the high confidence motifs found in the most active (category 4 and 5) NSC NCREs for which BRAIN-MAGNET predicts a major impact when disturbed. b) Functional validation of 4 rare heterozygous variants identified in GEL overlapping with NCREs for which BRAIN-MAGNET predicts an effect on NCRE function. Upper diagram shows experimental validation strategy. Bar plot showing the percentage of GFP+ cells in NSCs upon cell transfection experiments with reporter plasmids containing either empty reporter plasmid control (grey), wild type NCREs (blue) or the same NCREs with patient mutations found in GEL (pink). The motifs predicted from JASPAR, the BRAIN-MAGNET cb score, the patient mutation and other variant prioritization outputs displayed as heatmap are indicated in the right panel. NCREs are indicated with the name of the presumed target gene. Plotted on the left is the percentage of GFP+ in cells co-transfected with an mCherry expressing plasmid, to correct for transfection efficiency. Bars show the average from two independent experiments, with each NCRE tested in duplicate. Error bars represent standard deviation. * p < 0.05; ** p < 0.01; *** p < 0.001; **** p < 0.0001 (one-way ANOVA test followed by multiple comparison test (Fisher’s LSD test)). c) Genome browser view of the *RAB7A* locus, a gene implicated in an autosomal dominant form of Charcot-Marie-Tooth disease, showing ChIP-STARR-seq RNA for ESCs (blue) and NSCs (red) tracks and plasmid library sequencing results. This identified a *RAB7A* NCRE (highlighted) located ∼45kb upstream of *RAB7A* that interacts with the main *RAB7A* transcriptional start site as confirmed by HiC. Zoom-in shows BRAIN-MAGNET cb scores and predicted functional motifs in the *RAB7A* NCRE. The inserted IGV browser view shows whole genome sequencing data from GEL identifying a heterozygous rare variant that affects the NCRE predicted functional YY1 motif. This variant significantly affects NCRE function (as tested in b), and was detected in a genetically unexplained individual clinically affected by Charcot-Marie-Tooth disease, possibly indicating a novel enhanceropathy. d) CRISPRi experiment in HEK293T cells. Schematic on the left shows the targeting by gRNAs of the *RAB7A* NCRE by ZIM3-KRAB-dCas9. Right diagram shows qRT-PCR detecting *RAB7A* expression (normalized to TBP as housekeeping control) upon targeting with 2 different gRNA combinations. Wild type expression levels for RAB7A are depicted using the dashed line. Error bars represent standard deviation. * p < 0.05; ** p < 0.01; *** p < 0.001; **** p < 0.0001 (one- way ANOVA test followed by multiple comparison test (Fisher’s LSD test). e) NCRE *in vivo* reporter assay in zebrafish. Upper diagram illustrates experimental approach. Left lower panel shows representative confocal microscopy of transgenic zebrafish larvae (with transposase) at 24 hpf in lateral view. Numbers indicated in the images present the GFP positive zebrafish larvae per condition. Right lower panel shows quantification (**** p < 0.0001, P_value (Chisq test)). Compared to the wild type, the patient-specific mutant *RAB7A* NCRE showed reduced expression in the central nervous system. Also, a larger proportion of larvae with the patient-specific mutant NCRE showed bright, non-specific expression at aberrant locations not seen with the wild type *RAB7A* NCRE. Together, this confirms that the patient- specific variant *RAB7A* NCR leads to an altered *in vivo* spatiotemporal NCRE expression pattern indicating NCRE dysfunction.

Finally, we screened through three independent WGS sources. First, focussing on a set of 249 trio WGS of unsolved rare disorder patients and solely on *de novo* variants overlapping NCREs with a cb score above the 95^th^ percentile, we identified 12 variants in NCREs which target genes could potentially provide a clinical match. We then screened for deletions overlapping the same set of NCREs in 3,971 singleton WGS of undiagnosed rare disease patients and compared HPO-terms of NCRE target genes and the individuals. This identified 5 heterozygous carriers harbouring deletions of NCREs which were regulating known disease genes causing overlapping phenotypes as observed in the patients but with an autosomal recessive mode of inheritance, likely representing carrier status. In addition, we identified three deletions of NCREs regulating genes with an autosomal dominant mode of inheritance, possibly representing a clinical phenotypic match. This included a heterozygous ∼5.7 kb deletion of an NCRE for *NEFL* (**Extended Data Fig. 14**f), a gene associated with various dominant forms of Charcot-Marie-Tooth disease^48^, found in a genetically undiagnosed adult with motor and sensory neuropathy providing a clinical phenotypic match. Ultimately, we applied BRAIN-MAGNET to clinical WGS data from our center analyzing a genetically unexplained patient clinically suspected of Mowat-Wilson syndrome and ruled out a non-coding variant in an intron of *ZEB2* as the likely disease cause (**Supplementary Note 3, Extended Data 16**). This shows how BRAIN-MAGNET can also be used to clinically eliminate potential candidate disease-causing non-coding variants.

Together, this indicates that the NCRE atlas presented herein, coupled with BRAIN-MAGNET, can facilitate the search for possibly disease-implicated rare variants identified by WGS in rare disease cohorts. Furthermore, BRAIN-MAGNET can predict the impact of such variants on functional NCRE activity and aid in their clinical interpretation (**Fig. 6**).

**Fig. 6:**
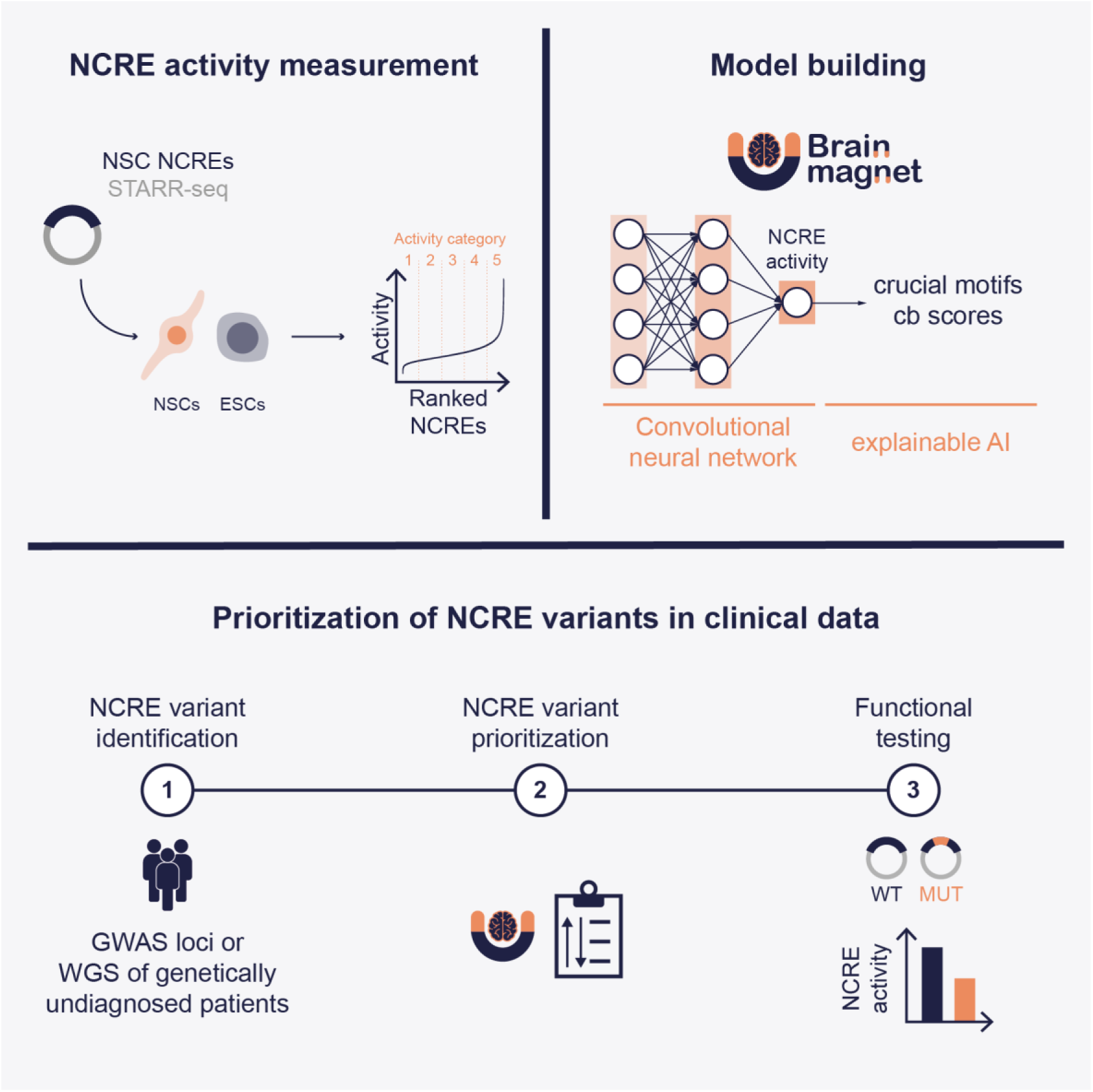
Graphical abstract summarizing BRAIN-MAGNET and its application to prioritize genetic variants in NCREs in rare and common disease

## Discussion

Here, we used ChIP-STARR-seq to build an extensive atlas of NCREs functionally active in NSCs. Besides informing on the biological mechanisms regulating gene expression in NSCs and on NCREs distinguishing sequence characteristics, we provide evidence that a CNN model trained on the experimental data allows to predict NCRE activity solely based on NCRE nucleotide composition, similar to what other recent MPRA studies have found^49–51^. BRAIN-MAGNET can be used to predict the functional effects of genomic variants overlapping with NCREs, and such predictions occur with high confidence as testified by our functional validation. We show how BRAIN-MAGNET can be used to interpret effects of common SNPs identified in GWAS, as well as rare variants identified by WGS in affected individuals with currently unexplained neurological and neurodevelopmental phenotypes. We thus anticipate that this resource might be useful to diminish so-called missing heritability of non-coding variants in the field of human genetics, by providing an additional option for variant prioritization based on BRAIN-MAGNET cb scores.

Our study has several limitations. First, MPRAs measure NCRE activity in an episomal context, outside of the natural chromatin environment, hence, results might not always reflect the endogenous NCRE activity. Notwithstanding this potential limitation, previous work has already extensively shown that multiple MPRA findings can be reproduced when altering NCREs at the endogenous locus^9,49,52^. To minimize potentially confounding effects, we generated ChIP-STARR-seq plasmid libraries from chromatin marked in NSCs with histone modifications associated with active NCREs. In addition, the episomal approach might also reveal insights in mechanisms that are alternatively difficult to study at the endogenous locus, such as the potential priming of NCREs in ESCs for later activation that we observe here when using comparative ChIP-STARR-seq. Second, results of MPRAs are obtained in a given cell type, and it remains to be investigated how cell type specific findings and rules identified by CNN models can be extrapolated to other cell types and tissues. Third, although we show the utility of this resource to prioritize non-coding genomic variants for functional effects on NCRE activity based on their BRAIN-MAGNET cb scores, it remains challenging to draw final conclusions regarding pathogenicity of such variants and causality in causing rare disease. One of the largest conceptual challenges in this regard is the fact that the clinical phenotype of an NCRE-disrupting variant does not necessarily have to equal the phenotype of a protein coding pathogenic variant affecting the gene that the NCRE is regulating. This is illustrated by tissue-specific effects of mutations in the *PTF1A* enhancer, where such NCRE mutation only causes part of the phenotype compared to that of the *PTF1A* disease gene^53^. Hence, when finding a variant that strongly impacts NCRE function, its associated phenotype might be similar to the known phenotype of the target gene, such as is likely the case of the *RAB7A* example presented here, but might also cause phenotypes that represent only “partial known syndromes”, or novel phenotypes which might complicate the clinical interpretation of such variants. Future studies will be required to more clearly dissect correlations between cell type specific effects of NCREs and phenotypes that NCRE dysfunction might cause. The NCRE atlas presented herein and BRAIN-MAGNET will form a solid starting point for such studies.

## Acknowledgements

We thank all members of the Barakat lab, the Clinical Genetics Discovery Unit, Innovation Team and Whole Genome Sequencing Implementation and Research Task Force at Erasmus MC for helpful discussions. Shimriet Zeidler (Erasmus MC) is acknowledged for sharing clinical information on the *ZEB2* case. Some of the authors are members of the European Reference Network on Rare Congenital Malformations and Rare Intellectual Disability ERN-ITHACA. RD was supported by a China Scholarship Council (CSC) PhD Fellowship (201906300026 to RD) for her PhD studies at the Erasmus Medical Center, Rotterdam, The Netherlands. KL was supported by a ZonMw PSIDER Doorbraken grant (grant 10250042110005), a Brain and Behavior Research Foundation Young Investigator award (grant 30787) and a NWO Veni grant (grant 501100003246). JP was supported by the Clinician Scientist program “PRECISE.net” funded by the Else Kröner-Fresenius-Stiftung and by the intramural TÜFF program (3049-0-0). TBH was supported by the Deutsche Forschungsgemeinschaft (DFG, German Research Foundation – 418081722, 433158657), and the European Commission (Recon4IMD - GAP-101080997). GR was supported by the ZonMw Veni grant 1936320. Part of this research was made possible through access to the data and findings generated by the 100,000 Genomes Project. The 100,000 Genomes Project is managed by Genomics England Limited (a wholly owned company of the Department of Health and Social Care). The 100,000 Genomes Project is funded by the National Institute for Health Research and NHS England. The Wellcome Trust, Cancer Research UK, and the Medical Research Council have also funded research infrastructure. The 100,000 Genomes Project uses data provided by patients and collected by the National Health Service as part of their care and support. Some of the analysis involved external data generated by the ENCODE and Roadmap projects, that received funding from the National Institutes of Health (NIH) (grants U01ES017166, U54HG004570, U41HG006992 and U01ES017155). The Barakat lab was supported by the Netherlands Organisation for Scientific Research (ZonMw Veni, grant 91617021; ZonMw Vidi, grant 09150172110002), a NARSAD Young Investigator Grant from the Brain & Behavior Research Foundation, an Erasmus MC Fellowship 2017, and Erasmus MC Human Disease Model Award 2018, and acknowledges other ongoing support for rare disease research from Stichting 12q, EpilepsieNL, CURE Epilepsy, Spastic Paraplegia Foundation, Inc and the Sophia Research Foundation (Stichting Sophia Kinderziekenhuis Fonds). Funding bodies did not have any influence on study design, results, and data interpretation or final manuscript.

## Data availability

Data generated herein, including ChIP-STARR-seq, RNA-seq, ChIP-seq, ATAC-seq and BRAIN-MAGNET can be visualized using the following UCSC browser session: https://genome.ucsc.edu/s/BarakatLab/BrainMagnet_NSC_ESC_cb_scoreshg38. Genome sequencing data from the 100,000 Genomes Project and limited phenotype information are available in the National Genomic Research Library from the 100,000 Genomes Project for which researchers can apply for access at Genomics England. Due to privacy regulations and given consent under which the Erasmus MC and IMGAG Tübingen data base patients were recruited, raw patient genomic sequencing data cannot be made available. Codes used for data analysis are available via GitHub: https://github.com/ruizhideng/NSC-ChIP-STARR-seq_paper.

## Declarations

All authors declare no conflict of interest.

## Author contribution

RD performed all computational and genomic analysis, with help of SY, RS, MM, JP, MS, TBH, GVR and EM. EP performed all multi-omics experiments. AN, KL, EMS and LES performed validation experiments. MJP provided clinical data. WvIJ and Genomics England Research Consortium performed next generation sequencing and genome sequencing. TSB conceived and supervised the work. RD, EP and TSB wrote the manuscript with input from all authors. All authors approved the final version of the manuscript.

## Supplementary Note 1

### Adapted ChIP-STARR-seq approach

Compared to our previous application of ChIP-STARR-seq in human embryonic stem cells (ESCs) ^9^ that focussed on NCREs enriched for the TFs NANOG, OCT4, and the histone modifications H3K4me1 and H3K27ac (herein referred to as ESC ChIP-STARR-seq), we performed a number of updates on the experimental procedures, aiming to reduce experimental noise, increasing reproducibility and reducing experimental complexity and costs (**Extended Data Fig. 1**a).

First, to enrich for genomic regions with putative NCRE function in NSCs, we performed ChIP in duplicates for the TFs SOX2 and YY1, that are important in NSCs^13,14^, and the histone modifications H3K27ac and H3K4me1, which are associated with putative NCREs^15^. ChIP-qPCR showed an enrichment at known NCREs surrounding the *FGFR1*, *PAX6*, *SOX2* and *NES* loci (**Extended Data Fig. 1**b). Sequencing of the ChIP material (ChIP-seq) showed a good correlation between replicates (Pearson correlation coefficient > 0.88) (**Extended Data Fig. 1**c,d). Next, using the same ChIP-DNA, ChIP-STARR-seq plasmid libraries were generated from both ChIP replicates and pooled. To further decrease the number of transfections and simultaneously increase the number of individual plasmids covering each putative NCRE, theoretically reducing experimental noise, we pooled together the plasmid libraries derived from ChIP for the histone modifications H3K27ac and H3K4me1, and for the TFs SOX2 and YY1. Hereafter, we refer to these combined plasmid libraries as HIST and TF libraries, respectively, and the application of these libraries for ChIP-STARR-seq in NSCs or ESCs as NSC ChIP-STARR-seq or comparative ChIP-STARR-seq, respectively. Visual inspection of sequencing results to assess whether the pooled plasmid libraries are a good representation of the initial ChIP material, showed that peaks from ChIP-seq experiments were still recognisable in the sequencing tracks of the pooled plasmids (**Extended Data Fig. 1**e), and analysis of normalized sequencing reads confirmed a high correlation between initial ChIP-seq peaks and corresponding pooled plasmid libraries (**Extended Data Fig. 1**c,d). On average, 93.04% of HIST ChIP-seq peaks and 96.86% of TF ChIP-seq peaks were covered by more than 5 different plasmids and 46.91% of HIST peaks and 67.54% of TF peaks by more than 20 plasmids, with a mean of 27 and 43 distinct plasmids per HIST and TF ChIP-seq peak, respectively. Together this indicates an increased complexity of the generated plasmid libraries compared to our earlier study using ChIP-STARR-seq in ESCs^9^.

Second, to overcome the variability between samples that we previously observed in ESC ChIP-STARR-seq^9^, we implemented the following changes in the experimental strategy. We performed 5 independent transfections per plasmid pool, each starting with ∼3 million NSCs seeded 48 hours before the transfection (or ∼6 million ESCs transfected in suspension, see **Methods**). 24 hours post-transfection, we collected and pooled together approximately 30 million cells from these 5 independent transfections (**Extended Data Fig. 1**a). FACS analysis showed that 26.5% and 30% of NSCs transfected with the HIST or TF libraries, respectively, were GFP positive (**Extended Data Fig. 1**f).

Third, RNA was isolated from these pooled, unsorted cells and subsequently split and treated as two replicates, which were subjected to DNase treatment and cDNA synthesis. To limit PCR artefacts, the first PCR amplification step which suppresses residual plasmid contamination by the use of primers spanning a synthetic intron in the STARR-seq plasmid, was performed in 10 parallel reactions for each of the two replicates. These 10 PCRs per replicate were then pooled, purified and further amplified with the index primers required for sequencing, using 5 independent PCR reactions and two independent index primers for each replicate. Thus, we sequenced a total of four replicates of STARR-seq RNA for each pooled HIST or TF library, with an average of 20 million reads per sample (**Extended Data Fig. 1**a). As the two technical replicates amplified with independent index primers showed a high correlation (Pearson correlation coefficient >0.96) (**Extended Data Fig. 1**g-j), they were merged for further downstream data analysis.

Together this indicates that our revised experimental strategy generated highly complex plasmid libraries capturing genomic regions marked by H3K27ac, H3K4me1 or bound by SOX2 and YY1 in NSCs, which upon transfection and RNA sequencing results in highly correlated replicate data.

## Supplementary Note 2

### Differential NCRE activity analysis in NSCs and ESCs

To identify differences in NCRE activity between the two cell types, we next performed a differential NCRE activity analysis^16^, comparing NCREs captured in the same NSC-derived plasmid library with high activity in the one cell type and low activity in the other, and vice versa (**Extended Data Fig. 4**a). To this end, we first selected those NCREs that ranked in the top-10% of NSC ChIP-STARR-seq activity in NSCs and in the bottom-90% of comparative ChIP-STARR-seq in ESCs, and those NCREs that ranked in the top-10% of comparative ChIP-STARR-seq activity in ESCs and the bottom-90% in NSC ChIP-STARR-seq. With these settings, we identified 6,850 out of 14,818 NCREs (46.2%) overlapping between the top-10% of NSCs and ESCs (referred to as common-high) (**Extended Data Fig. 4**b), possibly explaining part of the overlap between TF motifs and TEs in NSC and ESC active NCREs (**Fig. 1j and Extended Data Fig. 3**c), and 7,968 out of 14,818 NCREs (53.8%) per cell type that were either NSC- or ESC-high. Visual inspection of the tracks surrounding the *CHD2* locus shows both common-high NCREs and differentially active NCREs (**Extended Data Fig. 4**c). Investigating the distance distribution of these regions from the TSS, we observed that part of the NSC-high NCREs is located in closer proximity to TSSs, while common and ESC- high NCREs were spatially distributed in a more similar manner (**Extended Data Fig. 4**d). A deeper investigation revealed that all the three classes or NCREs are enriched mainly at intronic (NSC-high: 46.4%; ESC-high: 55.2%; common-high: 53.7%) and intergenic (NSC- high: 29.6%; ESC-high: 36.7%; common-high: 35.9%) regions with an increase at promoter-TSS regions for the NSC-high ones (NSC-high: 17.1%; ESC-high: 4.4%; common-high: 6.3%), in agreement with their tendency to be located in closer proximity to the TSS (**Extended Data Fig. 4**e). Next, we asked whether cell-type specific highly active NCREs were also more susceptible to LoF. As observed previously (**Extended Data Fig. 2**f,3b), the majority of the investigated regions are intolerant to LoF, with NSC-high NCREs showing a larger enrichment amongst intolerant regions, possibly indicating that alterations of these regions might be negatively selected in the human population or might be implicated in disease (**Extended Data Fig. 4**f). Investigating the pLI of the genes linked to these NCREs groups, we did not find a significant difference (**Extended Data Fig. 4**g).

Next, we aimed at understanding whether these NCREs might regulate cell-type specific gene expression. We observed that NSC-high NCREs are linked to genes expressed at a significantly higher level in NSCs but also in ESCs (**Extended Data Fig. 4**h), whereas genes linked to ESC-high and common-high NCREs did not show major expression differences. Gene ontology analysis identified for all three categories of NSC-high, ESC-high and common-high NCREs terms related to transcriptional regulation (**Extended Data Fig. 4**i). Common- high NCREs were also enriched for terms related to nervous system development and generation of neurons and axons. Like previously seen for all NCREs active upon comparative ChIP-STARR-seq in ESCs, also ESC-high NCREs were enriched for nervous system development and generation of neurons, rather than other expected terms for pluripotent stem cells, despite that these genes are not highly expressed in ESCs. This observation further provides support for the hypothesis that these ESC-high NCREs, despite showing episomal activity in comparative ChIP-STARR-seq in ESCs when using plasmid libraries obtained from NSCs, might not be active at the endogenous chromatin in ESCs. Rather, these ESC-high NCREs might be primed for future activation at the endogenous locus, and this priming might not be recapitulated in an episomal plasmid-based system.

To further investigate this hypothesis, we assessed various chromatin profiling data at the common and cell-type specific highly active NCREs. As expected, since all the investigated putative NCREs are derived from H3K27ac, H3K4me1, YY1 and SOX2 ChIP in NSCs, all the common and differentially active NCREs (ESC-high, NSC-high) show central enrichment of these chromatin marks across NSC ChIP-seq data (**Extended Data Fig. 4**j,k). To further characterize the 7,968 NSC-high NCREs at their endogenous locus, we retrieved H3K4me3 and H3K27me3 from previously published data of H9-derived NSCs^54^ and performed ATAC-seq in NSCs to assess chromatin accessibility. We observed that the NSC-high NCREs are enriched for accessible chromatin, as expected given the chromatin immunoprecipitation in this cell type, and a strong enrichment of the active chromatin mark H3K4me3 (**Extended Data Fig. 4**j,k) overlapping the enrichment of YY1 and H3K27ac. H3K4me3, as well as YY1, is highly enriched at promoters^55^, suggesting this fraction of loci might partially include the 17% of NSC-high NCREs found in proximity to promoters (as previously discussed, the STARR-seq design does not allow to test promoter activity and thus despite being located in proximity to promoters these sequences are NCREs). Compatible with the immunoprecipitation of markers associated with NCRE activity used to identify ChIP-STARR-seq NCREs, we did not see enrichment of the inactive chromatin mark H3K27me3 at these NCREs.

To investigate the hypothesis that ESC-high NCREs are not enriched in active marks at the endogenous chromatin loci, we collected H9 ESC epigenome data from publicly available sources and from our previously generated H3K27ac, H3K4me1, OCT4 and NANOG ChIP-seq^9^. Furthermore, we generated ChIP-seq data for YY1 and SOX2 in ESCs. ESC-high NCREs, even though located in accessible chromatin in ESCs, show no enrichment of the active chromatin marks H3K27ac and H3K4me1 in ESCs (**Extended Data Fig. 4**j,k). The majority of the ESC-high NCREs from comparative ChIP-STARR-seq in ESCs show a very mild enrichment of SOX2 and NANOG binding, but interestingly, a subset of the ESC-high NCREs (n=1,100 out of the 7,968 ESC-high NCREs) had an accumulation of the histone marks H3K4me2 and H3K4me3, with especially the latter mark showing a bi-modal distribution with a valley over the NCRE centre. Primed NCREs are indeed characterized by methylation of H3K4 and no acetylation of H3K27^37^. Therefore, this observation is compatible with the hypothesis that these NCREs, despite being highly active in the episomal setting, are not active at the endogenous locus but primed for activation at a later stage. Further pointing to this scenario, we noticed that ESC-high NCREs primed with H3K4 methylation in ESCs, gain H3K27 acetylation at the NSC stage, although to a lower level compared to NSC-high NCREs, compatible with the lower activity of these regions in NSCs (**Extended Data Fig. 4**j,k). Interestingly, these regions maintain H3K4me3 also at later stages of differentiation, in neurons and in astrocytes, where they also display H3K27ac. The same scenario described for ESC-high NCREs applies to common-high NCREs, for which we could observe a fraction of regions (n=1,200 out of 6,850 common-high) with H3K4me2/3 accumulation and no H3K27ac in ESCs, but with H3K27ac in NSCs. H3K4me3 regions have been previously reported to be located mainly at the TSS^55^. In line with this, we observed that the majority of the ESC-high and common-high NCREs marked by H3K4me3 were located in proximity of a TSS, within 5kb down- or upstream (**Extended Data Fig. 4**l), but as outlined as the STARR-seq design does not allow to capture promoter activity these are thus NCREs in proximity of the TSS. Moreover, when investigating the enrichment of TF binding motifs, the TATA-box was among the most enriched motifs in the H3K4me2/3 positive subset of both the ESC-high and common-high NCREs (**Extended Data Fig. 4**m). Further supporting the hypothesis that this subset of regions is primed for activation at a later stage, we noticed that the majority of them (>75%) in NSCs belong to the NCRE activity categories 4 and 5 (**Extended Data Fig. 4**n).

Analysing further the TF motifs enriched in these regions, we noticed that the most enriched in the common-high subset positive for H3K4me2/3 in ESCs were YY1, p73 and p53, possibly explaining their activity in NSCs where these regions are marked by H3K27ac. We also identified the OCT4 and OCT4-SOX2-TCF-NANOG^56^ motifs at this subset of ESC-high NCREs, possibly contributing to the episomal activity of these NCREs. Interestingly, the repressor protein ZNF281 was enriched at a small subset of ESC-high, H3K4me2/3 positive NCREs (**Extended Data Fig. 4**n). ZNF281 is a known repressor protein expressed at high levels in ESCs but downregulated upon neural differentiation^57^, and it will therefore be interesting to investigate whether ZNF281 or related factors might contribute to explain silencing of some of the ESC-high NCREs at the endogenous chromatin.

## Supplementary Note 3

### BRAIN-MAGNET eliminates a clinically suspected candidate disease causing non-coding variant

We applied the NCRE atlas to a patient from our clinic, which was clinically suspected of Mowat-Wilson syndrome, a severe neurodevelopmental disorder linked to dysfunction of the *ZEB2* gene. Previous extensive genetic investigations, including trio whole exome and genome sequencing did not reveal a protein coding mutation in *ZEB2*, or any other likely disease-causing variant, but identified a heterozygous chr2:144469837C>T variant in intron 2 of *ZEB2*. This variant overlapped with a highly active NCRE from NSCs (category 5), was absent in gnomAD v4, had a CADD score of 21.0, an ncER score of 99.6 and was predicted by the RegulomeDB v.2^58^ to contain a TF binding motif (**Extended Data Fig. 14**a). According to LINSIGHT^44^, an algorithm that aims to predict which non-coding variants are likely to have deleterious fitness effects, the chr2:144469837C>T variant was predicted to be likely pathogenic (score 0.98). In contrast, according to BRAIN-MAGNET, this nucleotide had a low cb score (65.7^th^ percentile within the NCRE (cb_each); 52.4^th^ percentile within all NCREs (cb_all)), while a different ONECUT1 TF motif located 36 nucleotides upstream is predicted to mostly impact on activity of this NCRE. To investigate this functionally, we cloned the NCRE in the STARR-seq plasmid and introduced by site directed mutagenesis either the variant identified in the patient or a 27 bp deletion centred on the ONECUT1 motif. Upon transfection in NSCs, the ONECUT1 deletion completely abolished the NCRE activity, while the patient variant did not majorly affect NCRE activity, in agreement with the BRAIN-MAGNET predictions (**Extended Data Fig. 14**b). In addition, clinical RNA-seq of patient derived fibroblasts did not reveal any *ZEB2* expression changes and a diagnostic Episignature did not reveal evidence of a Mowat-Wilson syndrome specific DNA-methylation pattern (data not shown). Even though we cannot completely exclude that the intronic *ZEB2* variant could have an effect in other cell types, based on the current functional testing the variant was clinically classified as a variant of unknown significance which was considered unlikely to contribute to the phenotype which remains unexplained. This illustrates how BRAIN-MAGNET can be used to clinically eliminate potential candidate causing non-coding variants identified during filtering of WGS data sets.

## Experimental procedures

### Cell culture

H9-derived human Neural Stem Cells (NSC, Gibco) were cultured as previously described^59^. Briefly, cells were seeded onto Geltrex (Gibco) coated plates and cultured in KnockOut DMEM/F12 (Gibco) supplemented with 2 mM L-Glutamine (Gibco), 2% StemPro neural supplement (Gibco), 20 ng/ml EGF (Peprotech), 20 ng/ml b-FGF (Peprotech), 100 U/ml penicillin and 100 µg/ml streptomycin. Cells resulted negative at mycoplasma testing.

H9 ESCs were cultured as previously described^59^ on Matrigel (Corning) coated plates in MTeSR-1 medium (STEMCELL Technologies).

### Chromatin immunoprecipitation

For chromatin immunoprecipitation, 5x10^7^ NSC were harvested in 9 mL of PBS and dual cross-linked first with 2mM Di(N-succinimidyl) glutarate (Sigma) for 45 min followed by 10 minutes 1.1% buffered formaldehyde (50mM Hepes-KOH pH7.6, 100mM NaCl, 1mM EDTA pH8.0, 0.5mM EGTA pH8.0, 11% formaldehyde). Formaldehyde was then quenched with 125 mM glycine (Sigma) for 10 min. Cross-linked cell pellets were snap-frozen and stored at - 80°C.

ChIP was performed as described^9^. Briefly, cell nuclei were sonicated at 4°C with Diagenode Bioruptor for 45 cycles (30 sec on, 30 sec off), prior to overnight incubation with antibody-coupled beads. The antibodies used were rabbit-anti-H3K4me1 (ab8895, Abcam), rabbit-anti- H3K27ac (ab4729, Abcam), rabbit-anti-YY1 (ab109237, Abcam) and goat-anti-SOX2 (AF2018, R&D systems). Immunoprecipitated chromatin and total input control were de-cross- linked in a final volume of 100 µL, by addition of 3 µL of 10% SDS and 10 µL proteinase K (10 mg/ml, Roche) and 5µl RNase (11119915001, 500 ng/µl, Roche) and incubation overnight at 65°C at 1400 RPM on a shaking thermomixer block. Following Phenol:Chloroform:IAA (Sigma) extraction and ethanol precipitation, the concentration of ChIP-DNA was determined by Qubit (Invitrogen) and the sonication was assessed by gel-electrophoresis of total input DNA (target fragment size between 200 and 600 bp).

### ChIP-qPCR

For qPCR, 10µl of ChIP material and total input material were diluted 6 times. 2 µL of DNA were used per qPCR reaction, using iTaq universal SYBR Green Supermix in a CFX96RTS thermal cycler (Bio-Rad), following manufacturer’s instructions. Fold enrichment was determined following the ΔΔct method. All data shown are averages of at least 2 biological replicates and 2 technical replicates.

### ChIP-seq, ChIP-STARR-seq plasmid library preparation

ChIP-seq and ChIP-STARR-seq plasmid libraries were generated as described^9^. Briefly, 10 ng of ChIP DNA were end-repaired, dA tailed and adaptor ligated using NEBNext ChIP-seq library preparation kit (E6240, NEB), according to manufacturer’s instructions. Adaptor ligated DNA was eluted into 32 µL of 0.1xTE, 25 µL were used for ChIP-seq library preparation and 5 µl were processed for ChIP-STARR-seq plasmid libraries preparation.

For ChIP-seq library preparation, purified adaptor ligated DNA was PCR amplified with Illumina index primers (E7335 and E7500, NEB) according to manufacturer’s instructions. ChIP-seq libraries were assessed on an Agilent Tapestation. All sequencing occurred on an Illumina HiSeq 2500 platform, using 50bp single-end sequencing.

For details about the plasmid library preparation see Barakat *et al*^9^. Briefly, 5 µL of purified adaptor ligated DNA for each replicate were separately amplified with primers 147 STARRseq libr FW and 148 STARRseq libr RV^11^, and used in a Gibson assembly reaction with an AgeI- SalI digested STARR-seq plasmid and a home-made Gibson reaction buffer (100mM Tris-HCl, 10mM MgCl2, 0.2 mM dNTP, 0.5U Phusion DNA polymerase (NEB), 0.16U 50 T5 exonuclease (NEB)). Gibson reactions for the ChIP replicates were pooled together, purified by ethanol precipitation and used for electroporation into electrocompetent MegaX DH10b *E. coli* bacteria (C640003, Invitrogen), according to manufacturer’s instructions. A total of 5 electroporation reactions per library were performed with 2 µl of DNA each. After recovery, bacteria from the 5 reactions were pooled together and 100 µL of a 1:100 and 1:10000 dilution was plated on Ampicillin containing Agar plates to enable estimation of the number of transformants. The remaining 5 mL of bacteria culture were incubated in 1 L of LB-media supplemented with Ampicillin over-night, and the plasmid DNA was isolated using a Maxiprep kit (Macherey-Nagel) according to manufacturer’s instructions.

### Transfection of plasmid libraries

For each transfection in NSC ChIP-STARR-seq, approximately 3 million NSC were seeded in a 10 cm dish and 48 hours later transfected with 6.8 µg of plasmid library DNA and 27.5 µl Lipofectamine Stem Transfection Reagent (STEM00015, Invitrogen) following manufacturer’s instructions. In total, 5 dishes of transfected cells were used. For ESCs transfections in ESC ChIP-STARR-seq, approximately 6 million cells were seeded in a 10 cm dish and transfected in suspension with an identical transfection mix to NSCs in the presence of 5µM ROCK-inhibitor (Y27632, Millipore). After 24h of transfection, cells were collected, and RNA was extracted with Tri-reagent (Sigma) following the manufacturer’s instruction. A small fraction of cells was kept for estimating the fraction of GFP positive cells by flow cytometry.

### ChIP-STARR-seq RNA and plasmid sequencing

ChIP-STARR-seq RNA libraries were prepared as described^9^, with minor modifications. Briefly, RNA was isolated with Trizol (Sigma), following manufacturer’s instruction and split into two replicates. The mRNA fraction was captured from each replicate of total RNA using Oligo (dT) 25 beads (61002, Life Technologies), DNaseI treated (18068-015, Life Technologies), and reverse transcribed with a GFP-mRNA specific primer (149 STARRseq rep RNA cDNA synth) following Superscript III protocol (18080-044, Life Technologies). To avoid plasmid DNA contamination, cDNA was PCR amplified in 10 parallel reactions for each replicate using primers spanning a synthetic intron (152 STARR reporter specific primer 2 fw and 153 STARR reporter specific primer 2 rv), as previously described^11^. 35 ng of PCR amplified DNA were then used in a second round of PCR to add Illumina index primers (E7335, E7500, NEB). PCR was performed in 5 parallel reactions and using 2 independent index primers for each of the two replicates. In total, we sequenced 4 replicates of STARR-seq RNA for each pooled HIST or TF plasmid library. Corresponding plasmid libraries were similarly amplified using two independent Illumina index primers for each plasmid library. Quantity and quality of generated sequencing libraries was assessed on an Agilent Tapestation. All sequencing occurred on an Illumina HiSeq 2500 platform, using 50bp paired-end sequencing.

### ATAC-seq

The Assay for Transposase Accessible Chromatin coupled to high-throughput sequencing (ATAC-seq) was performed as previously described^60^ with slight modifications. Briefly, approximately 50.000 single cells were lysed in Resuspension buffer containing 0,1% IgePal, 0,1% Tween-20 and 0,01% Digitonin. Immediately after lysis, the buffer was washed out by Resuspension buffer with only Tween-20. Nuclei were centrifuged at 500xg for 10 min at 4°C. Nuclei were re-suspended in 25 μl 2x TD buffer (Illumina), 2.5 μl TDE1 (transposase, Illumina),

16.5 μl PBS, 0.5 μl Tween-20, 0.5 μl Digitonin, and incubated for 30 min at 37°C. Afterward, the sample was purified using the MinElute PCR Purification Kit (QIAGEN) according to the manufacturer’s protocol and eluted with 10 μl elution buffer. Samples were amplified using the NEBNext High Fidelity PCR master mix (New England Biolabs) and afterwards purified using a MinElute PCR purification kit (Qiagen) according to the manufacturer’s instructions. One microliter was loaded on an Agilent Technologies 2100 Bioanalyzer using a DNA 1000 assay to determine the library concentration and for quality check.

Cluster generation was performed according to the Illumina TruSeq SR Rapid Cluster kit v2 (cBot) Reagents Preparation Guide (www.illumina.com). Briefly, for sequencing libraries were pooled together to get a stock of 10 nM. One microliter of the 10 nM stock was denatured with NaOH, diluted to 10 pM and hybridized onto the flowcell. The hybridized products were sequentially amplified, linearized and end-blocked according to the Illumina Single Read Multiplex Sequencing user guide. After hybridization of the sequencing primer, sequencing-by-synthesis was performed using the HiSeq 2500 with paired end 50-cycle protocol followed by dual index sequencing. ATAC-seq libraries were sequenced to a depth of 20x10^6^ reads.

### RNA-seq

For RNA sequencing of ESCs, we used our previously published data^59^. For RNA sequencing of NSCs, RNA was isolated from two independent cultures following TRI reagent (Sigma) manufacturer’s instruction. RNA was further purified using column purification (Qiagen, #74204). mRNA capture, library prep and barcoding were performed according to standard procedures of the Erasmus MC Biomics facility. Libraries were then single end sequenced, 50bp, on an Illumina HiSeq2500 machine at a depth of approximately 20 million reads per sample.

### Data processing

The FASTQ files of RNA-, ChIP-, ATAC- and STARR-seq data were trimmed using Trimmomatic (version 0.39)^61^ to remove possible adaptor contamination and remove low quality reads. The trimmed RNA- and ATAC-seq data were mapped to the GRCh38/hg38 human genome build using the HISAT2 aligner (version 2.2.1)^62,63^. For RNA-seq, aligned reads were counted for each gene using htseq-count (version 0.12.4)^64^ and further processed using edgeR^65^ to normalize the data and perform downstream analysis. Bowtie2 (version 2.4.2) (with --very-sensitive parameter)^66^ was used to align both ChIP- and STARR-seq data against the GRCh38/hg38 human genome build. Only properly and uniquely mapped reads, with mapping quality more than 30 (MAPQ >=30), were kept followed by removing possible duplicated reads (only for ChIP-seq data) using Picard’s MarkDuplicates (version 4.0.1.1) (http://broadinstitute.github.io/picard/). The aligned reads were converted to bigwig format using ”bamCoverage” function with --binSize 10 and --normalizeUsing CPM parameters, the read coverage was then computed using “multiBigwigSummary” and pearson correlation between replicates was calculated using “plotCorrelation” via deepTools (version 3.4.3)^67^. Peak calling was performed using MACS2 (version 2.1.2) (with -q 0.05)^68^ for each replicate of ESCs and NSCs separately, and took sequencing data of total input as background. Peaks overlapping blacklist regions defined by the ENCODE project (Accession: ENCFF356LFX, Date: 05.05.2020) were removed. The overlapping peaks between replicates were merged as common peaks using intersectBed (version2.30.0)^69,70^.

### NCRE activity analysis

To generate a non-redundant set of scaffold regions to assess activity of NCREs, ChIP-seq and RNA-plasmid peaks (H3K4me1, H3K27ac, YY1 and SOX2 of ChIP-seq and histone and TF plasmid DNA-seq libraries of STARR-seq) were merged after extending the peak summit to a width of 1000 bp (500 bp from each side). All regions larger than 1000 bp were split in half until all regions were at most 1000 bp in length, avoiding large regions and preserving high genomic resolution for later analysis. All further analysis for both NSC ChIP-STARR-seq and comparative ChIP-STARR-seq was performed on these scaffold regions.

To define active regions, reads under the defined scaffolds in both NSC and comparative ChIP-STARR-seq in ESCs were counted by FeatureCount^71^. Read counts for each scaffold region were normalized by the total number of aligned reads in each library, after removing low coverage regions with less than 20 reads in at least two samples. The average number of normalized reads between replicates was taken for computing NCRE activity followed by the ratio of RNA-seq and DNA-seq (plasmid) reads as measurement of NCRE activity. The scaffold regions were ranked based on the average activity of RNA-seq samples in 5 different, equally sized, categories and the top-10% regions were defined as the most active NCREs in each cell line.

As NSC ChIP-STARR-seq we define those experiments that used NSC-derived plasmid libraries transfected in NSCs. As comparative ChIP-STARR-seq we refer to those data obtained from the transfection of NSC-derived plasmid libraries in ESCs. When discussing ESC ChIP-STARR-seq, we refer to our previous study that generated data obtained from transfections of ESC-derived plasmid libraries in ESCs^9^. Furthermore, we defined cell-specific regions as the overlapping regions between active regions (top-10%) of a given cell line and regions with less activity (bottom90%) of another cell line (i.e., from NSC ChIP-STARR-seq or comparative ChIP-STARR-seq in ESCs). We also considered the active regions that were in the top-10% of both NSC ChIP-STARR-seq and comparative ChIP-STARR-seq as common highly active regions.

### NCRE-gene interaction

To integrate gene expression data, NCREs were linked to their target genes using HiC data and distance to the closest TSS. Previously published HiC data from the germinal zone (GZ) at gestation weeks 17–18 of 3 human foetal brains were used^17^ to link NSC NCREs to target

genes. This data provides 10 kb resolution bins for gene loop interactions and 40 kb resolution for TADs. Moreover, to link NCREs identified in comparative ChIP-STARR-seq in ESCs to their target genes, we used published HiC data from H9 human ESCs with 25 kb resolution bins for both gene loop interactions and TADs^36^.

NCREs were then intersected with pre-calculated significant interactions using intersectBed to define NCRE-gene interaction. NCRE-gene interactions located within the same TAD were considered for downstream analyses. The coordinates of HiC data were liftedOver to the GRCh38/hg38 human genome build before intersecting with regions. In addition, NCREs were annotated for target genes based on the distance to nearest TSS using the “annotatePeaks.pl” function of HOMER (version 4.11)^72^. Finally, all NCRE-gene interactions either using HiC or distance to closest TSS were considered for further analysis. In total, we found 28,514 target genes (coding and non-coding) for NSCs and 20,347 target genes for ESCs, of which 19,739 target genes are identified based on the closest TSS and the other target genes are based on HiC.

We only included protein-coding genes in our analysis. Gene expression levels were plotted and statistical comparison was performed using Wilcoxon signed rank test in R. The *Online Mendelian inheritance in Man* (OMIM) gene list (updated 21-04-2022) was downloaded from the OMIM database to further assist in annotation of target genes.

### Gene ontology analysis

The web interface of Enrichr^18,19^ (https://maayanlab.cloud/Enrichr/) was used for functional enrichment analysis using the default settings, and the whole genome was set as background. The geom_point function of ggplot2 (version 3.5.0) was used to visualize the GO enrichment.

### Functional enrichment

NCREs were annotated for genome features using the “annotatePeaks.pl” function of HOMER (version 4.11)^72^. Motif enrichment analysis was performed using the “findMotifsGenome.pl” function of HOMER ^72^ with default setting and whole genome as background. Significant enriched motifs were selected based on p-value ≤ 0.01.

### Sequence characteristics analysis

To determine whether different DNA sequence features distinguish defined regions between NSCs and ESCs, we considered the following features: (i) the non-coding essential regulation (ncER) score (https://github.com/TelentiLab/ncER_datasets/; updated 06-03-2019)^20^; (ii) GC content, as determined by the GCcontent R packages based on BSgenome. Hsapiens.UCSC.hg38 (version 1.4.3); (iii) conservation score for each NCRE, as derived from the gscores R packages based on phastCons100way.UCSC.hg38 (version 3.7.2)^21^; (iv) Orion scores^22^; (v) CADD scores^23^; (vi) Haploinsufficiency scores^27^ and (vii) probability of loss-of-function intolerance (pLI) score^73^. The overlaps between DNA sequence features and regions were defined using intersectBed. The coordinates of data were converted to the GRCh38/hg38 human genome build by liftOver prior to intersections. The scores of the genes linked to the regions were plotted to show pLI distribution. Statistically significant differences between groups were determined using the Wilcoxon signed rank test in R.

### Transposable element enrichment

The RepeatMask (GRCh38/hg38, updated 03-09-2021) was downloaded from the UCSC table browser and intersected to the regions. To determine enrichment of transposable elements in NCREs, we followed a strategy previously used for investigating active NCREs in human embryonic stem cells^9^. The number of overlaps of each type of repeat (n_overlaps) with all regions (n) was used to calculate the relative frequency (f_all = n_overlaps/n). Multiplication of the relative frequency with the number of regions (n_test, e.g., ESC, NSC etc.) in any tested group yields the expected frequency (E). This number was compared with the actual observed frequency in the subgroups (f_test = (n_overlap, test)/n_test = O) to calculate the observed versus expected ratio (O/E). We considered repeats with O/E > 2 as enriched. For the subsequent data interpretation, we only focused on transposable elements that were present multiple times (n_overlap > 15).

### Epigenome profiling of NCREs

To find the enrichment of endogenous marks around NCREs in ESC and NSC, we collected various ChIP-seq data from previously published studies^9,54,74^ and from the ENCODE project^75^.

The FASTQ files of ChIP-seq data was re-analyzed along with ATAC-seq data (from this study) based on what we described in the “*Data processing*” section.

Heatmaps for differentially active regions across the endogenous marks were created using deeptools (version 3.4.3)^67^. We first converted bam files of endogenous marks into bigwig format using the “bamCoverage” function with --binSize 10 and --normalizeUsing RPKM parameters. The bigwig files were then used to compute the number of reads across 6 kb centered on NCREs using “computeMatrix” with --referencePoint center, --upstream 3000 and --downstream 3000 parameters. Finally, the line plots and heatmaps were generated using the “plotProfile” and “plotHeatmap” functions, respectively.

### Predicting NCRE activity using DeepSTARR

In our initial attempts to predict NCRE activity based on DNA sequence composition (**Extended Data. 6a**), we used the previously published DeepSTARR framework^50^ As this model requires 249 bp input sequences, we first generated 249 bp regions centered on the NCRE regions. Subsequently, we applied the prediction function using the default settings of DeepSTARR, as instructed on the author’s Github repository (https://github.com/bernardo-de-almeida/DeepSTARR). Finally, we calculated the Pearson correlations between DeepSTARR output and ChIP-STARR-seq experimental results.

### Construction of BRAIN-MAGNET

To generate BRAIN-MAGNET, we first removed from the analysis scaffold of 148,198 genomic regions assessed in ChIP-STARR-seq a small number of patch sequences resulting in a final number of 148,114 genomic sequences. These ranged from 500 to 1000 base pairs in length, and were augmented with their reverse complement to yield a total of 296,228 examples. A random 10% of the augmented sequences were held out for both validation and testing purposes. The DNA sequences were extracted from the human genome using bedtools getfasta (version 2.30.0) and their reverse complements were obtained using revseq of EMBOSS (version 6.6.0.0)^76^. The input sequences were then converted into an one-hot encoded matrix of length 1000, with each nucleotide being represented by a binary vector (A = [1,0,0,0], C = [0,1,0,0], G = [0,0,1,0], T = [0,0,0,1]). Sequences shorter than 1000 bp were padded with zeros. The corresponding sequence activities served as outputs of the model. We subsequently used input and output in BRAIN-MAGNET, a single task convolutional neural network, to predict either activities of NCREs identified by NSC ChIP-STARR-seq or by comparative ChIP-STARR-seq in ESCs. The model is composed of three convolutional layers (filters = 128, 256, 512; size = 11, 9, 7), each followed by batch normalization, a ReLU non-linearity, and MaxPool (size = 2). After the convolutional layers, there are two fully connected layers, each with 1024 neurons, followed by batch normalization, a ReLU non- linearity, and dropout where the fraction is 0.4. The final layer is mapped to either NSC or comparative ChIP-STARR-seq outputs.

Hyperparameters were manually adjusted to optimize the performance on the validation set. The model was implemented and trained in Pytorch (version 1.13.1 with CUDA version 11.7)^77^ using the Adam optimizer with a learning rate of 0.01, mean squared error (MSE) as the loss function, a batch size of 128, and early stopping criterion with a patience of 20 on 100 epochs. Model training, hyperparameter tuning, and performance evaluation were conducted on different sets of genomic regions.

The performance of the model was evaluated separately for NSC and ESC predictions on the held-out test sequences. We used the Pearson and Spearman correlation coefficient (R and Rho) across all bins for a quantitative genome-wide evaluation (**Extended Data Fig. Sb**). Furthermore the area under the precision-recall and receiver operating characteristic curve (PR and ROC; calculated using pr.curve and pr.roc functions from R package PRROC (version 1.3)^78^ were used for NCRE classification in the various activity groups (top-10% vs. the other defined NCRE activity classes) (**Extended Data Fig. 6**c). To benchmark BRAIN-MAGNET to existing variant prioritization scoring methods, we first retrieved LINSIGHT^44^, CADD^23^, GERP^45^ and ncER^20^ scores for all nucleotides for which BRAIN-MAGNET provides cb scores. We subsequently calculated the Pearson correlation between BRAIN-MAGNET cb scores and the outputs of the other variant prioritization scoring methods (**Extended Data Fig. 6**d). To reduce the computational burden, for this analysis we used the previously used test set of NCREs, containing randomly selected 10% of our NCRE atlas. Furthermore, we similarly calculated and plotted the correlation between BRAIN-MAGNET cb scores and LINSIGHT, CADD, GERP and ncER score for all nucleotides of the functionally investigated motifs and mutations studied in this work (**Extended Data Fig. 6**e), and visualized all these scores as heatmaps for the individual studied loci.

### Motif discovery using TF-MoDISco-lite

DeepExplainer^42^, a DeepSHAP implementation of DeepLIFT, was used to compute contribution (cb) scores for all nucleotides of the assessed NCREs, which reflects the individual contribution of each nucleotide to the NCRE activity in either NSCs or ESCs. 100 dinucleotide-shuffled versions of each input sequence were used as reference sequences (the fork from https://github.com/kundajelab/shap/blob/master/shap/explainers/deep/deep_pytorch.py). The obtained hypothetical importance scores were then multiplied by the one-hot encoded matrix of the sequences to derive the actual nucleotide cb scores, which were visualized using the ggseqlogo (version 0.2)^79^. The cb scores of each NCRE can be visualized using the R-shinny app for which the code is available in our github repository: https://github.com/ruizhideng/NSC-ChIP-STARR-seq_paper.

The hypothetical importance scores, along with the corresponding one-hot encoded matrix of the sequences, were subjected to TF-MoDISco-lite (https://github.com/jmschrei/tfmodisco-lite), a more efficient and user-friendly version of TF-MoDISco^41^, to identify the most relevant motifs associated with changes of each nucleotide. The TF-MoDISco-lite algorithm in default settings was used to extract and cluster similar seqlets across all selected sequences, the identified seqlets were matched against JASPAR 2022 CORE vertebrate non-redundant database (https://jaspar.genereg.net/download/data/2022/CORE/JASPAR2022_CORE_non-redundant_pfms_jaspar.zip) by Tomtom (version 5.5.1)^80^ within TF-MoDISco-lite^41^. To further facilitate the interpretation of the cb scores across the genome for downstream analysis, we first converted the negative cb scores to positive values allowing to include the impact of enriched motifs at sequences with negative cb scores. Subsequently, we calculated normalized cb scores taking either a normalisation of all NCREs (cb_all) or normalisation within each NCRE (cb_each) into account, and calculated percentile scores for these. All raw and normalized cb scores used throughout the study are (given the file size) available via a defined USCS browser session: https://genome.ucsc.edu/s/BarakatLab/BrainMagnet_NSC_ESC_cb_scoreshg38

### GWAS analysis

To test the utility of BRAIN-MAGNET cb scores for the functional interpretation of GWAS loci, we made use of a recent MPRA data set provided by Guo et al, which tested 2,221 non-coding variants associated with 10 neuropsychiatric disorders by MPRA, with an average of 5 SNPs per disease risk loci being tested^46^. Comparing the activity of matched risk versus reference allele pairs, the authors identified 892 SNPs that showed differential activity in their MPRA depending on the genotype of the given SNP, which the authors refer to as daSNVs. We retrieved the daSNVs and non-daSNVs from Supplementary Data 3 and 5 of Guo et al ^46^ and intersected these with NCREs and cb scores from our data set. We subsequently plotted the distribution of cb scores (>60 cb_all) over the daSNV and non-daSNVs and additionally plotted the GERP, LINSIGHT, CADD and ncER scores for the same SNPs. Furthermore, we used the BRAIN-MAGNET cb scores to fine map two previously identified GWAS loci for which multiple SNPs in LD were assessed in the Guo et al study and present in multiple NCREs from our data set.

### Genomic variant analysis

For the gnomAD variant analysis, the curated VCFs of gnomAD v4 were downloaded from https://gnomad.broadinstitute.org/downloads#v, which contains 909,084,110 short variants mapped to the GRCh38 genome build obtained from whole-genome sequencing of 76,215 individuals. bcftools (v1.8) was used to keep high-quality variants (“PASS” flag) and rare variants (AF < 0.1%). tabix (v1.9) was used to extract the variants overlapping our NCREs.

The total number of high-quality rare variants was calculated without chromosome Y and mitochondrial DNA (chromosome M), as these sequences were not covered in our NCRE analysis scaffold. The VISTA enhancers, ENCODE_cCREs, fiveUTR, threeUTR and codingExon regions (GRCh38/hg38, updated 14-02-2024) were downloaded from the UCSC table browser. DAE and nDAE regions were retrieved from Yousefi et al^26^. The total number of filtered rare variants (MAE <0.1) genome wide (variant_genome) was normalized by the size of the genome (size_genome) and used to calculate the expected frequency (E = n_variant/size_ genome). The number of filtered rare variants located in the target regions (listed in **Extended Data Fig. 5**a) (variant_region) was normalized by the size of the target region (size_region) to calculate the observed frequency (O = variant_region/size_region). The O/E ratio was calculated by comparing the observed frequency over the expected frequency.

For the *de novo* variant analysis from the Genomics England 100,000 Genomes Project (GEL), *de novo* variants of GEL data version 18 (main-programme_v18_2023-12-21) were retrieved by Labkey from the *de novo*_flagged_variants table in the GRCh38 genome build. Next, high confidence *de novo* variants (stringent_filter = 1) were kept. Then, solved cases, labelled with “yes”, “unknown” and “partically” in the gmc_exit_questionnaire table, were removed. Finally, 4,415 individuals with unexplained neurology and neurodevelopmental disorders were kept, for which 13,276 *de novo* variants overlapped with our NSC NCREs. Also we analysed 4,558 genomes from individuals with a variety of other rare disease categories (including cardiovascular disorders, hearing an ear disorders, ultra-rare disorders, dermatological disorders, gastroenterological disorders, renal and urinary tract disorders, respiratory disorders, endocrine disorders, dysmorphic and congenital abnormality syndromes, growth disorders, skeletal disorders, ophthalmological disorders, metabolic disorders, hematological and immunological disorders and rheumatological disorders), harbouring 13,564 *de novo* variants overlapping NSC NCREs. The *de novo* variants were further annotated by the BRAIN-MAGNET cb scores, and the cb_each distribution of NCREs were plotted.

For the GEL analysis using variants overlapping with high-confidence motifs, we used 20 bp sequences with coordinates centred on the high-confidence motifs called by TF-MoDISco (q-values < 0.05) from the NSC NCRE category 4 and 5. An aggregation of gVCFs (AggV2, only available in version 10) comprising 78,195 genomes from GEL harbouring over 722 million variants was intersected with these high-confidence motif coordinates. Only variants marked with the “PASS” flag, indicating high quality, were retained, resulting in 11,788 variants overlapping 2,980 motifs. We next retained only variants in unsolved individuals (55,379 genomes, 9,787 variants) and subsequently individuals with neurodevelopmental disorders (10,536 genomes, 4,259 variants). Next we retained only variants with allele count (AC) <= 10 (2,700 genomes, 2,684 variants). Finally, only variants located in NCREs with target genes recorded as OMIM disease genes were kept, resulting in a final list of 705 variants from 824 genomes that overlapped 440 motifs.

For screening of an independent WGS data set, we first used 330 trio WGS from individuals affected by rare disorders from the Institute of Medical Genetics and Applied Genomics (IMGAG) database from Tübingen, Germany. Removing those individuals in which already a definitive molecular diagnosis was found (n=81) left 249 individuals, of which 60 harbored a candidate disease explaining variant. In those 249 individuals, a total of 16,437 *de novo* variants was found. Intersecting these variants with the NCRE atlas and filtering for variants with a BRAIN-MAGNET cb score >95th percentile resulted in 70 *de novo* variants that were filtered for potential clinical phenotype matches by HPO terms and manual curation. Finally, 3,971 singleton WGS of undiagnosed individuals were screened for rare deletions overlapping with NCREs that contained nucleotides with a cb score >95th percentile. Among the 12,172 deletions called in this data set, 1,584 were rare (filtered by <10 exact in-house calls and <50 larger in house deletion calls). For 117 of those deletions, the associated HPO terms of the target gene of the deleted NCRE matched the patient HPO terms. We then manually curated for matching clinical phenotypes, focusing on deletions <20 kb in size not containing additional coding sequences, resulting in the 8 NCRE deletions, of which 3 are potentially disease- explaining and 5 represent carrier deletions with currently no second coding hit identified.

### Experimental validation of wild type and mutant NCREs *in vitro*

For experimental validation, we randomly selected 15 NCREs of which the target gene was expressed in NSCs. NCREs were amplified from genomic DNA and cloned into the STARR-seq reporter plasmid (kind gift of A.Stark)^11^ as previously described^9^. Subsequently, site-directed mutagenesis was used to generate NCRE deletions or point mutations in the generated plasmids. NSCs were then transfected with wild type or mutant NCRE plasmids using polyethylenimine (PEI, Sigma) or Lipofectamine™ Stem Transfection Reagent (Thermo Scientific) respectively. Twenty-four hours post transfection cells were collected, stained with Hoechst dye and the NCRE activity was measured by FACS analysis (20,000 cells per sample). GFP-positive cells within the mCherry-positive population were quantified to assess NCRE activity compared to an empty STARR-seq vector control. Two independent transfection experiments were performed, each in duplicates. Statistical analysis was performed using a one-way ANOVA test followed by multiple comparison test (Fisher’s LSD test). Calculations were conducted in GraphPad Prism (version 8).

For CRISPRi experiments, we first generated a HEK293 cell line stably expressing ZIM3- KRAB-dCas9^81^ upon doxycycline induction, by transduction with pLX303-ZIM3-KRAB-dCas9 (a kind gift from Mikko Taipale, Addgene plasmid # 154472). gRNAs targeting the *RAB7A* NCRE were cloned into a pgRGFP plasmid (Addgene #82695^82^, a kind gift of Allan Mullen) and transfected into the ZIM3-KRAB-dCas9 expressing cells using polyethylenimine (PEI, Sigma). 48h post transfection cell were collected for RNA isolation (Tri reagent, Sigma). cDNA was generated with the iSCRIPT cDNA synthesis kit (Bio-Rad) and the change in expression of *RAB7A* was quantified by qRT-PCR (iTaq universal SYBR Green Supermix) (Sigma), performed in a CFX96RTS thermal cycler (Bio-Rad), as previously described^59^. The ΔΔct method was used to calculate the fold change in gene expression, using TBP expression as housekeeping normalisation control. Statistical analysis was performed using one-way ANOVA test and calculations were conducted in GraphPad Prism (version 8).

### Transgenic NCRE assays in zebrafish

Zebrafish (Danio rerio) were raised and maintained under standard conditions, as previously described^26^. Adult and larval fish were kept on a 14h/10h light–dark cycle at 28°C. Larvae were kept in HEPES-buffered E3 medium. Media was refreshed daily and at 24 hours post fertilization (hpf) 0.003% 1-phenyl 2-thiourea (PTU) was added to prevent pigmentation. All zebrafish experiments were performed in compliance with Dutch animal welfare legislation and only larvae <5 days post fertilization (dpf) were used for experimentation. The wild type and mutant *RAB7A* NCRE used in the *in vitro* experiments were transferred by Gibson assembly between the AscI and PacI sites of an E1b-GFP-Tol2 enhancer assay plasmid (a kind gift from Ramon Birnbaum^83^) containing an E1b minimal promoter followed by GFP, using the following transfer primers: Transfer_fw: 5’- AGATGGGCCCTCGGGTAGAGCATGCACCGG-3’ and Transfer_rv: 5’-TCGAGAGATCTTAATGGCCGAATTCGTCGA-3’. Constructs were injected into fertilized eggs at the 1 cell stage (25 pg per egg) using standard procedures, with or without 25 pg transposase mRNA. At least 50 eggs were injected per construct in at least two different injection experiments. GFP expression was observed and annotated at 24 hpf, using a fluorescent Leica M165FC stereomicroscope. Embryos representative of each category were subsequently imaged on a Leica SP5 intravital imaging setup with a 20 × /1.0 NA water-dipping lens. Images were analyzed using imageJ (FIJI).

---

**Extended Data Fig. 1:**
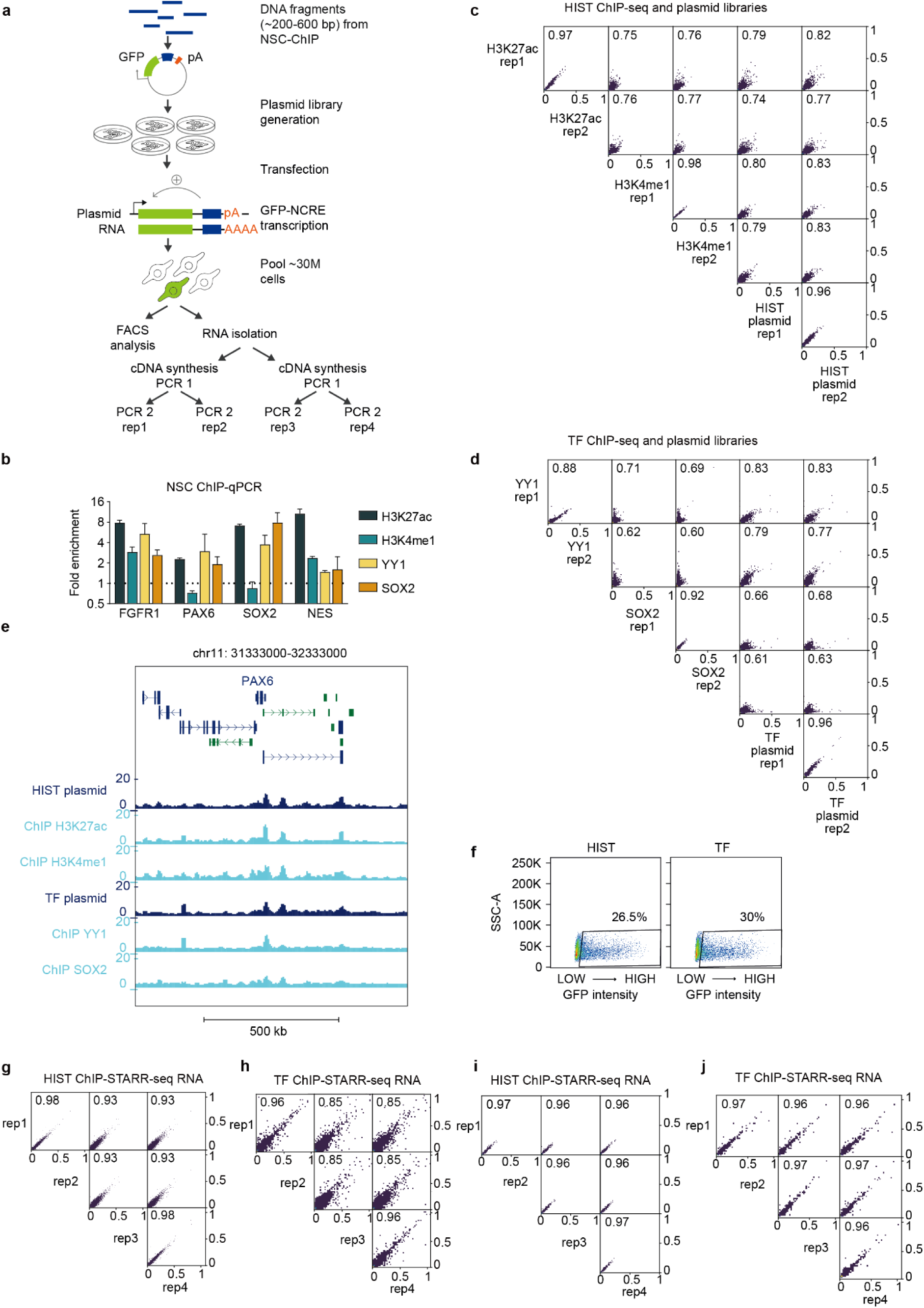
Experimental strategy and quality control for ChIP-STARR-seq. a) Schematic representation of the experimental strategy used in this study. b) ChIP-qPCR in NSC showing the average fold enrichment over total input, normalized to a non-bound site in *NCAPD2* (for H3K4me1 and H3K27ac histone ChIP) or in *ACTB* (for YY1 and SOX2 ChIP). Boxes represent the average of 2 replicates. Error bars represent standard deviations. c) Scatterplot depicting the correlation between read counts (ln(CPM)) per genomic regions between the H3K27ac and H3K4me1 ChIP-seq and the HIST ChIP-STARR-seq plasmid libraries. Reported is the Pearson correlation coefficient. d) As in c) but now for YY1 and SOX2 ChIP-seq and for the TF ChIP-STARR-seq plasmid libraries. e) Genome browser tracks surrounding the *PAX6* locus showing good representation of H3K27ac, H3K4me1, YY1 and SOX2 ChIP-seq signals (light blue) in the HIST and TF ChIP-STARR-seq plasmid libraries (dark blue), and correlation between ChIP-STARR-seq RNA tracks (green) f) FACS plot showing GFP expression of NSCs transfected with the HIST or TF ChIP-STARR-seq plasmid libraries. g) Scatterplot depicting the correlation between read counts (ln(CPM)) per genomic regions between the HIST ChIP-STARR-seq RNA replicates from transfection of HIST plasmid libraries in NSCs (i.e., NSC ChIP-STARR-seq). Reported is the Pearson correlation coefficient. h) As in g) but now for TF ChIP-STARR-seq RNA replicates from transfections of TF plasmid libraries in NSCs (i.e., NSC ChIP-STARR-seq). i) As in g) but now for HIST ChIP-STARR-seq RNA replicates from transfections of HIST plasmid libraries in ESCs (i.e., comparative ChIP-STARR-seq in ESCs). j) As in g) but now for TF ChIP-STARR-seq RNA replicates from transfections of TF plasmid libraries in ESCs (i.e., comparative ChIP-STARR-seq in ESCs).

**Extended Data Fig. 2:**
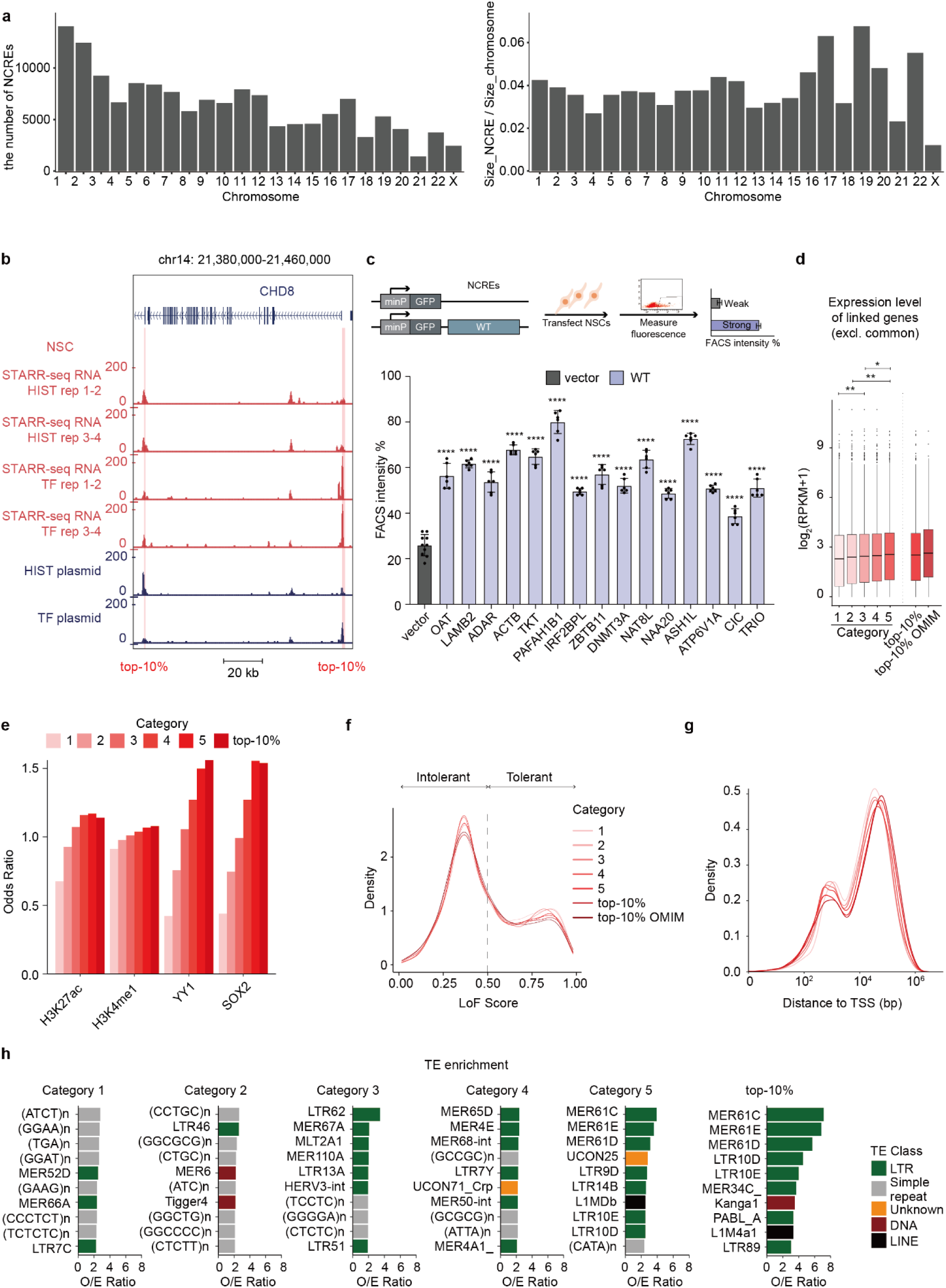
Extended data for identification of active NCREs in NSCs a) Bar graph showing the number of NCRE scaffold regions per chromosome (left) and bar graph showing the ratio between total NCRE scaffold size divided by chromosome size (right). Both graphs exclude chromosome Y and chromosome M (mitochondrial DNA) which were not included in the analysis scaffold. b) Genome browser tracks surrounding the *CHD8* locus showing two NCREs belonging to the top-10% category of activity. c) Bar plot showing the percentage of GFP+ cells in NSCs upon cell transfection experiments with reporter plasmids containing either empty reporter plasmid control (grey) or 15 different wild type NCREs (blue). NCREs are indicated with the name of the presumed target gene. Plotted on the left is the percentage of GFP+ in cells co-transfected with an mCherry expressing plasmid, to correct for transfection efficiency. Bars show the average from two independent experiments, with each NCRE tested each in duplicate. Error bars represent standard deviation. * p < 0.05; ** p < 0.01; *** p < 0.001; **** p < 0.0001 (one-way ANOVA test followed by multiple comparison test (Fisher’s LSD test)). d) Expression level (log2(RPKM+1)) of the genes linked to each of the 5 NCRE activity categories, to the top-10% most active NCREs and of the OMIM disease-related genes linked to the top-10% most active NCREs, considering only the genes unique for each NCRE category. Boxes represent the interquartile range (IQR); lines represent the median; whiskers extend to 1.5 the IQR; dots represent outliers. (* p < 0.05; ** p < 0.01, Wilcoxon test). e) Bar plot showing the ratio between the overlap of each NCRE category with each ChIP-seq and the total overlap of the 148,198 scaffold regions with each ChIP-seq, both normalized for the group size. f) Kernel density plot showing the distribution of loss-of-function tolerance scores for non-coding sequences for all the NCRE categories from NSC ChIP-STARR-seq. g) Density plot showing the distance in bps to the TSS for all NCRE categories. h) Bar graph showing the 10 most enriched transposable elements (TEs) overlapping with the NCREs belonging to the different activity categories in NSCs. Plotted is a ratio between the observed (O) number of TEs over the expected (E).

**Extended Data Fig. 3:**
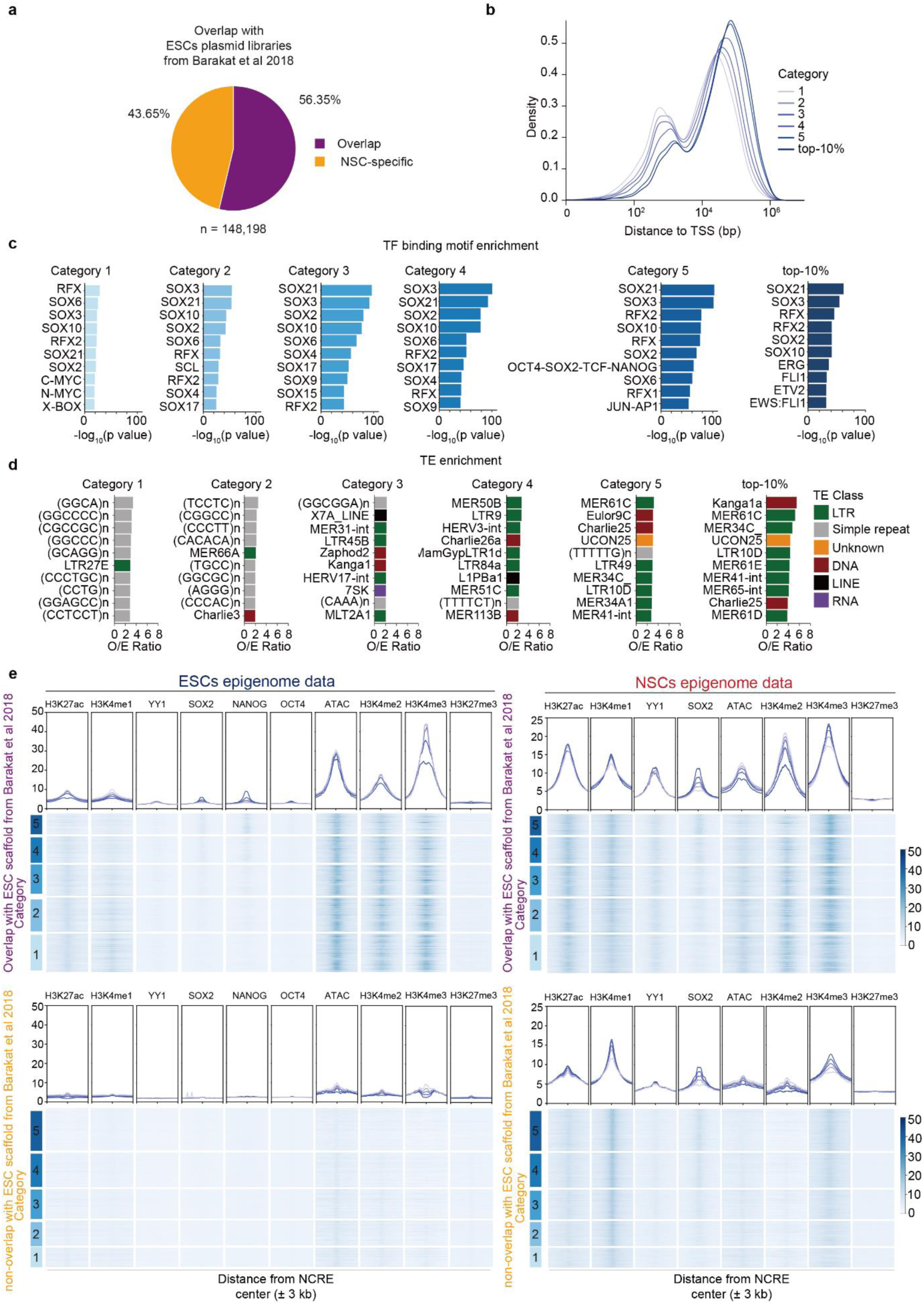
Extended data for comparative ChIP-STARR-seq using NSC derived plasmid libraries in ESCs a) Pie chart showing the fraction of NSC ChIP-STARR-seq scaffold regions overlapping with our previously studied ESC scaffold regions from ESC ChIP-STARR-seq^9^. b) Density plot showing the distance in bps to the TSS for all NCRE categories. c) Bar graph showing the 10 most enriched TF motifs in the NCREs belonging to the different activity categories from comparative ChIP-STARR-seq in ESCs. Plotted is the -log10 p-value, as calculated by Homer. d) Bar graph showing the 10 most enriched transposable elements (TEs) overlapping with the NCREs belonging to the different activity categories from comparative ChIP-STARR-seq in ESCs. Plotted is a ratio between the observed (O) number of TEs over the expected (E). e) Epigenome profiling of NCREs with comparative ChIP-STARR-seq activity in ESCs (plasmid libraries derived from ChIP experiments in NSCs). Results are shown for the 148,198 scaffold regions that overlap (top row) or do not overlap (bottom row) with the scaffold regions used earlier in ESC ChIP-STARR-seq^9^. For each row at the Top: line plot showing the average enrichment of ChIP-seq (or ATAC-seq) signal from ESCs (left panel) or NSCs (right panel) across the genomic regions encompassed by NCREs from the 5 categories of NCRE activity detected by comparative ChIP-STARR-seq in ESCs (bins of 3 kb up and down centred at the middle of the NCRE). For each row at the Bottom: corresponding heatmaps of H3K27ac, H3K4me1, YY1, SOX2, NANOG, OCT4, H3K4me2, H3K4me3 and H3K27me3 ChIP-seq signals and ATAC-seq in ESCs, or H3K27ac, H3K4me1, YY1, SOX2, H3K4me2, H3K4me3 and H3K27me3 ChIP-seq signals and ATAC-seq in NSCs across the assessed scaffold regions ranked by comparative ChIP-STARR-seq activity in ESCs from the highest to the lowest.

**Extended Data Fig. 4:**
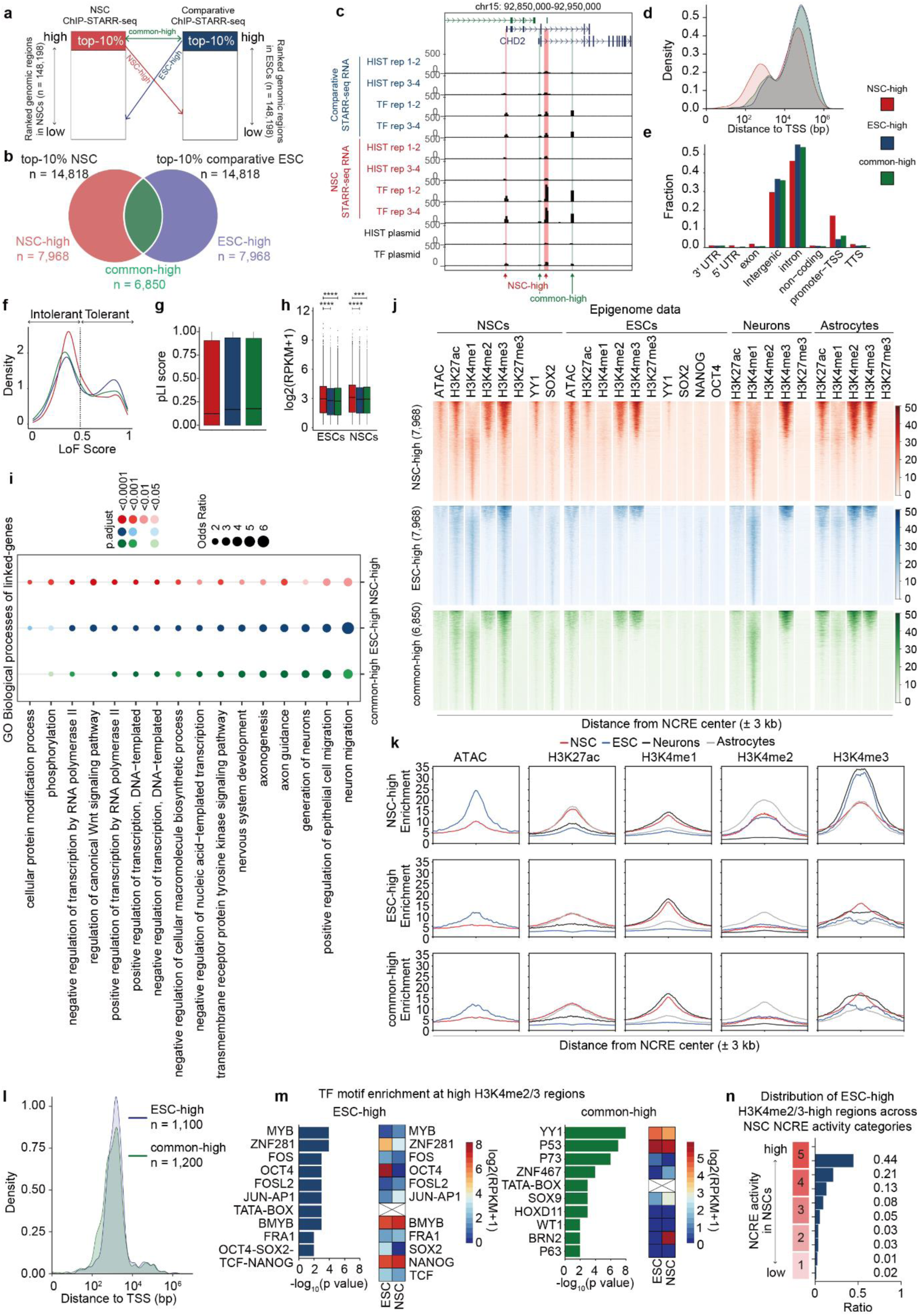
Comparative ChIP-STARR-seq identifies differentially active NCREs in NSCs and ESCs. a) Schematic overview of the process to define common-high (green arrow), NSC-high (red arrow) and ESC-high (blue arrow) NCREs. b) Venn diagram showing the overlap between the top-10% most active NCREs in NSCs and ESCs. c) Genome browser tracks surrounding the *CHD2* locus showing ChIP-STARR-seq RNA-seq and DNA-seq (plasmid) tracks. d) Density plot showing the distance to the TSS of differentially active NCREs (NSC-high (red) and ESC-high (blue)) and common highly active NCREs in ESCs and NSCs (common-high, green). e) Bar graphs showing the fraction of differentially active NCREs (NSC-high (red) and ESC- high (blue)) and common highly active NCREs in ESCs and NSCs (common-high, green) per genomic region using Homer. UTR = untranslated region; non-coding = non-coding RNAs such as snRNA, lncRNA, etc; TSS = Transcriptional start site; TTS = Transcriptional termination site. f) Kernel density plot showing the distribution of loss-of-function tolerance scores for non-coding sequences for differentially active NCREs (NSC-high (red) and ESC-high (blue)) and common highly active NCREs in ESCs and NSCs (common-high, green). g) pLI score of the genes linked to differentially active NCREs (NSC-high (red) and ESC-high (blue)) and common highly active NCREs in ESCs and NSCs (common-high, green). h) Expression level (log2(RPKM+1)) in ESCs and NSCs of the genes linked to differentially active NCREs (NSC-high (red) and ESC-high (blue)) and common highly active NCREs in ESCs and NSCs (common-high). Boxes represent the interquartile range (IQR); lines represent the median; whiskers extend to 1.5 the IQR; dots represent outliers. (* p < 0.05; Wilcoxon test). i) Gene ontology analysis using Enrichr for the protein-coding genes linked to differentially active NCREs (NSC-high (red) and ESC-high (blue)) and common highly active NCREs in ESCs and NSCs (common-high). j) Heat maps of ATAC-seq and ChIP-seq signals in NSCs, ESCs, neurons and astrocytes across the 7,968 NSC-high, 7,968 ESC-high and the 6,850 common-high active NCREs k) Line plots showing the average enrichment of ATAC-seq and ChIP-seq signal for H3K27ac, H3K4me1, H3K4me2 and H3K4me3 in NSCs (red), ESCs (blue), neurons (black) and astrocytes (grey) l) Density plot showing the distance to the TSS of the 1,100 ESC-high NCREs with high H3K4me2/3 m) Bar graphs showing the 10 most enriched TF motifs in the 1,100 ESC-high NCREs and the 1,200 common highly active NCREs with high H3K4me2/3. Plotted is the -log10 p-value. Flanking the bar graphs is a heatmap showing the expression (log2(RPKM+1)) of each TF in ESCs and NSCs. n) Bar graph showing the distribution of the 1,100 ESC-high NCREs with high H3K4me2/3 across the different categories of NCRE activity in NSCs. Indicated is the fraction of regions per category.

**Extended Data Fig. 5:**
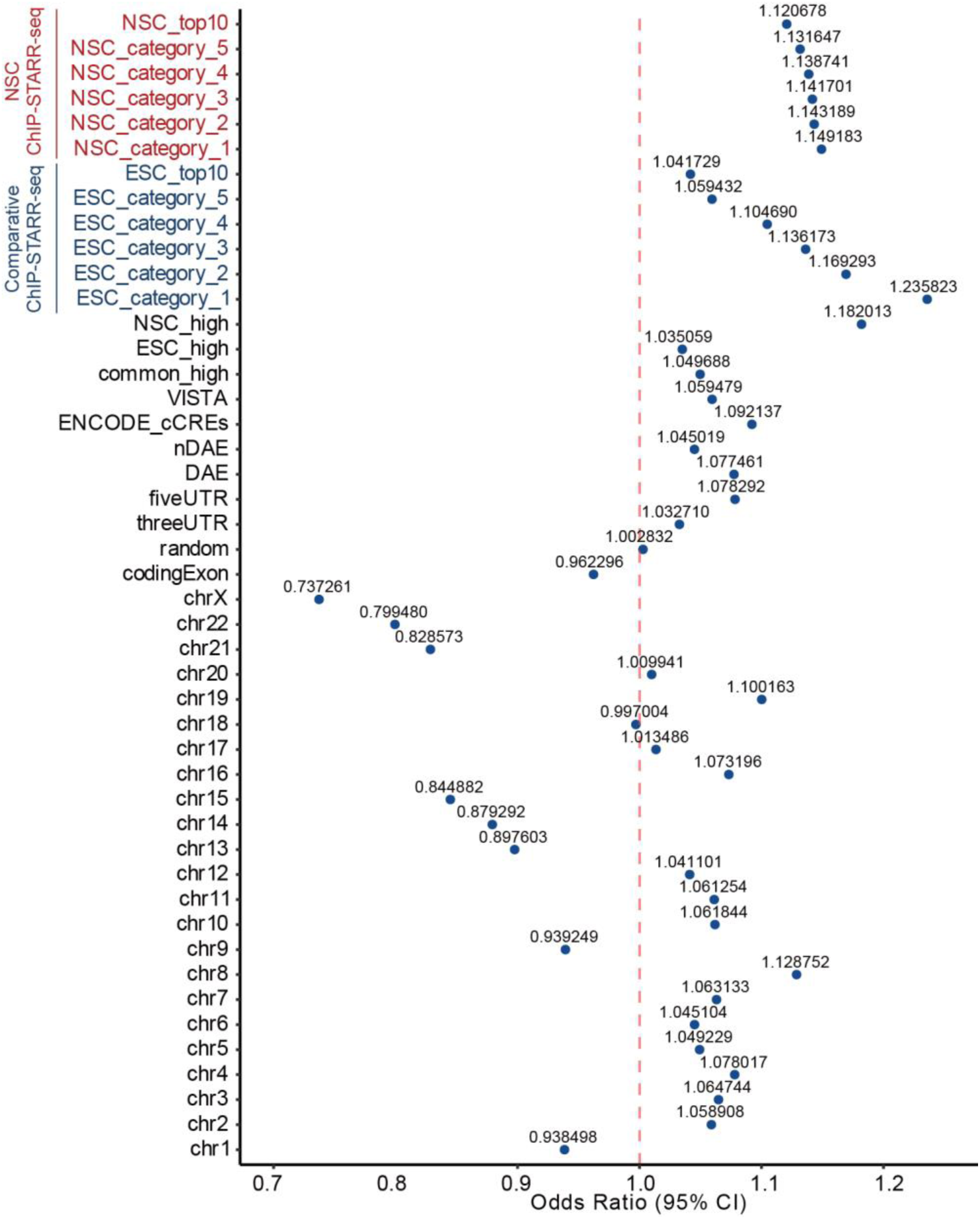
Extended data on rare variants in NCREs Plot showing the odds ratio of observed and expected rare variants (minor allele frequency <0.1%) from 76,215 genomes from individuals that did not present with early onset neurodevelopmental phenotypes from gnomAD v4^1^ that overlap with the various categories of genomic sequences: NSC category 1-5 and NSC top-10%, NCRE activity categories in NSCs; ESC category 1-5 and ESC top-10%, NCRE activity categories in ESCs; NSC_high, ESC_high and common_high, NCRE categories from the differential activity analysis (see **Extended Data Fig. 4**); VISTA^38^, ENCODE candidate cis regulatory elements^39^, nDAEs and DAEs^26^, 5’ and 3’ UTR regions and coding exome obtained from the UCSC Genome browser; 148,114 random regions. For reference, the odds ratio was also plotted for the various individual chromosomes (amongst others, the acrocentric chromosomes and chromosome X are depleted for rare variants).

**Extended Data Fig. 6:**
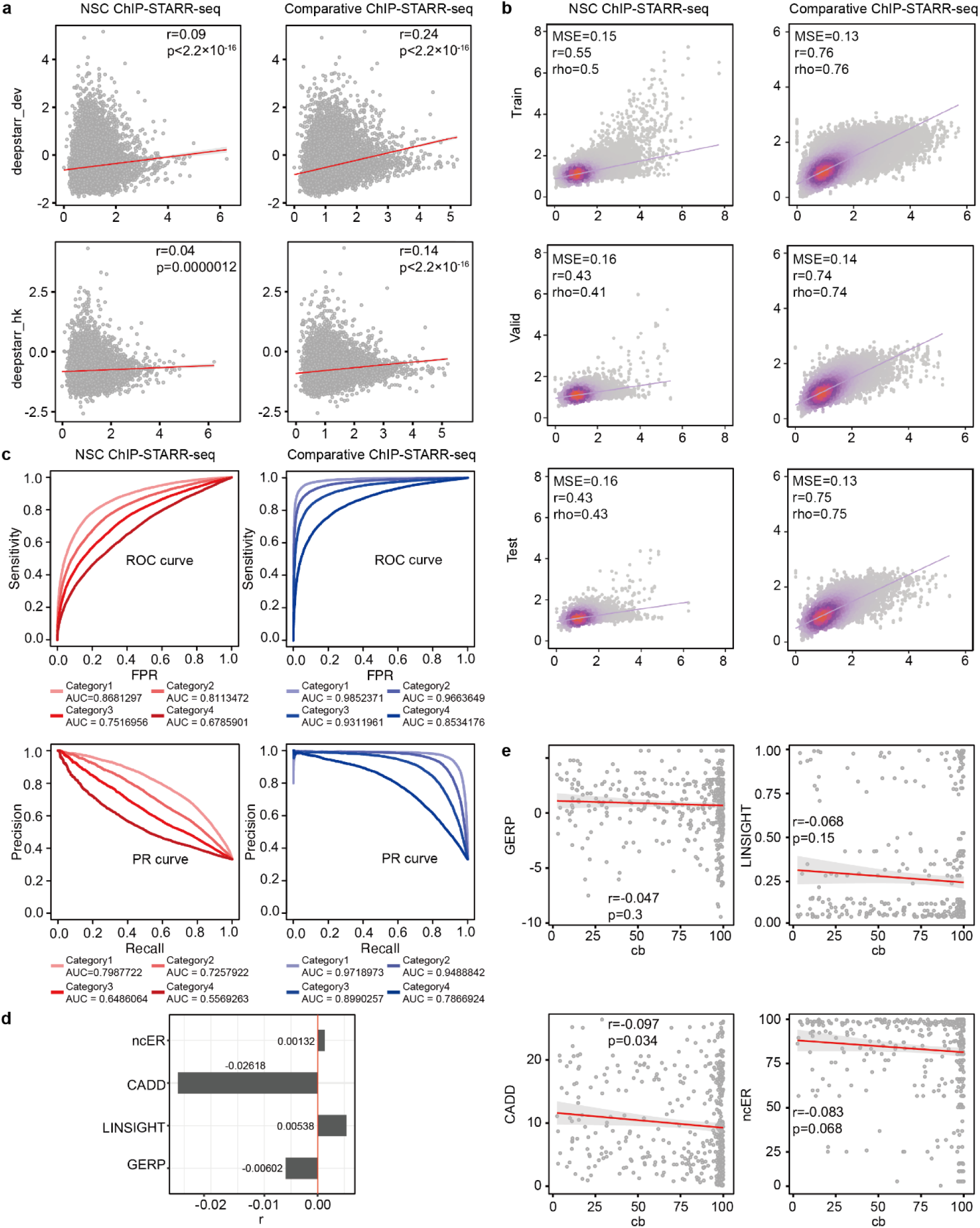
Extended data on BRAIN-MAGNET a) Scatter plot depicting the correlation between DeepSTARR^50^ predictions (using the housekeeping or developmental enhancer DeepSTARR model) of NCRE activity and experimentally obtained ChIP-STARR-seq data from NSC ChIP-STARR-seq or comparative ChIP-STARR-seq. Indicated are the Pearson (r) correlation and p-value. b) Scatter plot indicating relationship between BRAIN-MAGNET predicted and observed NCRE activity scores for each cell type. Also indicated are the mean square error (MSE), Pearson (r) and Spearman (rho) correlation values after concatenating the observations and predictions for the train, valid or test set of data. c) Receiver Operating Characteristic (ROC) (upper panel) and Precision-Recall (PR) curves (lower panel) for a classification task comparing different NCRE activity groups and the top- 10% NCRE activity class using the R package PRROC^78^ for NSC ChIP-STARR-seq (left) and Comparative ChIP-STARR-seq in ESCs (right). The PR curve reflects the precision-recall trade-off, and the ROC curve portrays the true positive and false positive rate trade-off. d) Bar plot indicating the correlation between cb percentiles and GERP, CADD, LINSIGHT and ncER scores for all nucleotides contained in the test set of NCREs. Indicated are the Pearson (r) correlation and p-value. e) Scatter plots depicting the correlations between cb percentiles and GERP, CADD, LINSIGHT and ncER scores for all nucleotides contained in the 15 NCRE motifs that were experimentally studied in **Fig. 3d**. Indicated are the Pearson (r) correlation and p-value.

**Extended Data Fig. 7:**
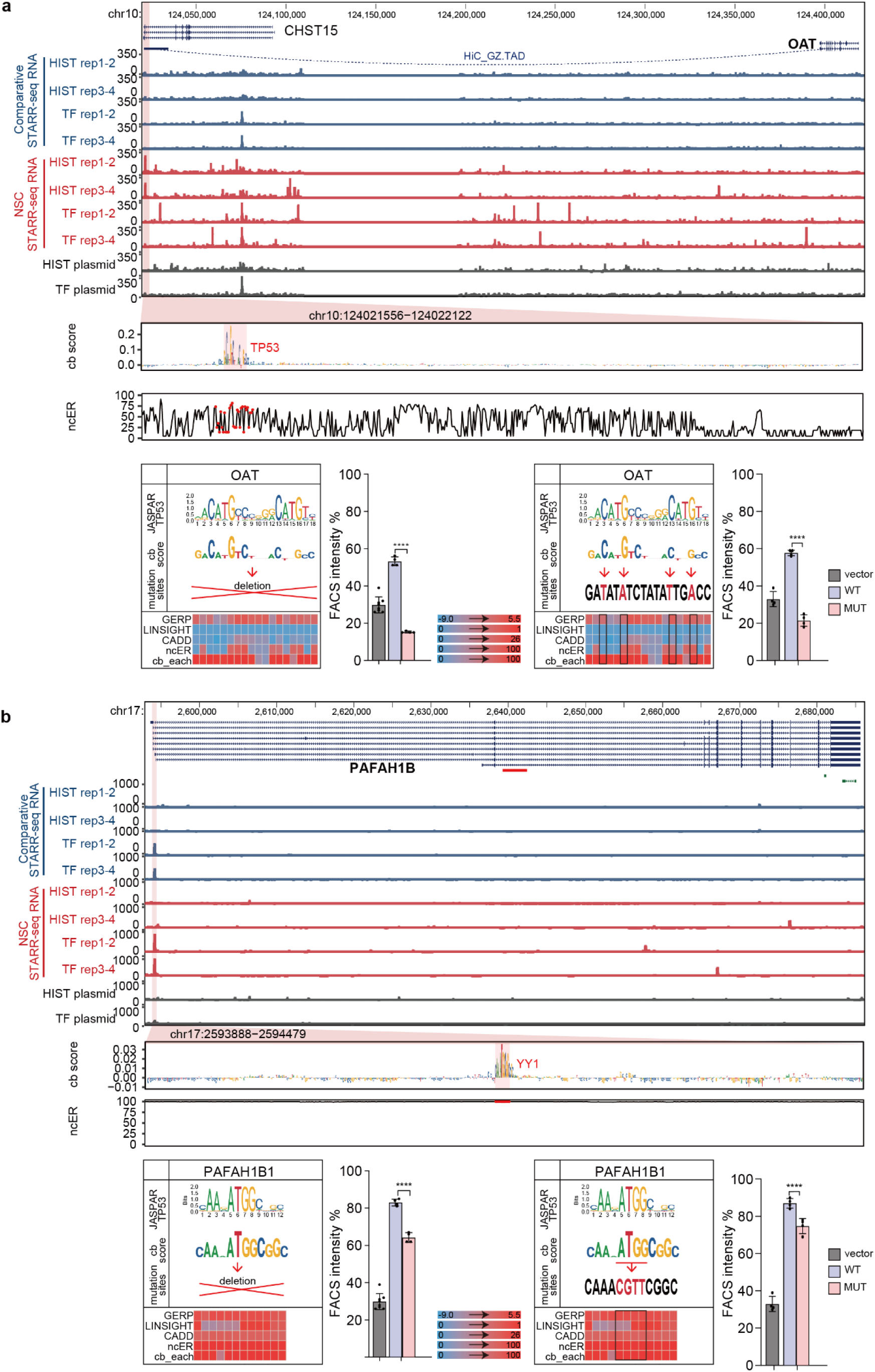
Example loci of *OAT* and *PAFAH1B* for which BRAIN-MAGNET prioritized NCREs and motifs, including functional validation. A) Genome browser view of the *OAT* and *CHST15* locus, showing an NCRE in an intron of *CHST15* that interacts by HiC in foetal brain with *OAT*. Top panel: genome browser tracks showing ChIP-STARR-seq RNA in ESCs (blue) and NSCs (red) and sequencing of ChIP-STARR-seq plasmids (grey); highlighted region in magenta indicates the NCRE. The zoom-in of the NCRE shows the BRAIN-MAGNET cb scores from the NSC model and the ncER scores of the same region. A region with high cb scores and an overlapping TP53 motif is highlighted. Lower panel shows the motif from JASPAR, a visualization of the cb score, the generated deletion (left) or point mutation (right) that was introduced in the NCRE and the outputs per nucleotide of GERP, LINSIGHT, CADD, ncER and cb scores visualized in a heatmap. Bar plots show the results from experimental testing of the created deletion (left) or point mutation (right). Plotted is the percentage of GFP+ in cells co-transfected with an mCherry expressing plasmid, to correct for transfection efficiency. Bars show the average from two independent experiments, with each NCRE tested each in duplicate. Error bars represent standard deviation. * p < 0.05; ** p < 0.01; *** p < 0.001; **** p < 0.0001 (one-way ANOVA test followed by multiple comparison test (Fisher’s LSD test). b) As a, but now for an NCRE in *PAFAH1B* for which BRAIN-MAGNET predicts a functional role for a YY1 motif.

**Extended Data Fig. 8:**
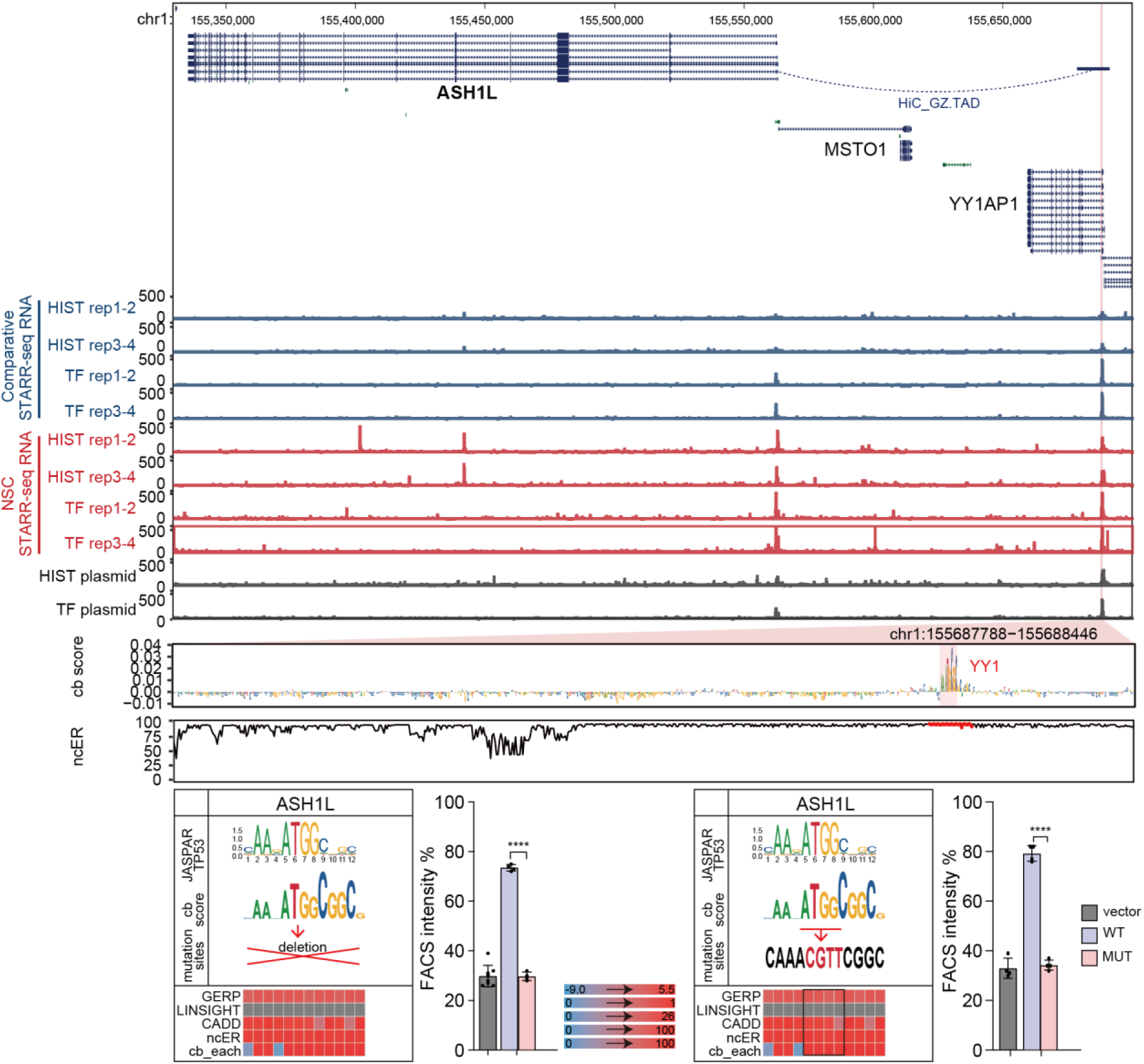
Example locus of *ASHL1*, for which BRAIN-MAGNET prioritized NCRE and motif, including functional validation. Genome browser view of the *ASHL1* locus and flanking genes, showing an NCRE in *YY1AP1* that interacts by HiC in foetal brain with *ASHL1*. Top panel: genome browser tracks showing ChIP-STARR-seq RNA in ESCs (blue) and NSCs (red) and sequencing of ChIP-STARR-seq plasmids (grey); highlighted region in magenta indicates the NCRE. The zoom-in of the NCRE shows the BRAIN-MAGNET contribution (cb) scores from the NSC model and the ncER scores of the same region. A region with high cb scores and an overlapping YY1 motif is highlighted. Lower panel shows the motif from JASPAR, a visualization of the cb score, the generated deletion (left) or point mutation (right) that was introduced in the NCRE and the outputs of per nucleotide GERP, LINSIGHT, CADD, ncER and cb scores visualized in a heatmap. Bar plots show the results from experimental testing of the created deletion (left) or point mutation (right). Plotted is the percentage of GFP+ in cells co-transfected with an mCherry expressing plasmid, to correct for transfection efficiency. Bars show the average from two independent experiments, with each NCRE tested each in duplicate. Error bars represent standard deviation. * p < 0.05; ** p < 0.01; *** p < 0.001; **** p < 0.0001 (one-way ANOVA test followed by multiple comparison test (Fisher’s LSD test).

**Extended Data Fig. 9:**
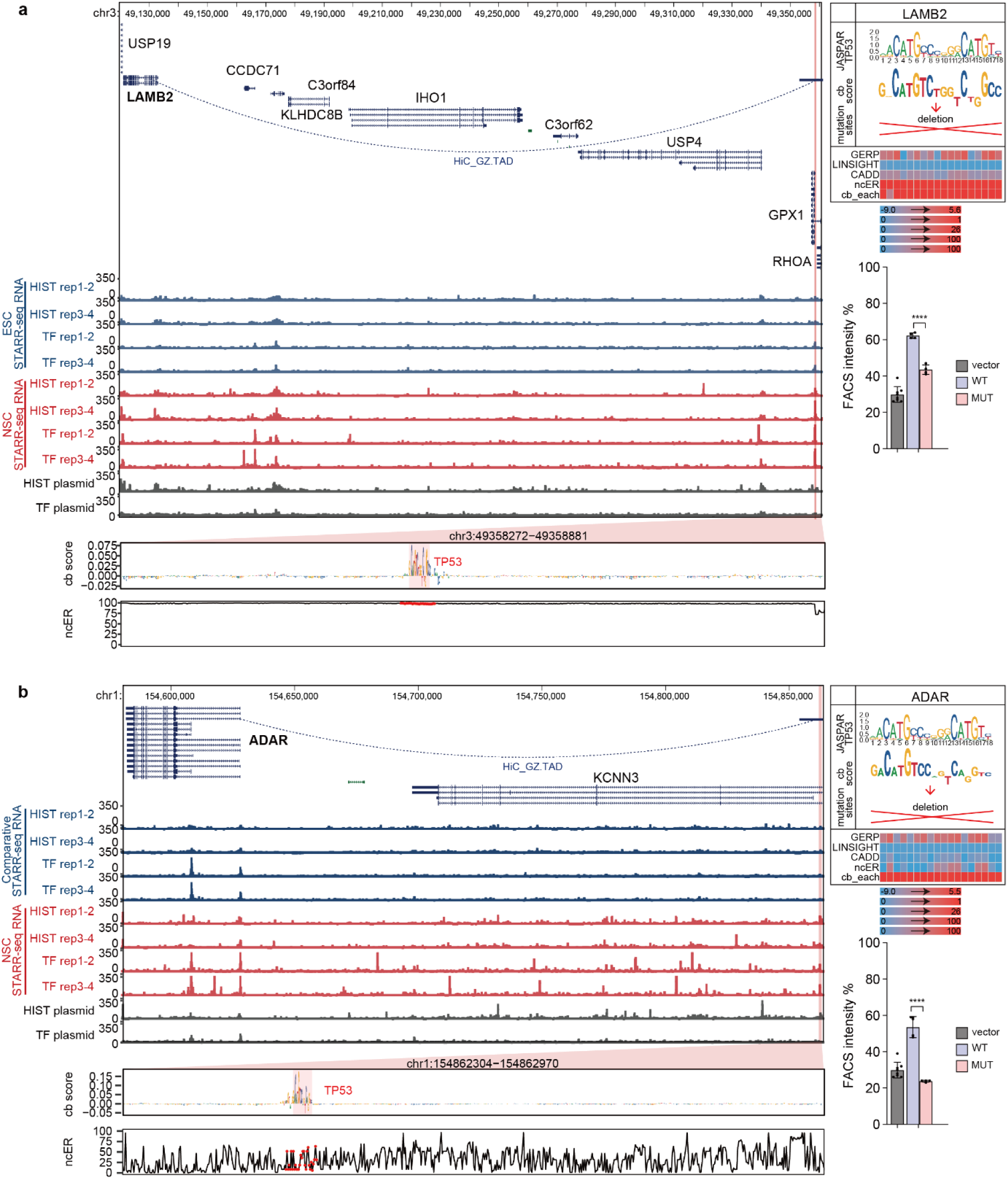
Example loci of *LAMB2* and *ADAR* for which BRAIN-MAGNET prioritized NCREs and motifs, including functional validation. a) Genome browser view of the *LAMB2* locus and flanking genes, showing an NCRE located between *RHOA* and *GPX1* that interacts by HiC in foetal brain with *LAMB2*. Left panel: genome browser tracks showing ChIP-STARR-seq RNA in ESCs (blue) and NSCs (red) and sequencing of ChIP-STARR-seq plasmids (grey); highlighted region in magenta indicates the NCRE. The zoom-in of the NCRE shows the BRAIN-MAGNET contribution (cb) scores from the NSC model and the ncER scores of the same region. A region with high cb scores and an overlapping TP53 motif is highlighted. Upper right panel shows the motif from JASPAR, a visualization of the cb score, the generated deletion that was introduced in the NCRE and the outputs per nucleotide of GERP, LINSIGHT, CADD, ncER and cb scores visualized in a heatmap. Lower bar plots show the results from experimental testing of the created deletion. Plotted is the percentage of GFP+ in cells co-transfected with an mCherry expressing plasmid, to correct for transfection efficiency. Bars show the average from two independent experiments, with each NCRE tested each in duplicate. Error bars represent standard deviation. * p < 0.05; ** p < 0.01; *** p < 0.001; **** p < 0.0001 (one-way ANOVA test followed by multiple comparison test (Fisher’s LSD test). b) As a), but now for an NCRE located in *KCNN3* that interacts by HiC in foetal brain with *ADAR*.

**Extended Data Fig. 10:**
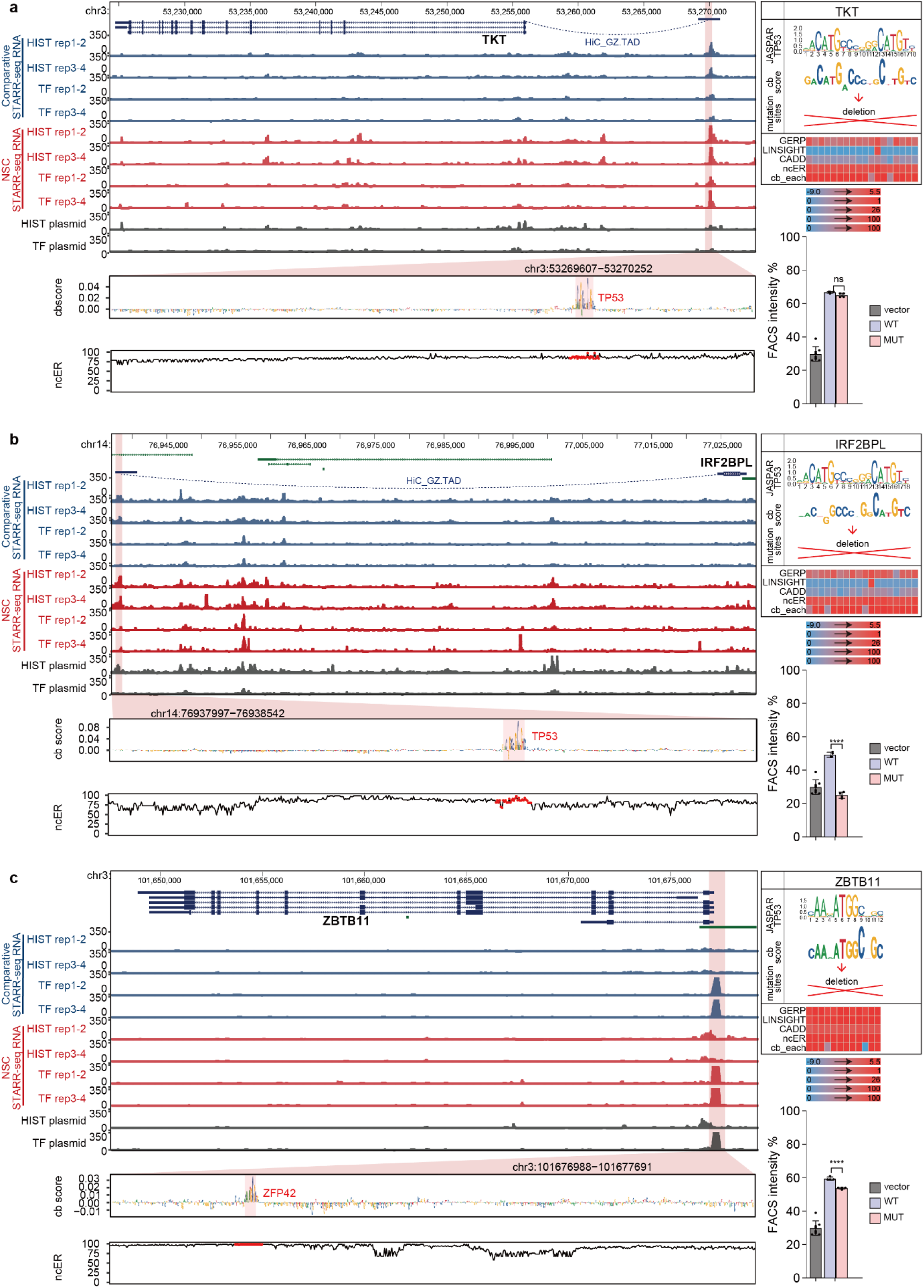
Example loci of *TKT, IRF2BPL* and *ZBTB11* for which BRAIN-MAGNET prioritized NCREs and motifs, including functional validation. a) Genome browser view of the *TKT* locus, showing a distal NCRE that interacts by HiC in foetal brain with *TKT*. Left panel: genome browser tracks showing ChIP-STARR-seq RNA in ESCs (blue) and NSCs (red) and sequencing of ChIP-STARR-seq plasmids (grey); highlighted region in magenta indicates the NCRE. The zoom-in of the NCRE shows the BRAIN-MAGNET contribution (cb) scores from the NSC model and the ncER scores of the same region. A region with high cb scores and an overlapping TP53 motif is highlighted. Upper right panel shows the motif from JASPAR, a visualization of the cb score, the generated deletion that was introduced in the NCRE and the outputs per nucleotide of GERP, LINSIGHT, CADD, ncER and cb scores visualized in a heatmap. Lower bar plots show the results from experimental testing of the created deletion. Plotted is the percentage of GFP+ in cells co-transfected with an mCherry expressing plasmid, to correct for transfection efficiency. Bars show the average from two independent experiments, with each NCRE tested each in duplicate. Error bars represent standard deviation. * p < 0.05; ** p < 0.01; *** p < 0.001; **** p < 0.0001 (one-way ANOVA test followed by multiple comparison test (Fisher’s LSD test). b) As a), but now for a distal NCRE that interacts by foetal brain HiC with *IRF2BPL*. c) As a), but now for an NCRE upstream of *ZBTB11* that contains a ZFP42/YY1 motif.

**Extended Data Fig. 11:**
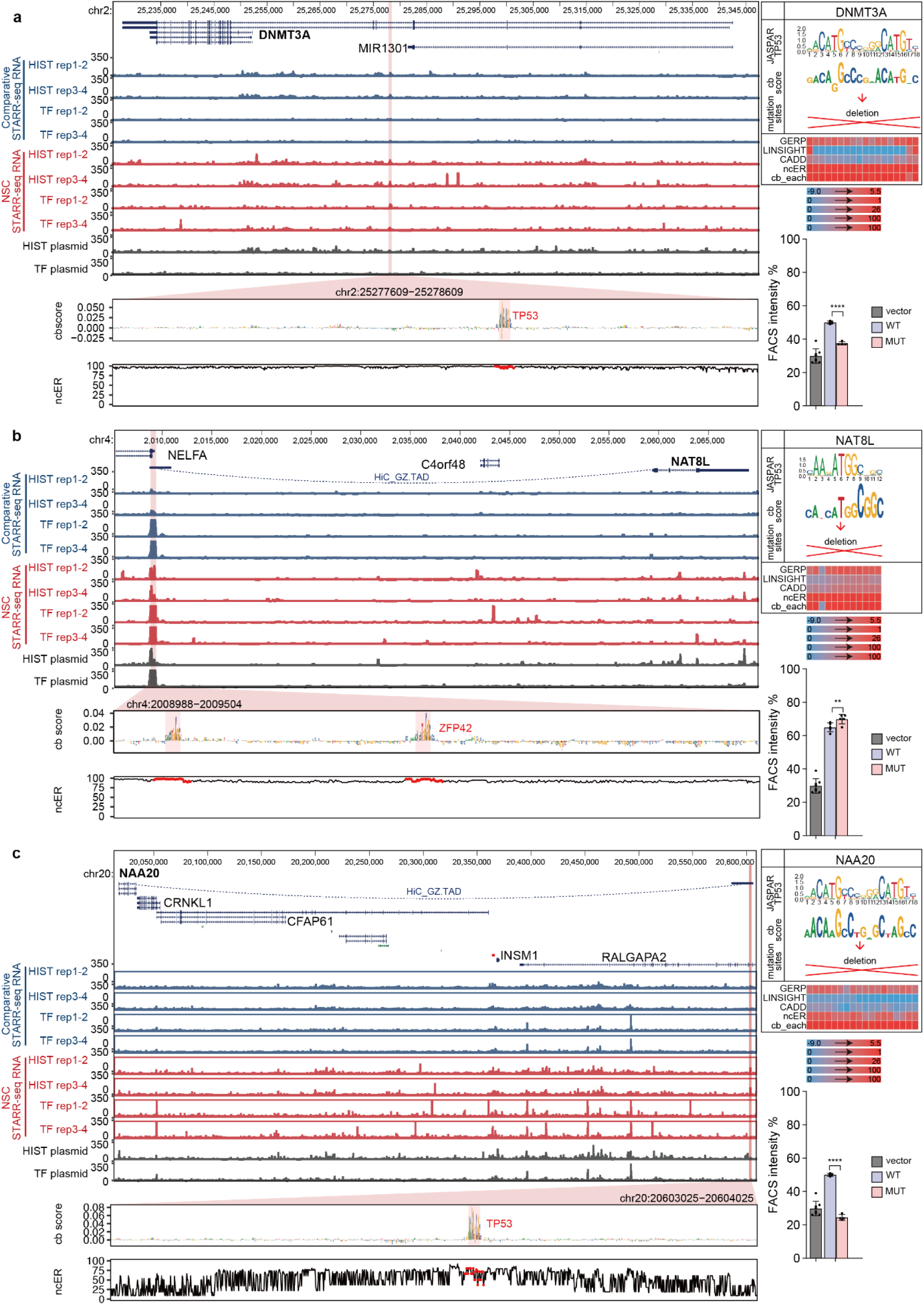
Example loci of *DNMT3A, NAT8L* and *NAA20* for which BRAIN-MAGNET prioritized NCREs and motifs, including functional validation. a) Genome browser view of the *DNMT3A* locus, showing an intronic NCRE. Left panel: genome browser tracks showing ChIP-STARR-seq RNA in ESCs (blue) and NSCs (red) and sequencing of ChIP-STARR-seq plasmids (grey); highlighted region in magenta indicates the NCRE. The zoom-in of the NCRE shows the BRAIN-MAGNET contribution (cb) scores from the NSC model and the ncER scores of the same region. A region with high cb scores and an overlapping TP53 motif is highlighted. Upper right panel shows the motif from JASPAR, a visualization of the cb score, the generated deletion that was introduced in the NCRE and the outputs per nucleotide of GERP, LINSIGHT, CADD, ncER and cb scores visualized in a heatmap. Lower bar plots show the results from experimental testing of the created deletion. Plotted is the percentage of GFP+ in cells co-transfected with an mCherry expressing plasmid, to correct for transfection efficiency. Bars show the average from two independent experiments, with each NCRE tested each in duplicate. Error bars represent standard deviation. * p < 0.05; ** p < 0.01; *** p < 0.001; **** p < 0.0001 (one-way ANOVA test followed by multiple comparison test (Fisher’s LSD test). b) as for a), but now for a distal NCRE located near *NELFA* harboring a ZFP42/YY1 motif, that interacts with *NAT8L* as confirmed by HiC in foetal brain. c) as for a), but now a distal NCRE that interacts by foetal brain HiC with *NAA20*.

**Extended Data Fig. 12:**
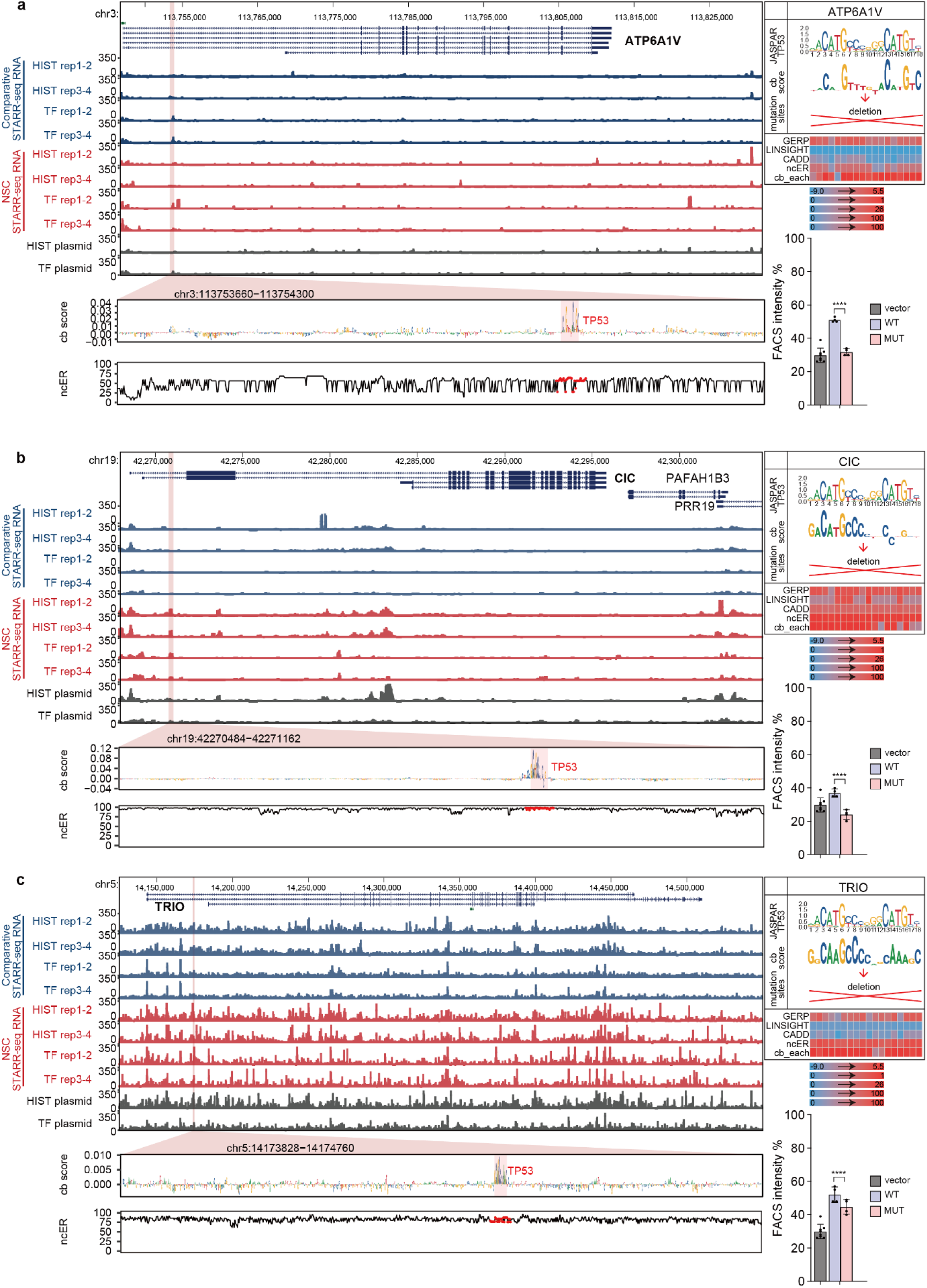
Example loci of *ATP6A1V, CIC* and *TRIO* for which BRAIN-MAGNET prioritized NCREs and motifs, including functional validation. a) Genome browser view of the *ATP6A1V* locus, showing an intronic NCRE. Left panel: genome browser tracks showing ChIP-STARR-seq RNA in ESCs (blue) and NSCs (red) and sequencing of ChIP-STARR-seq plasmids (grey); highlighted region in magenta indicates the NCRE. The zoom-in of the NCRE shows the BRAIN-MAGNET contribution (cb) scores from the NSC model and the ncER scores of the same region. A region with high cb scores and an overlapping TP53 motif is highlighted. Upper right panel shows the motif from JASPAR, a visualization of the cb score, the generated deletion that was introduced in the NCRE and the outputs per nucleotide of GERP, LINSIGHT, CADD, ncER and cb scores visualized in a heatmap. Lower bar plots show the results from experimental testing of the created deletion. Plotted is the percentage of GFP+ in cells co-transfected with an mCherry expressing plasmid, to correct for transfection efficiency. Bars show the average from two independent experiments, with each NCRE tested each in duplicate. Error bars represent standard deviation. * p < 0.05; ** p < 0.01; *** p < 0.001; **** p < 0.0001 (one-way ANOVA test followed by multiple comparison test (Fisher’s LSD test). b) as for a), but now for an intronic NCRE in *CIC*. c) as for a), but now for an intronic NCRE in *TRIO*.

**Extended Data Fig. 13:**
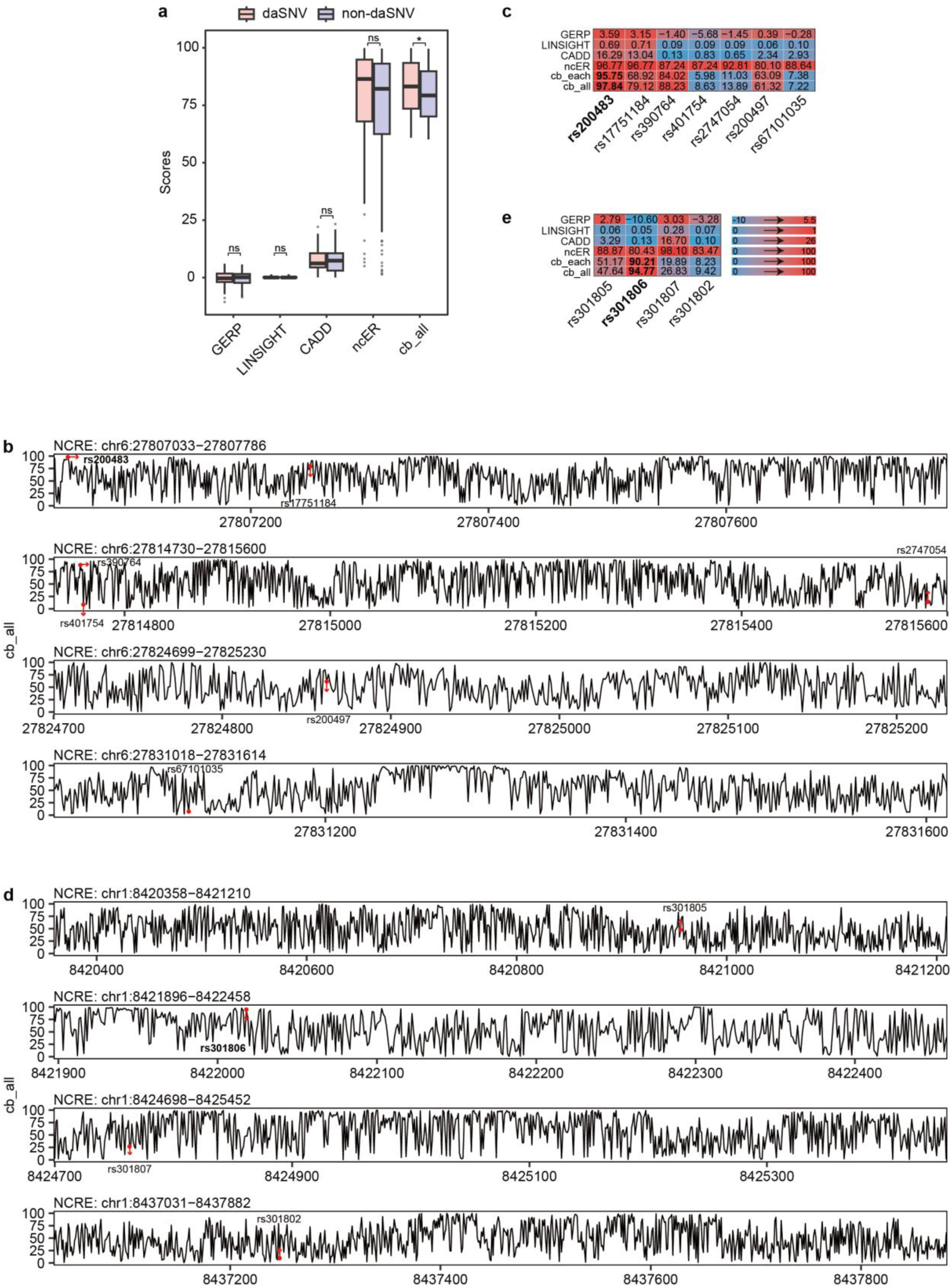
Utility of the NCRE atlas and BRAIN-MAGNET for prioritizing genomic variants to fine map GWAS associated regions a) Box plot showing the distribution of BRAIN-MAGNET cb scores (normalized cb_all >60) for daSNVs (pink) and non-daSNVs (blue) from Guo et al^46^ that overlap with NCREs assessed in our study, and GERP, LINSIGHT, CADD, and ncER scores for the same SNPs. Boxes represent the interquartile range (IQR); lines represent the median; whiskers extend to 1.5 the IQR; dots represent outliers. (* p < 0.05; t.test). b) Zoom-in showing the normalized BRAIN-MAGNET cb scores (cb_all) for the 4 NCREs from our data set that overlap with the GWAS associated risk locus for schizophrenia on chromosome 6. The 7 SNPs in LD and their cb scores are indicated. Of note is that within each NCRE, there are nucleotides which are not common SNPs not assessed in GWAS studies that have higher individual cb scores and might thus impact more on NCRE function if mutant. c) Heatmap depicting the GERP, LINSIGHT, CADD, ncER and BRAIN-MAGNET cb score (cb_all and cb_each) for the 7 SNPs in LD at the schizophrenia associated GWAS locus at chromosome 6 from b). d) As b), but now for the 4 NCREs that overlap with a GWAS associated locus for major depression on chromosome 1. e) As c), but now for the 4 SNPs in LD at the major depression associated GWAS locus on chromosome 1 from d).

**Extended Data Fig. 14:**
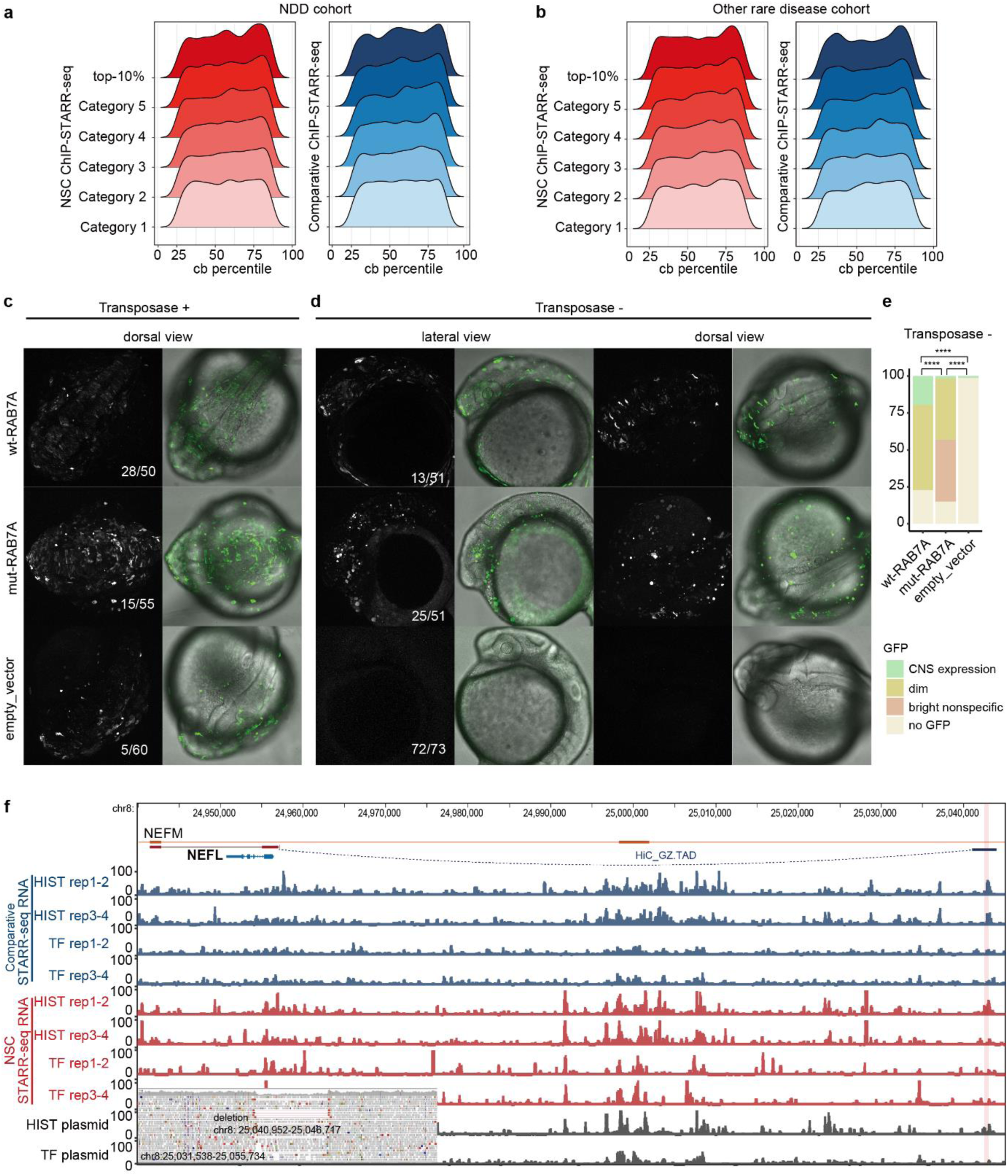
Extended data showing the utility of the NCRE atlas and BRAIN-MAGNET for prioritizing genomic variants found in rare disease a) Density plots depicting the number of *de novo* variants found in 4,415 genetically unexplained individuals with neurological and neurodevelopmental disorders from the Genomics England 100,000 Genomes project, located in NCREs from the different NCRE categories for NSCs (left) or ESCs (right) stratified for their BRAIN-MAGNET cb percentile. b) Density plots depicting the number of *de novo* variants found in 4,558 genetically unexplained individuals with other rare disorders (including cardiovascular disorders, hearing an ear disorders, ultra-rare disorders, dermatological disorders, gastroenterological disorders, renal and urinary tract disorders, respiratory disorders, endocrine disorders, dysmorphic and congenital abnormality syndromes, growth disorders, skeletal disorders, ophthalmological disorders, metabolic disorders, hematological and immunological disorders and rheumatological disorders) from the Genomics England 100,000 Genomes project, located in NCREs from the different NCRE categories for NSCs (left) and ESCs (right), stratified for their BRAIN-MAGNET cb percentile. c) Extended data of NCRE *in vivo* reporter assay in zebrafish at 24 hfp (with transposase). Shown are representative confocal microscopy images of transgenic zebrafish larvae at 24 hpf, generated by injection of empty vector, or transgenic reporter construct containing wild type (wt-RAB7A) or patient-specific mutant *RAB7A* NCRE (mut-RAB7A), in the presence of transposase. Images are shown in dorsal view. Compare to Fig. 5e. d) NCRE *in vivo* reporter assay in zebrafish. Shown are representative confocal microscopy of transgenic zebrafish larvae (generated without transposase) at 24 hpf, in lateral and dorsal view. Numbers indicated in the images present the GFP positive zebrafish larvae per condition. Compared to the wild type, the patient-specific mutant *RAB7A* NCRE showed reduced expression in the central nervous system. Also, a larger proportion of larvae with the patient-specific mutant NCRE showed bright, non-specific expression at aberrant locations not seen with the wild type *RAB7A* NCRE. Together, this confirms that the patient-specific variant *RAB7A* NCR leads to an altered *in vivo* spatiotemporal NCRE expression pattern indicating NCRE dysfunction. d) Quantification of NCRE *in vivo* reporter assay in zebrafish shown in panel d) (****p < 0.0001, Chi-square test). f) Genome browser view showing an NCRE ∼87 kb upstream of *NEFL*, a gene implicated in various forms of autosomal dominant Charcot-Marie-Tooth disease^48^. The NCRE interacts with the *NEFL* promotor region as confirmed by HiC. Top panel: genome browser tracks showing ChIP-STARR-seq RNA in ESCs (blue) and NSCs (red) and sequencing of ChIP-STARR-seq plasmids (grey); highlighted region in magenta indicates the NCRE. The IGV browser view insert shows a heterozygous ∼5.7 kb deletion encompassing the *NEFL* NCRE, identified in a genetically undiagnosed adult with motor and sensory neuropathy. Given the phenotypic overlap with Charcot-Marie-Tooth disease, the NCRE deletion is considered to be possibly disease implicated. No consent for recontacting was obtained, preventing further clinical evaluation and functional studies.

**Extended Data Fig. 15:**
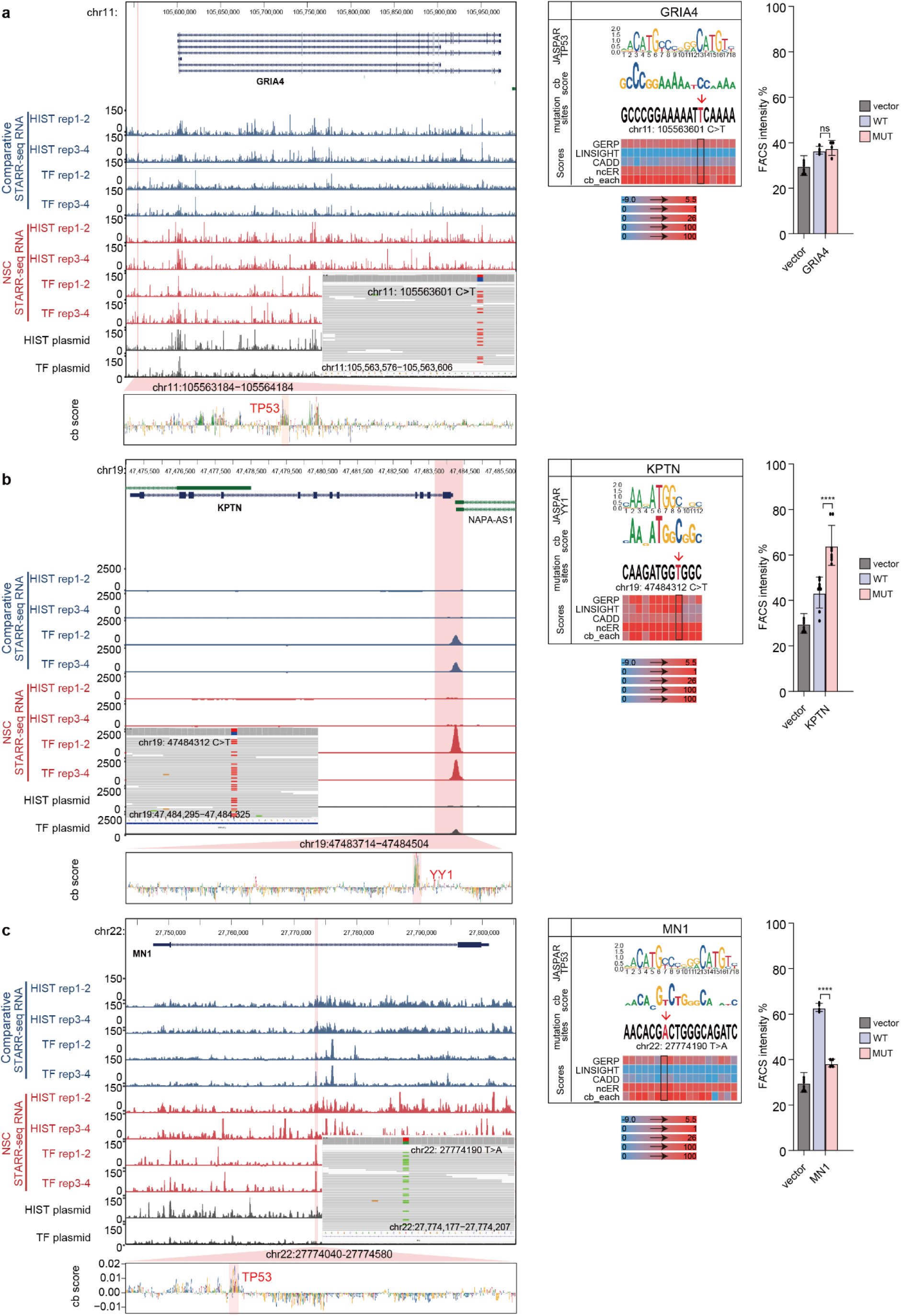
Selected variants from the Genomics England 100,000 Genomes Project for which BRAIN-MAGNET predicts an effect on NCRE activity a) Left panel: Genome browser view of the *GRIA4* locus showing ChIP-STARR-seq RNA for ESCs (blue) and NSCs (red) tracks and plasmid library sequencing results. The identified NCRE is highlighted and the zoom-in shows BRAIN-MAGNET cb scores and predicted functional motifs. The IGV view insert shows a heterozygous rare variant that affects the NCRE predicted functional motifs, detected in a genetically unexplained individual affected with a complex neurological phenotype including ataxia and spasticity. *GRIA4* is linked to autosomal dominant neurodevelopmental disorder with or without seizures and gait abnormalities (OMIM #617864), possibly indicating a phenotypic match. Middle panel shows the motif from JASPAR, a visualization of the cb score and the outputs of GERP, LINSIGHT, CADD, ncER and cb scores visualized in a heatmap for the patient variant and flanking nucleotides. Right bar plots show the results from experimental testing of the patient variant (pink), the wild type NCRE (blue) or the empty plasmid (grey), in which no significant difference for the patient variant was found. Plotted is the percentage of GFP+ in cells co-transfected with an mCherry expressing plasmid, to correct for transfection efficiency. Bars show the average from two independent experiments, with each NCRE tested each in duplicate. Error bars represent standard deviation. * p < 0.05; ** p < 0.01; *** p < 0.001; **** p < 0.0001; ns, not significant (one-way ANOVA test followed by multiple comparison test (Fisher’s LSD test). b) As a), but now for the *KPTN* locus. *KPTN* is linked to autosomal recessive intellectual developmental disorder 41 (OMIM #615637). The rare NCRE variant was identified in two unrelated individuals affected by epilepsy and ataxia, and significantly increases the NCRE activity. We cannot exclude that potentially increased downstream *KPTN* expression might represent a novel disease mechanism linked to this gene. c) As a), but now for the *MN1* locus. *MN1* is linked to autosomal dominant CEBALID syndrome, (OMIM #618774), that presents with dysmorphic features, intellectual disability, and brain malformations. The identified NCRE variant which significantly reduced NCRE activity was found in an unexplained individual reported to have a complicated epilepsy phenotype with amongst other features abnormalities of the cerebral cortex possibly presenting a (partial) phenotypic match.

**Extended Data Fig. 16.**
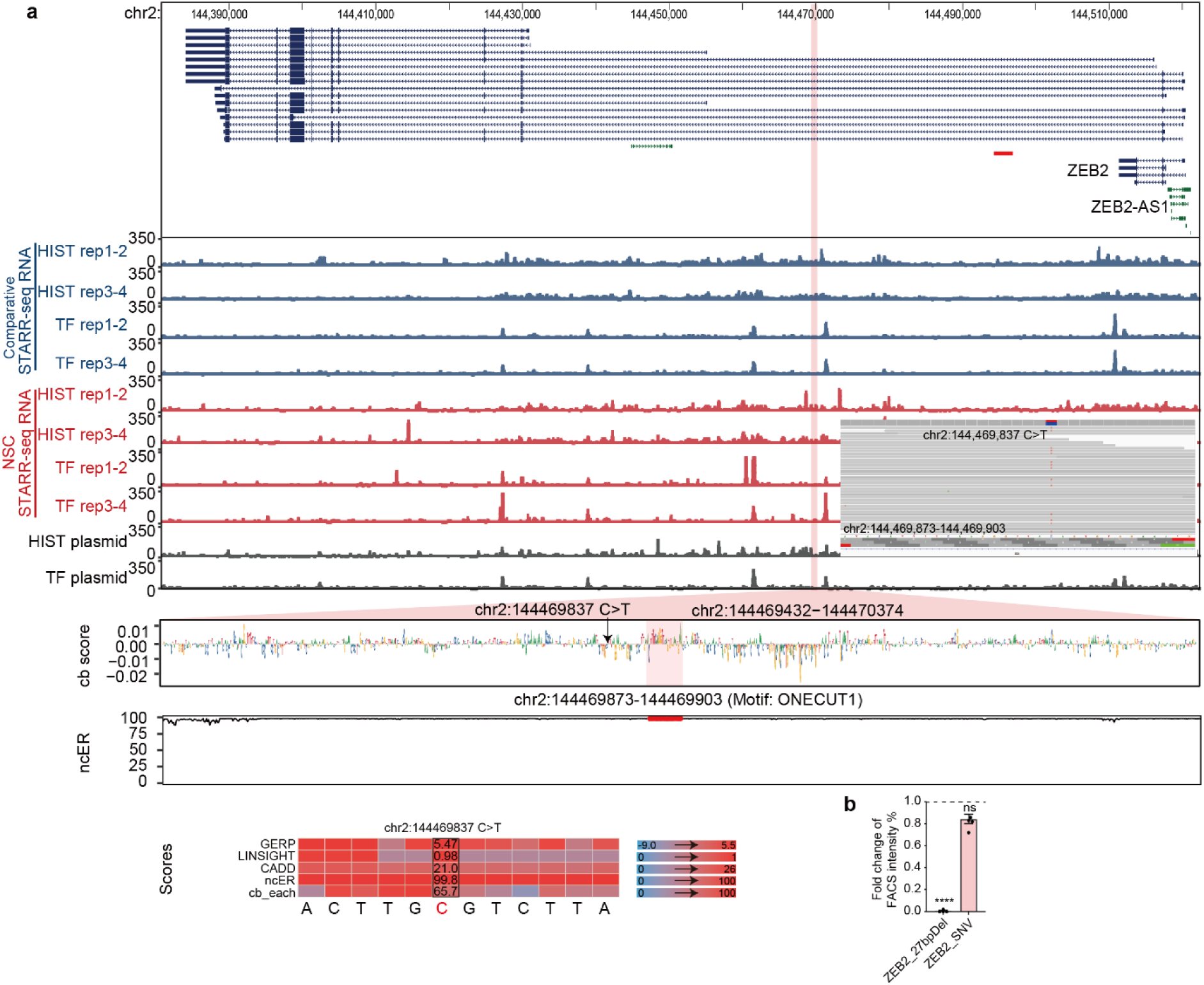
BRAIN-MAGNET analysis eliminates a clinically suspected candidate disease causing non-coding variant in Mowat-Wilson syndrome a) Genome browser view of the *ZEB2* locus, showing an intronic NCRE in which a rare variant was found in a genetically unexplained individual clinically suspected of Mowat-Wilson syndrome. Top panel: genome browser tracks showing ChIP-STARR-seq RNA in ESCs (blue) and NSCs (red) and sequencing of ChIP-STARR-seq plasmids (grey); highlighted region in magenta indicates the NCRE. The zoom-in of the NCRE shows the BRAIN-MAGNET contribution (cb) scores from the NSC model and the ncER scores of the same region. A region with high cb scores and an overlapping ONECUT1 motif and the mutation site of the variant found in the affected individual are highlighted. The IGV browser view show the heterozygous variant identified in the individual. The heatmap below depicts the outputs of GERP, LINSIGHT, CADD, ncER and cb scores for the patient variant and its flanking nucleotides. b) Bar plot showing the results from experimental testing of the created deletion of the ONECUT1 motif or the NCRE variant identified in the affected individual. Plotted is the percentage of GFP+ in cells co-transfected with an mCherry expressing plasmid, to correct for transfection efficiency. Bars show the average from two independent experiments, with each NCRE tested each in duplicate. Error bars represent standard deviation. * p < 0.05; ** p < 0.01; *** p < 0.001; **** p < 0.0001 (one-way ANOVA test followed by multiple comparison test (Fisher’s LSD test).

